# Genomic analyses reveal new insights into Alzheimer’s disease

**DOI:** 10.1101/2025.10.10.25337470

**Authors:** Emil Uffelmann, Douglas P. Wightman, Shahram Bahrami, Alexey A. Shadrin, Vera Fominykh, Takafumi Ojima, Chenyang Jiang, Christian Benner, Elisa Moreno, Adrian I. Campos, Jesper Q. Thomassen, Emmanuel Minois-Genin, Hei Man Wu, G. Bragi Walters, Richard Sherva, Tian Lin, Xuemin Wang, Julien Bryois, Kristi Krebs, Marijn Schipper, Akira Narita, Alessandro Serretti, Anja H. Simonsen, Anna L. van Seumeren, Anne Corbett, Anne-Brita Knapskog, Annette M. Hartmann, Anouk den Braber, Argonde C. van Harten, Arvid Harder, Arvid Rongve, Bengt O. Madsen, Betty M. Tijms, Bitten Aagaard, Bjørn Lichtwarck, Bjørn E. Kirsebom, Byron Creese, Chandra A. Reynolds, Sara Hägg, Ida Karlsson, Christian Erikstrup, Christina Mikkelsen, Clive Ballard, Dag Aarsland, Daichi Shigemizu, Dan Rujescu, Daniel Gudbjartsson, Eivind Aakhus, Erik Sørensen, Eystein Stordal, Flora H. Duits, Frank J. Wolters, Frederic Blanc, Geert Jan Biessels, Geir Selbæk, Geir Bråthen, Gen Tamiya, Gunhild Waldemar, Harro Seelaar, Helga Eyjolfsdottir, Henne Holstege, Henning Bundgaard, Henrik Zetterberg, Henrik Ullum, Ina Giegling, Ingmar Skoog, Ingrid T. Medbøen, Ingvild Saltvedt, Irena Rektorova, J. Michael Gaziano, Jan Haavik, Jens Hjerling-Leffler, Jiao Luo, Jon Snaedal, Everard G.B Vijverberg, Julia M. Sealock, Kaj Blennow, Kaja Nordengen, Karin Persson, Katja Scheffler, Koichi Matsuda, Kouichi Ozaki, Lasse Pihlstrøm, Lavinia Athanasiu, Lene Pålhaugen, Marc Hulsman, Margda Waern, Maria Averina, Marianne Wettergreen, Marta R. Moksnes, Martijn Huisman, Masayuki Yamamoto, Mathias Toft, Matthew S. Panizzon, Mie Topholm Bruun, Mohsen Ghanbari, Monique Franc, Nancy L. Pedersen, Nathaniel Y. Bell, Niccoló Tesi, Ole B. Pedersen, Oleksandr Frei, Olivier Bousiges, Per Svenningsson, Pieter J. Visser, Qingqin S. Li, Richard Hauger, Rui Zhang, Shinichi Namba, Sigrid B. Sando, Silke Kern, Srdjan Djurovic, Steinunn Thordardottir, Tanya N. Phung, Thomas Truelsen, Thomas Werge, Thomas F. Hansen, Tomoki Kyosaka, Torgeir Engstad, Tormod Fladby, Victoria Merritt, Sverre Bergh, Wiesje M. van der Flier, Rujin Wang, Eli A. Stahl, Basavaraj Hooli, 23andMe Research Institute, LifeLines Cohort Study, DBDS Genomic consortium, Regeneron Genetics Center, Penn Medicine Biobank, GHS-RGC DiscovEHR collaboration, Mayo Clinic-RGC Project Generation, Colorado Center for Personalized Medicine – RGC Collaboration, UCLA-RGC ATLAS collaboration, INDIANA-CHALASANI, Mount Sinai Million Health Discoveries Program, Estonian Biobank research team, VA Million Veteran Program, Lea K. Davis, Mark W. Logue, Kelli Lehto, Anna Zettergren, Ben M. Brumpton, Jian Zeng, Peter M. Visscher, Paul F. O’Reilly, Anubha Mahajan, Manuel Ferreira, Yukinori Okada, Sven J. van der Lee, Sisse R. Ostrowski, Ruth Frikke-Schmidt, Hreinn Stefansson, Karl Heilbron, Ole A. Andreassen, Danielle Posthuma

## Abstract

Alzheimer’s disease (AD) is the most common cause of dementia, with global case numbers projected to reach 153 million in 2050^1^. AD is highly heritable, with twin-based heritability estimates of 60-80%^2^. While 1,200 causal loci are predicted to exist for AD^3^, approximately 80 have been associated with AD in two recent studies^4,5^, suggesting that many loci remain to be discovered^6^. Here, we analyzed data from 109,479 cases, 74,141 proxy cases, 2,131,799 controls, and 499,708 proxy controls from diverse ancestries, identifying 118 loci in a multi-ancestry analysis and 9 additional loci in ancestry-specific analyses, 48 of which are new. We identified new AD risk genes, prioritized potential drug targets, and identified microglia and, for the first time, several neuronal cell types enriched for AD-associated genetic risk. Moreover, we improved polygenic prediction and estimated a single-nucleotide polymorphism (SNP) heritability of 16%. Together, our findings offer insights into the genetic architecture and potential pathobiology of AD, as well as specific targets for future drug development research.

## Introduction

Alzheimer’s disease (AD) is the most common cause of dementia, affecting tens of millions of people worldwide and projected to more than double by 2050^1^. Current disease-modifying treatments for AD typically target only one aspect of its pathology: amyloid beta aggregates. However, this has shown limited efficacy in alleviating symptoms^7,8^. Further insight into the disease etiology may highlight other mechanisms leading to the development of more effective drugs. Given the large twin-based heritability of AD 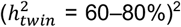, genetic studies can provide valuable insights into its underlying biological mechanisms. Moreover, polygenic scores (PGS) may improve clinical risk prediction models and help identify individuals for early intervention before the onset of neurodegenerative processes.

Recent genome-wide association studies (GWAS) of AD have successfully identified 75 risk loci, highlighted the central role of microglia, and suggested immune function, amyloid catabolism, and lipid function as important biological processes^4,5^. Despite these discoveries, the PGS generated from these studies have had limited predictive performance^4^, and liability-scale summary-statistics-based heritability 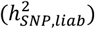 estimates have been low (∼5%)^9^. This discrepancy has prompted the suggestion that there may be little additional genetic risk that can be identified with GWAS^6^ beyond the strongest genetic risk factor for AD, *APOE*. Others, however, proposed that approximately 1,200 causal loci are expected to influence genetic risk for AD^3^. To reconcile these differences, larger studies leveraging improved, well-calibrated methodologies will be critical. Such efforts may yield more accurate heritability estimates, enhance the predictive power of PGS, and, importantly, uncover additional risk genes informing disease mechanisms. Moreover, including individuals of non-European ancestry can further improve PGS prediction in underrepresented populations.

Here, we report the largest multi-ancestry GWAS of AD to date, encompassing a total of 109,479 cases, 74,141 proxy cases, 2,131,799 controls, and 499,708 proxy controls. Proxy cases are genotyped individuals who reported their parents to have had AD (see **Methods**). Using this sample, we identified 118 loci in a multi-ancestry analysis and 9 additional loci in ancestry-specific analyses, 48 of which are new. We estimated a 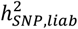 of 16% and improved the out-of-sample prediction. We also found enrichment in microglia and, for the first time, in neuronal cell types, and highlighted relevant biological processes and potential drug targets. To maximize the utility of our results, we have provided publicly available summary statistics from the main analyses, as well as from multiple stratified analyses.

## Results

### Data overview

Our study included 109,479 AD cases, 74,141 proxy AD cases, 2,131,799 controls, and 499,708 proxy controls across 30 cohorts (see **Figure 1 Supplementary Figure 1**, and **Supplementary Data 1**), resulting in a total effective sample size^10^ (*N*_*eff*_ ; see **Methods**) of 416,493. The total effective sample size consisted of 91% individuals of European (EUR), 5% African (AFR), 3% East Asian (EAS), <1% Admixed-American (AMR), and <1% South Asian (SAS) ancestry (**Supplementary Figure 1**). A subset of this dataset was available for sex-stratified analyses, with 20,402 female cases and 261,889 female controls, and 15,689 male cases and 214,956 male controls. Another subset with a *N*_*eff*_ of 196,822 was available for a GWAS on the X chromosome. A full description of each cohort is available in the **Supplementary Methods**.

**Figure 1.**
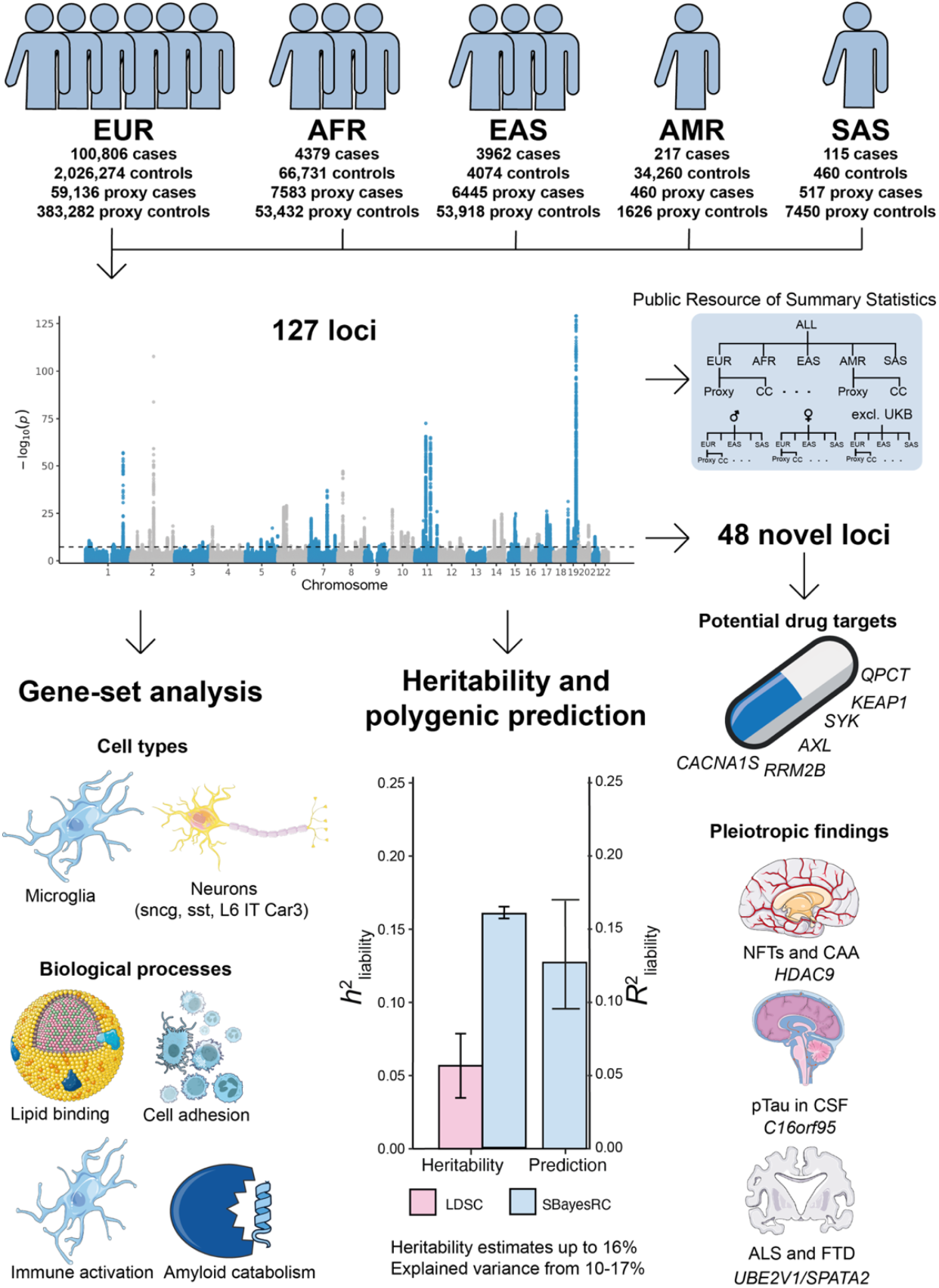
Overview of study design and key findings. Flowchart summarizing the workflow and main results, including genome-wide association analysis (Manhattan plot from our multi-ancestry GWAS), gene-set enrichment, SNP heritability and polygenic risk prediction, and characterization of new loci. NFT = neurofibrillary tangles, CAA = cerebral amyloid angiopathy, pTau = phosphorylated tau, CSF = Cerebrospinal Fluid, ALS = amyotrophic lateral sclerosis. FTD = frontotemporal dementia.

### GWAS meta-analyses identify new loci

We conducted a multi-ancestry GWAS of AD, including five broad ancestry groups: EUR (*N*_*eff*_ = 379,465), AFR (*N*_*eff*_ = 21,163), EAS (*N*_*eff*_ = 13,790), AMR (*N*_*eff*_ = 1,222), and SAS (*N*_*eff*_ = 852) (see **Figure 2** and **Supplementary Data 1**). After QC, this resulted in 8,152,095 SNPs with a minimum minor allele frequency (MAF) of 0.004 and a mean of 0.17. We identified 118 significant loci (*p* < 5×10^-8^), 41 of which are new. We did not identify risk loci on the X chromosome. In the ancestry-specific analyses, we identified 114 significant loci (38 new) in the EUR GWAS, 3 loci in AFR GWAS (*TREM2, SORL1, APOE*), 2 loci in EAS GWAS (*SORL1* and *APOE*), and no loci in SAS or AMR GWAS (**Supplementary Figure 2**). The discrepancy in identified loci is as expected based on differences in sample size, with more than 90% of the total sample consisting of EUR individuals. After meta-analyzing the ancestry specific results, we identified 118 loci. By meta-analyzing the ancestries, an additional 13 loci were identified, while 9 of the loci identified in the EUR only analysis dropped below the significance threshold (*TMEM163, MAP3K13, GPAM, MCF2L, KIAA0125*/IGH gene cluster, *GLCE, MCTP2, MNT*/*SGSM2*/*DPH1*/*RAP1GAP2, AC104532*.*2*/*VMAC*). These 9 loci were still suggestive (at least *p* < 1×10^-5^) in the multi-ancestry meta-analysis. Overall, we identified 127 loci across all meta-analyses; 118 in the multi-ancestry analysis and an additional 9 in the EUR only analysis (**Supplementary Data 2**). Of the 127 loci, 48 were new (see **Supplementary Data 3** and **4** for previously published studies and loci). As a sensitivity analysis to address potential heterogeneity in effect sizes across ancestries for all loci except APOE, we conducted a random-effects meta-analysis between ancestries using METASOFT^11^ after performing a fixed-effects meta-analysis within each ancestry with METAL. The P-values of the lead SNPs remained almost unchanged (see **Methods** and **Supplementary Figure 3**).

**Figure 2.**
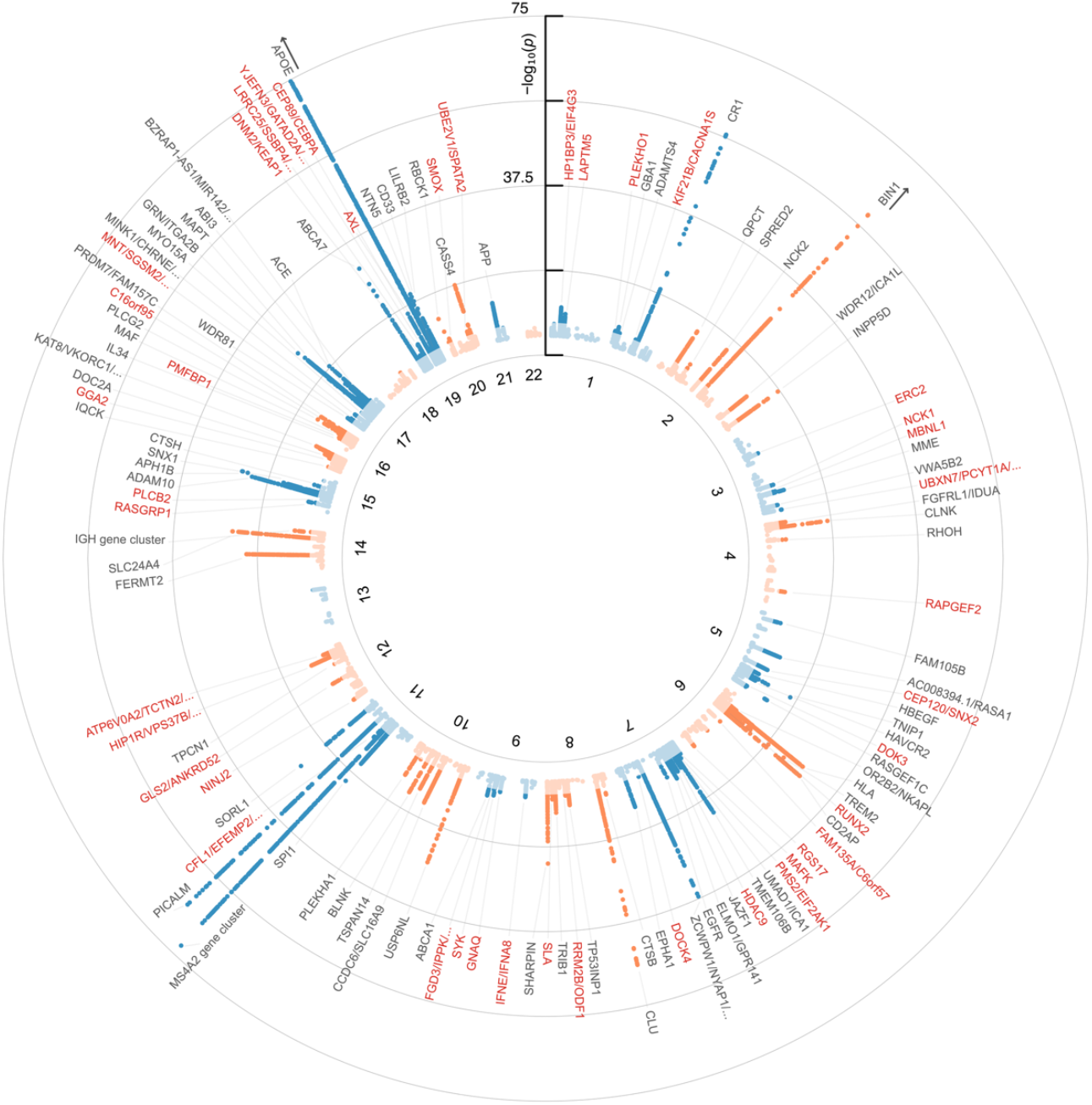
Circular Manhattan plot of the multi-ancestry GWAS. The x-axis represents the chromosomal position of SNPs, and the y-axis their strength of association measured as –log_10_(*p*). Genome-wide significant (*p* < 5×10^-8^; darker points) loci are annotated with their predicted effector gene. New genes are highlighted in red. Lighter-shaded points correspond to SNPs not reaching genome-wide significance. Only SNPs with *p* < 0.00005 are plotted. The y-axis is capped at 75 for visual clarity; however, the APOE locus reaches –log_10_(*p*) ∼ 5697 and the BIN1 locus reaches –log_10_(*p*) = 108.

We also meta-analyzed the data stratified by sex (male vs. female) and phenotype definition (case-control vs. proxy). The sex-stratified GWAS (excluding proxy cases) identified 12 loci in the female-only GWAS (*CR1, BIN1, OR2B2/NKAPL, TREM2, CLU, MS4A2, PICALM, APH1B, MAPT, ABCA7, APOE, LILRB2*). We identified six loci in the male-only GWAS (*CR1, BIN1, MS4A2, PICALM, APOE, NTN5*) (**Supplementary Figure 4**). The relatively low number of associated loci compared to the overall GWAS was expected given the much smaller effective sample size of the sex-stratified analyses (*N*_*eff, female*_ = 64,771; *N*_*eff, male*_ = 49,748). The *APOE-ε4* tagging SNP had a 13% larger odds ratio in females (2.85 vs. 2.52, *p* = 9 × 10^−8^). We estimated the genetic correlation (*r*_*g*_) between EUR males and females with LD-Score Regression (LDSC) at 0.77 (95%-CI: 0.48 - 1.06). Likely due to small sample size and insufficient heritability, LDSC failed to compute genetic correlations for all other ancestries. Because the standard error of LDSC’s *r*_*g*_ scales inversely with the SNP heritabilities of the contributing traits, and LDSC appears to underestimate AD’s 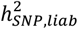 (see below), we also approximated a more precise lower bound of the genetic correlation of 0.82 (95%-CI: 0.76 – 0.88) between males and females in EUR using SBayesRC (see **Methods**).

The case-control only meta-analysis across all ancestries (*N*_*eff*_ = 353,189) identified 95 unique significant loci, and the proxy-only meta-analysis (*N*_*eff*_ = 60,826) identified 12 significant loci (**Supplementary Figure 5**). Ten of the 12 proxy loci were also genome-wide significant in the case-control meta-analysis, while the other 2 loci were borderline significant (*p* = 6×10^-8^; *p* = 9×10^-6^). The LDSC-computed genetic correlation between the case-control and proxy meta-analysis was 0.99 (s.e. = 0.14, *p* = 3×10^-13^), indicating high similarity (see **Supplementary Note 1**). A summary of association across the 127 loci in the ancestry, sex, and phenotype stratified meta-analyses is available in **Supplementary Data 5** and **Supplementary Figures 6** and **7**. The multi-ancestry meta-analysis incorporated several cohorts not included in previous AD GWAS^4,5^, allowing us to assess the replication of recent GWAS findings in samples independent from previously included samples. Overall, we found high replicability of loci (**Supplementary Note 2** and **Supplementary Data 6** and **7**), and that lead-SNP effect sizes correlated strongly (*r* = 0.98; see **Supplementary Figure 8**). To maximize the value of the data collected, we have made the summary statistics from the primary and stratified meta-analyses publicly available (after the exclusion of 23andMe Research Institute data). A summary table of the available summary statistics can be found in **Supplementary Data 8**.

We observed heterogeneous APOE-ε4 effect sizes across ancestries (see **Supplementary Figure 9**), ranging from the smallest odds ratio (OR) in AFR (OR = 1.7) to the largest in EAS (OR = 3.4). We note the APOE-ε4 variant was not present in All of Us, the source of all AMR participants (see **Supplementary Methods**). This variation is not explained by differences in proxy case representation: we find no consistent relationship between proxy case definitions and APOE-ε4 effect size, with proxy-based estimates larger than genotyped-case estimates in EUR and AFR, but smaller in EAS and SAS. However, interpretation is complicated by substantial cross-cohort heterogeneity in APOE-ε4 effect sizes even within European cohorts (*I*^2^ = 97.7%, p = 8.19×10^−210^; see **Supplementary Figure 10**), suggesting that cohort-level differences may account for much of the observed variation rather than true differences in APOE-ε4 penetrance across ancestries.

To ass the impact of three cohorts that did not adjust for genotyping array or batch, we repeated the meta-analysis excluding them. Although 29 loci were lost, all were near the significance threshold both before and after exclusion, consistent with reduced power rather than false-positive signal in the primary analysis (**Supplementary Note 3**).

### Effector gene prediction for 127 loci

For each locus, we predicted the most likely effector genes based on several methodologies and sources of evidence. We nominated genes in a locus based on associations in previous exome-wide association studies (ExWAS) of AD (see **Supplementary Data 4**), effector gene predictions from a machine learning model (FLAMES)^12^, colocalization^13^ and Mendelian randomization (MR)^14^ results using expression, splicing, and protein quantitative trait loci (eQTL, sQTL, and pQTL, respectively) datasets (see **Supplementary Data 9**), and gene nominations from previous GWAS (see **Supplementary Data 3**) (see **Methods**). We aimed to select a single gene per locus, prioritizing ExWAS-identified genes followed by FLAMES predictions. If these were not available for a locus, we selected the genes reported in previous GWAS. For new loci without ExWAS and FLAMES predictions, we selected genes from the colocalization and Mendelian randomization analysis and assessed their evidence in previous literature. A full description of the new loci is available in the **Supplementary Note 4**. For loci with conflicting evidence, we additionally searched the literature for experimental evidence for the implicated genes (**Supplementary Note 5**). A summary of the experimental evidence from the literature for these genes is available in **Supplementary Data 10**. For some loci, this approach did not consistently resolve the conflicting predictions; in such cases, we report multiple genes (**Table 1**; **Supplementary Data 2**).

**Table 1:** Genome-Wide Significant Loci and Effector Gene Predictions The novel significant loci (*p*<5×10^-8^) that were identified across the multi-ancestry and EUR only meta-analyses. The loci marked with an asterisk were only significant in the EUR-only meta-analysis. More information for all known and novel loci is available in **Supplementary Data 2**. chr = chromosome, A1 = the tested allele, A1 freq = tested allele frequency, OR = odds ratio, 95% CI = 95% confidence interval around the OR. Loci with more than two effector genes list the first two followed by ellipses.

| locus | predicted effector gene(s) | chr | start | end | lead SNP | A1 | A1 freq | OR | 95% CI | p-value |
| --- | --- | --- | --- | --- | --- | --- | --- | --- | --- | --- |
| 1 | HP1BP3/EIF4G3 | 1 | 20815167 | 21785330 | rs12030051 | A | 0.55 | 1.04 | 1.02-1.05 | $2.17 \times 10^{-11}$ |
| 2 | LAPTM5 | 1 | 30965364 | 31504273 | rs34854980 | A | 0.19 | 1.04 | 1.03-1.06 | $5.48 \times 10^{-10}$ |
| 3 | PLEKHO1 | 1 | 149749228 | 150399016 | rs6700343 | A | 0.069 | 0.94 | 0.92-0.96 | $2.09 \times 10^{-8}$ |
| 6 | KIF21B/CACNA1S | 1 | 200750305 | 201264966 | rs67602696 | T | 0.28 | 1.03 | 1.02-1.04 | $4.09 \times 10^{-8}$ |
| 15 | ERC2 | 3 | 56023512 | 56523512 | rs11926826 | A | 0.067 | 1.07 | 1.04-1.09 | $3.50 \times 10^{-8}$ |
| 16 | NCK1 | 3 | 136371007 | 136921504 | rs1347209 | T | 0.60 | 1.03 | 1.02-1.04 | $1.11 \times 10^{-8}$ |
| 17 | MBNL1 | 3 | 151845708 | 152437279 | rs892667 | A | 0.68 | 1.03 | 1.02-1.04 | $1.09 \times 10^{-8}$ |
| 20* | MAP3K13 | 3 | 184871143 | 185405925 | rs190301053 | A | 0.02 | 0.9 | 0.86-0.93 | $2.21 \times 10^{-8}$ |
| 21 | UBXN7/PCYT1A/... | 3 | 195981308 | 196483136 | rs34547573 | A | 0.26 | 0.97 | 0.95-0.98 | $4.45 \times 10^{-9}$ |
| 25 | RAPGEF2 | 4 | 159456526 | 160107539 | rs11100203 | A | 0.61 | 1.03 | 1.02-1.04 | $3.45 \times 10^{-9}$ |
| 28 | CEP120/SNX2 | 5 | 122409064 | 123178965 | rs752200 | A | 0.30 | 0.96 | 0.95-0.98 | $1.23 \times 10^{-10}$ |
| 32 | DOK3 | 5 | 176732662 | 177238487 | rs28510818 | T | 0.13 | 1.07 | 1.04-1.09 | $1.85 \times 10^{-8}$ |
| 37 | RUNX2 | 6 | 45157654 | 45657654 | rs2677109 | A | 0.34 | 0.97 | 0.96-0.98 | $4.06 \times 10^{-8}$ |
| 39 | FAM135A/C6orf57 | 6 | 70847590 | 71347590 | rs4707802 | T | 0.22 | 1.04 | 1.02-1.05 | $1.59 \times 10^{-8}$ |
| 40 | RGS17 | 6 | 153114643 | 153642908 | rs7761831 | T | 0.33 | 0.97 | 0.96-0.98 | $1.15 \times 10^{-8}$ |
| 41 | MAFK | 7 | 1314810 | 1840443 | rs3808335 | A | 0.43 | 0.97 | 0.96-0.98 | $3.08 \times 10^{-8}$ |
| 42 | PMS2/EIF2AK1 | 7 | 5816461 | 6320856 | rs66482438 | C | 0.22 | 1.04 | 1.02-1.05 | $4.21 \times 10^{-9}$ |
| 45 | HDAC9 | 7 | 17478001 | 18248418 | rs10282707 | T | 0.40 | 0.97 | 0.96-0.98 | $8.39 \times 10^{-11}$ |
| 50 | DOCK4 | 7 | 111330166 | 111830166 | rs144867634 | T | 0.98 | 1.1 | 1.07-1.14 | $3.48 \times 10^{-8}$ |
| 55 | RRM2B/ODF1/... | 8 | 103325949 | 103829609 | rs2436888 | C | 0.73 | 0.97 | 0.96-0.98 | $2.48 \times 10^{-8}$ |
| 57 | SLA | 8 | 133814152 | 134339676 | rs1533910 | T | 0.21 | 1.04 | 1.02-1.05 | $5.23 \times 10^{-9}$ |
| 59 | IFNE/IFNA8 | 9 | 21146216 | 21737443 | rs10964985 | T | 0.071 | 0.94 | 0.93-0.96 | $1.18 \times 10^{-8}$ |
| 60 | GNAQ | 9 | 80282374 | 80968995 | rs7038083 | A | 0.52 | 0.97 | 0.96-0.98 | $2.31 \times 10^{-8}$ |
| 61 | SYK | 9 | 93309029 | 93813884 | rs34221447 | T | 0.22 | 0.96 | 0.95-0.98 | $2.41 \times 10^{-9}$ |
| 62 | FGD3/IPPK/... | 9 | 95376730 | 95876730 | rs1022834 | A | 0.65 | 1.03 | 1.02-1.04 | $4.55 \times 10^{-8}$ |
| 68* | GPAM | 10 | 113213315 | 113713315 | rs79672230 | A | 0.20 | 1.04 | 1.02-1.05 | $4.54 \times 10^{-8}$ |
| 72 | CFL1/EFEMP2/... | 11 | 65331070 | 65912985 | rs77779142 | T | 0.16 | 0.96 | 0.94-0.97 | $1.60 \times 10^{-10}$ |
| 75 | NINJ2 | 12 | 651559 | 1199141 | rs10774462 | A | 0.68 | 1.03 | 1.02-1.05 | $2.12 \times 10^{-8}$ |
| 76 | GLS2/ANKRD52 | 12 | 56610020 | 57116962 | rs2657879 | A | 0.82 | 0.96 | 0.95-0.97 | $2.44 \times 10^{-10}$ |
| 78 | HIP1R/VPS37B/... | 12 | 123059475 | 123600204 | rs2061098 | T | 0.15 | 0.96 | 0.94-0.97 | $1.76 \times 10^{-8}$ |
| 79 | ATP6V0A2/TCTN2 /... | 12 | 123888346 | 124496004 | rs7302788 | T | 0.63 | 0.97 | 0.96-0.98 | $2.39 \times 10^{-9}$ |
| 80* | MCF2L | 13 | 113422477 | 113922477 | rs57258740 | T | 0.18 | 1.04 | 1.03-1.05 | $4.89 \times 10^{-8}$ |
| 85 | RASGRP1 | 15 | 38584033 | 39100330 | rs7173565 | T | 0.70 | 0.97 | 0.96-0.98 | $1.22 \times 10^{-9}$ |
| 86 | PLCB2 | 15 | 40334164 | 40839460 | rs2305650 | C | 0.76 | 0.97 | 0.96-0.98 | $4.48 \times 10^{-8}$ |
| 90* | GLCE | 15 | 69323077 | 69823077 | rs80343442 | T | 0.97 | 0.91 | 0.88-0.94 | $3.56 \times 10^{-8}$ |
| 92* | MCTP2 | 15 | 94552545 | 95052545 | rs6497185 | A | 0.73 | 0.97 | 0.95-0.98 | $3.92 \times 10^{-8}$ |
| 94 | GGA2 | 16 | 23153510 | 23898353 | rs2106454 | A | 0.77 | 1.04 | 1.03-1.05 | $1.06 \times 10^{-9}$ |
| 98 | PMFBP1 | 16 | 71903125 | 72457315 | rs72791115 | T | 0.82 | 1.04 | 1.03-1.06 | $1.15 \times 10^{-8}$ |
| 101 | C16orf95 | 16 | 86972153 | 87508088 | rs4843559 | A | 0.58 | 1.04 | 1.03-1.05 | $7.02 \times 10^{-12}$ |
| 104* | MNT/SGSM2/... | 17 | 2052387 | 2552387 | rs1381247 | T | 0.64 | 1.03 | 1.02-1.05 | $2.92 \times 10^{-8}$ |
| 113* | AC104532.2/VMAC | 19 | 5658969 | 6158969 | rs10404195 | A | 0.14 | 0.95 | 0.94-0.97 | $2.95 \times 10^{-8}$ |
| 114 | DNM2/KEAP1 | 19 | 10587677 | 11103296 | rs892086 | A | 0.58 | 1.03 | 1.02-1.04 | $1.55 \times 10^{-8}$ |
| 115 | LRRC25/SSBP4/ ... | 19 | 18260925 | 18862257 | rs1985157 | T | 0.58 | 0.97 | 0.96-0.98 | $1.11 \times 10^{-10}$ |
| 116 | YJEFN3/GATAD2A /... | 19 | 19124061 | 19907500 | rs2163804 | A | 0.18 | 1.04 | 1.03-1.06 | $6.90 \times 10^{-11}$ |
| 117 | CEP89/CEBPA | 19 | 33010762 | 33754259 | rs11665689 | A | 0.35 | 0.95 | 0.94-0.96 | $7.41 \times 10^{-18}$ |
| 118 | AXL | 19 | 41484560 | 41990895 | rs12979764 | T | 0.54 | 1.03 | 1.02-1.04 | $7.47 \times 10^{-9}$ |
| 124 | SMOX | 20 | 3889757 | 4397075 | rs13040038 | A | 0.44 | 0.97 | 0.96-0.98 | $7.69 \times 10^{-9}$ |
| 125 | UBE2V1/SPATA2 | 20 | 48328385 | 48840965 | rs113558364 | A | 0.72 | 0.96 | 0.95-0.98 | $7.47 \times 10^{-9}$ |

### New genes highlight druggable targets, pathways, and pleiotropy

Among the 48 new loci identified, we found 17 genes that interacted with non-AD approved drugs (see **Methods** and **Supplementary Data 11**). Five of these genes have been evaluated for their potential as drug targets for AD in previous research, three showed promise in animal models (*RRM2B*^15^, *SYK*^16,17^, and *KEAP1*^18^), while drugs targeting the other two genes (*CACNA1S* and *AXL*) were unsuccessful in improving AD symptoms in humans^19^ and mice^1920^ (see **Supplementary Note 6**).

Among the known loci, there were ten instances where the gene predicted by FLAMES differed from the gene reported in previous literature (see **Supplementary Note 5**). One of these genes (*QPCT*) is a potential drug target and was selected as an effector gene in locus 8 instead of two genes listed in previous GWAS (*PRKD3* and *NDUFAF7*). *QPCT* catalyzes the formation of pyroglutamate on position 3 of Aβ (AβpE3-X)^21^, which is the specific modification of Aβ targeted by an approved anti-amyloid antibody drug (donanemab); *QPCT* inhibitors are in development for several other neurodegenerative disorders as well^22^. Overall, multiple new genes, as well as newly prioritized genes in known loci, represent potential targets for drug development.

We were interested in how the new genes we identified fit into known AD-affected pathways. We found that five new genes (*SNX2, DOK2, SYK, GGA2*, and *CEBPA*) interacted with other known AD genes across two pathways (amyloid processing and microglia response to Aβ). One of the newly identified genes (*SNX2*) encodes an endosomal membrane protein in a family of genes (sorting nexins) that have been suggested to interact with *APP* or *APP* cleaving enzymes to regulate Aβ degradation^23^. Another (*GGA2*) affects Aβ processing through intracellular trafficking. *GGA2* influences the distribution of a key enzyme for *APP* processing (*BACE1*) in the Golgi apparatus, which then influences *APP* secretion^24^. Moreover, *DOK3* and *SYK* interact with each other^25^ and *TREM2*^17,25,26^ to potentially mediate microglial response to Aβ. *CEBPA* is another new gene that interacts with *TREM2* to promote *CD36* expression in microglia and may influence Aβ clearance^27^. Collectively, these findings reinforce the central importance of amyloid and microglial pathways, demonstrating how newly discovered risk factors converge upon these key disease mechanisms.

Across the 48 new loci identified, 5 have been associated with endophenotypes of neurodegeneration or another neurodegenerative disease (*HDAC9, C16orf95, HIP1R, MBNL1*, and *UBE2V1/SPATA2*). The *HDAC9* locus has been previously associated with neurofibrillary tangles and cerebral amyloid angiopathy^28^. *HDAC9* encodes a histone deacetylase, a family of genes associated with multiple diseases and neurodegeneration^29^. *C16orf95* has been previously associated with phosphorylated tau (pTau) levels in cerebrospinal fluid (CSF)^30^, but the function of *C16orf95* is not well characterized. While this locus harbors genome-wide significant SNPs in both AD and pTau, the local genetic correlation was not significant between them (*r*_*g*_ = -0.31, *p* = 0.12; see **Supplementary Data 12**), which suggests there may be independent mechanisms of association. The *HIP1R* and *MBNL1* loci have been previously associated with Parkinson’s disease (PD)^31^, but colocalization analysis suggested that these loci are unlikely to be driven by shared causal variants across AD and PD (**Supplementary Data 12**). The *UBE2V1/SPATA2* locus has been previously identified as a shared locus between amyotrophic lateral sclerosis (ALS) and frontotemporal dementia (FTD)^32^. We identified a nominally significant positive local genetic correlation between AD and ALS at this locus (*r*_*g*_ = 0.47, *p* = 0.006; see **Supplementary Data 12**). Excluding cohorts with AD definitions based on ICD code F03 (unspecified dementia) did not change this result (see **Supplementary Note 7**). Colocalization analysis indicated that this locus was driven by a shared causal variant (PP.H4=0.95) with the lead SNP (rs113558364) being the most likely shared variant (**Supplementary Note 8**). This suggests that this may be a shared locus for multiple neurodegenerative diseases. *UBE2V1* encodes a ubiquitin-conjugating enzyme and has been suggested to regulate protein aggregation^33,34^, and *SPATA2* is a part of a signaling network that regulates apoptosis^35^. Both of these processes are relevant to multiple neurodegenerative diseases, and there is evidence in this study for both of these genes as the potential effector gene (**Supplementary Note 8**).

Among known loci, we identified that the *GBA1* locus was shared between AD and PD, and the *TMEM163* locus had opposite effects driven by distinct variants in AD and PD, and likely mediated through different cell types (**Supplementary Note 8**). Overall, this analysis highlights 7 loci (*HDAC9, C16orf95, HIP1R, MBNL1, UBE2V1/SPATA2, GBA1*, and *TMEM163*) that may be involved in fundamental processes leading to neurodegeneration.

### Gene-set analysis highlights lipid binding, immune activation, amyloid catabolism, and cell-cell adhesion

We performed gene set enrichment analyses using MAGMA^36^ to highlight biological mechanisms relevant to AD. We identified 72 significant MSigDB and SynGO gene sets at a false discovery rate of 5% (Benjamini-Hochberg) (see **Supplementary Note 9** and **Supplementary Data 13**). After clustering these gene sets (see **Methods**), 13 clusters of more than one gene set were identified. These clusters reflected 4 biological processes identified in previous GWAS studies^4,5,37^: lipid binding, immune activation, amyloid catabolism, and cell adhesion. The immune activation and amyloid catabolism gene sets were still significant when we repeated the analysis using a meta-analysis of cohorts not included in the previous meta-analyses (**Supplementary Note 9** and **Supplementary Data 13**). In addition to the clusters, 21 gene sets were independent from any other gene set. These gene sets highlighted 5 biological processes that have been connected to AD pathology but not through GWAS gene set analysis: EphrinB reverse signaling^38^, glutathione metabolism^39^, RAS signaling^40^, sphingolipid metabolism^41^, and synapse endocytosis^42^. EphrinB has been suggested to influence amyloid beta toxicity^38^, glutathione metabolism influences oxidative stress which has been linked to AD^39^, RAS signaling regulates blood pressure and fluid balance and has been linked to multiple aspects of AD^40^, sphingolipid metabolism has been linked to AD through regulation of cell death^41^, and synapse endocytosis has been shown to be impaired in AD^42^. Overall, we highlighted multiple pathways that influence AD development through a range of biological mechanisms.

### Cell type analysis identifies microglia and neurons

We aimed to identify cell types relevant to AD by examining the association between cell-type-specific gene expression and GWAS signal. Using gene expression data in healthy controls, we identified a significant positive association between overexpression in microglia and the GWAS gene association (see **Supplementary Data 14** and **Supplementary Figure 11**). This suggests that these genes are preferentially active in microglia compared to other brain cell types, implicating the role of microglia in AD. This finding was in line with our previous GWAS of AD^4^. The association was robust to the removal of chromosome 19 and the proxy phenotype data (see **Supplementary Data 14**). Based on this finding, we also performed gene-set analysis to identify specific microglial states associated with the AD GWAS signal. While no state was significantly associated with AD, inflammatory states showed the strongest association (see **Supplementary Note 10, Supplementary Figure 11**, and **Supplementary Data 15**).

Next, we used cell-type-specific differential gene expression between AD cases and controls^43^ for a MAGMA gene property analysis. We tested whether there was an association between the GWAS gene association and differential expression in a cell type. We identified a significant association between differential upregulation of genes in microglia and AD GWAS signal. We also identified a significant association with differential downregulation of genes in 2 types of GABAergic neurons (small neuropeptide-expressing neurons (Sncg) and somatostatin-expressing inhibitory interneurons (Sst)) and 1 type of glutamatergic neuron (layer 6 intratelencephalic projection neurons (L6 IT Car3)) (see **Figure 3** and **Supplementary Data 16**). The microglia and neuronal associations were robust to the exclusion of chromosome 19 and proxy data (see **Supplementary Data 16**). After correction for the average differential gene expression across all cell types, the association of microglia remained significant (p = 1.58×10^-6^), while the neuronal associations were rendered only nominally significant (p<0.05) (see **Supplementary Data 16**). This suggests that the associations of sncg, sst, and L6 IT Car3 neurons may not be specific and could instead represent a general association with downregulated genes in neurons. Together, these results support the known involvement of microglia in AD. Additionally, we found an enrichment of AD GWAS signal in genes under expressed in neuronal cell types of AD cases compared to controls.

**Figure 3.**
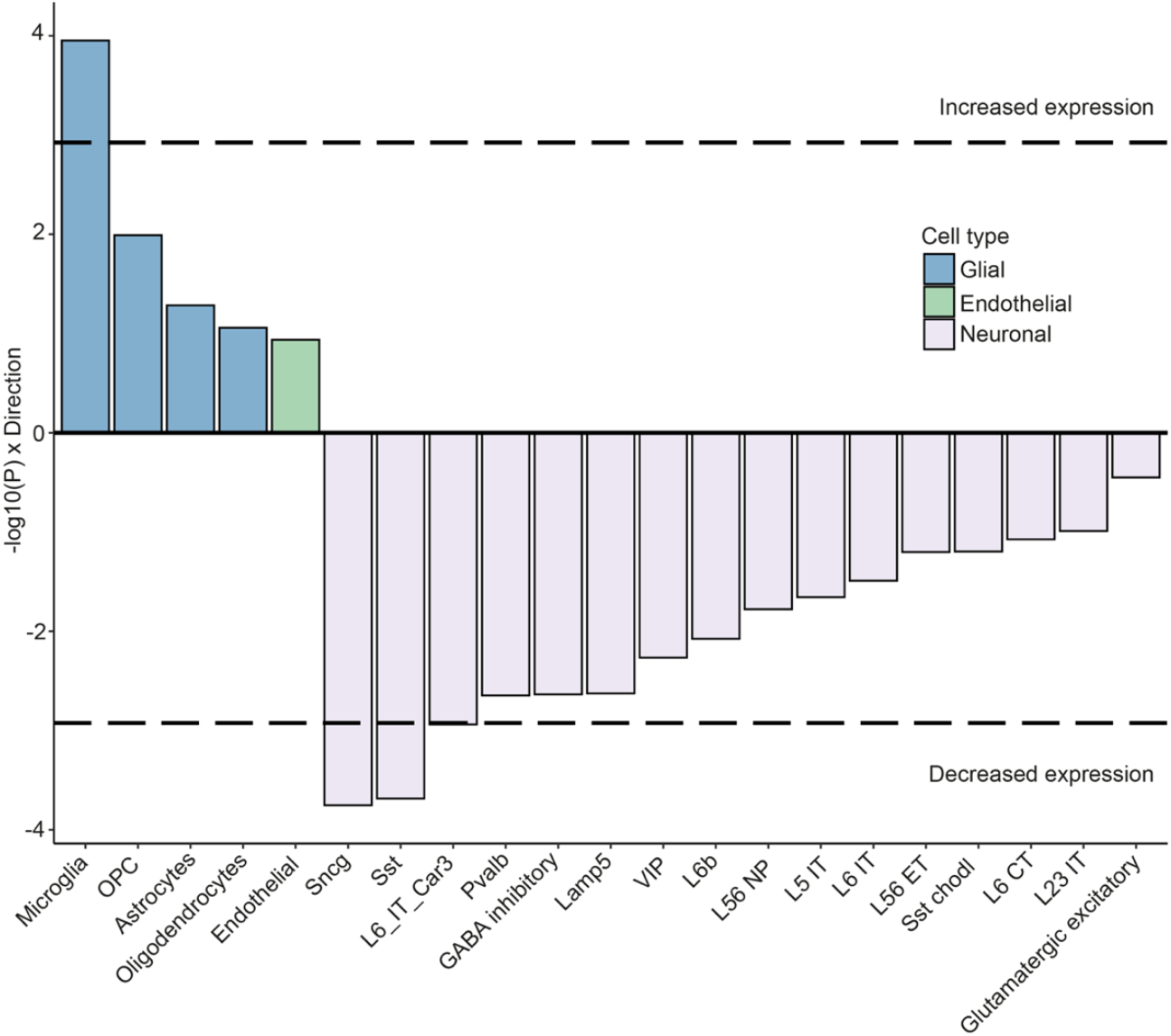
Cell-type enrichment analysis of differential gene expression data. Genes that are upregulated in AD microglia and downregulated in multiple neuronal subtypes in AD are enriched for AD GWAS signal. The dashed line marks the Bonferroni-corrected significance threshold for 42 tests (21 cell types × 2 directions; p < 0.05/42). For each cell type, only the more significant direction of effect (up- or downregulation) is displayed.

### Gene prioritization through specific cell types highlights microglial phagocytosis, astrocyte activation, myelination, and neuronal repair

To aid translation of the GWAS results, we sought to prioritize genes that influence AD through specific biological processes and cell types. We used the colocalization, MR, and cell type enrichment analyses to link genes to individual cell types, then drew on experimental literature to connect these genes to biological processes (**Supplementary Note 11**). Colocalization and MR analyses indicated that increased astrocytic expression of *KIF21B* may elevate AD risk. *KIF21B* upregulation has previously been linked to astrocyte activation and AD^44^, and aberrant astrocyte activation has in turn been associated with increased Aβ production and release^45^. This suggests that overexpression of *KIF21B* may influence AD through abnormal astrocyte activation and increased Aβ pathology. The same analysis indicated that decreased oligodendrocytic expression of *MAF* may also be linked to AD risk. Knockout of *MAF* has been associated with decreased myelination^46^, which, in turn, has been linked to AD^47^. As such, we speculate that decreased *MAF* expression reduces myelin production, rendering neurons more susceptible to AD pathology.

Through the colocalization, MR, and cell type enrichment analyses we identified multiple genes linked to microglia expression (**Supplementary Note 11**), 8 of which are relevant in microglia phagocytosis (*INPP5D, SYK, BLNK, HAVCR2, PICALM, AXL, FCER1G*, and *PLCG2*)^47–51^. We also prioritized genes that contributed to the differential cell type enrichment of neurons, three of which (*ZKSCAN1, APH1B*, and *SPRED2*) were nominally under expressed in neurons and significantly associated with AD. *APH1B* encodes a part of the γ-secretase complex that cleaves *App*^52^ and may contribute to AD through aberrant *App* processing in neurons. Decreased *SPRED2* expression has been observed after brain injury in a zebrafish model, where it has been implicated in neuro-regeneration^53^. While adult human neurogenesis remains debated, recent work has linked altered adult hippocampal neurogenesis to cognitive decline^54^. Therefore, we hypothesize that *APH1B* increases amyloid load, driving AD pathology, and that *SPRED2* may impair the neuronal response to neurodegeneration. In summary, by combining GWAS associations with expression data, we prioritized four biological processes acting through specific cell types and mediated by a small set of genes, providing targets for future experimental models.

### Polygenic Scores Capture a Large Proportion of SNP-Based Heritability

We used the meta-analysis results to estimate 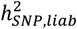 and improve polygenic prediction. We used SBayesRC^55^ with the EUR meta-analysis (excluding proxy cases) and estimated 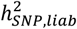 at 16% (s.e = 0.2%) (see **Figure 4a** and **Supplementary Data 17**). The 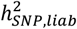 estimate for the proxy-case phenotype was nearly identical. The estimates for the EAS and AFR meta-analyses were 13% (s.e. = 4%) and 10% (s.e. = 6%), respectively (see **Supplementary Data 17**). The APOE-ε4 tagging variant rs429358 alone contributed 6% in EUR and EAS, but only 2% in AFR. These empirical results for *APOE* in EUR closely match theoretical derivations based on the liability threshold model (**Supplementary Note 12**). These estimates were based on a commonly used population prevalence of 5%; When using a population prevalence of 20% (e.g., in individuals older than 84^1^) the 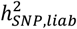 increased to approximately 25% in EUR (see **Supplementary Figure 12**). While the heritability point estimates differ between the populations for both LDSC and SBayesRC, the error bars in AFR and EAS contain the point estimates for all three populations, preventing conclusions about statistical differences between populations. Both LDSC and SBayesRC heritability estimates for AFR were not significantly different from zero, likely because of reduced power from smaller AFR sample sizes. Despite the larger AFR sample size relative to EAS, AFR populations showed larger standard errors under both methods, likely reflecting the greater genetic diversity and shorter LD blocks characteristic of African-ancestry populations, which impose a more complex correlation structure that existing LD reference panels may incompletely capture. Previous EUR GWAS of AD estimated the 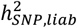 at approximately 5%^4,56^ using LDSC^57^ and a population prevalence of 5%. Using the EUR case-control meta-analysis, we arrive at a similar estimate of 6% when using LDSC (*s*.*e*. = 1%; see **Figure 4a**).

**Figure 4.**
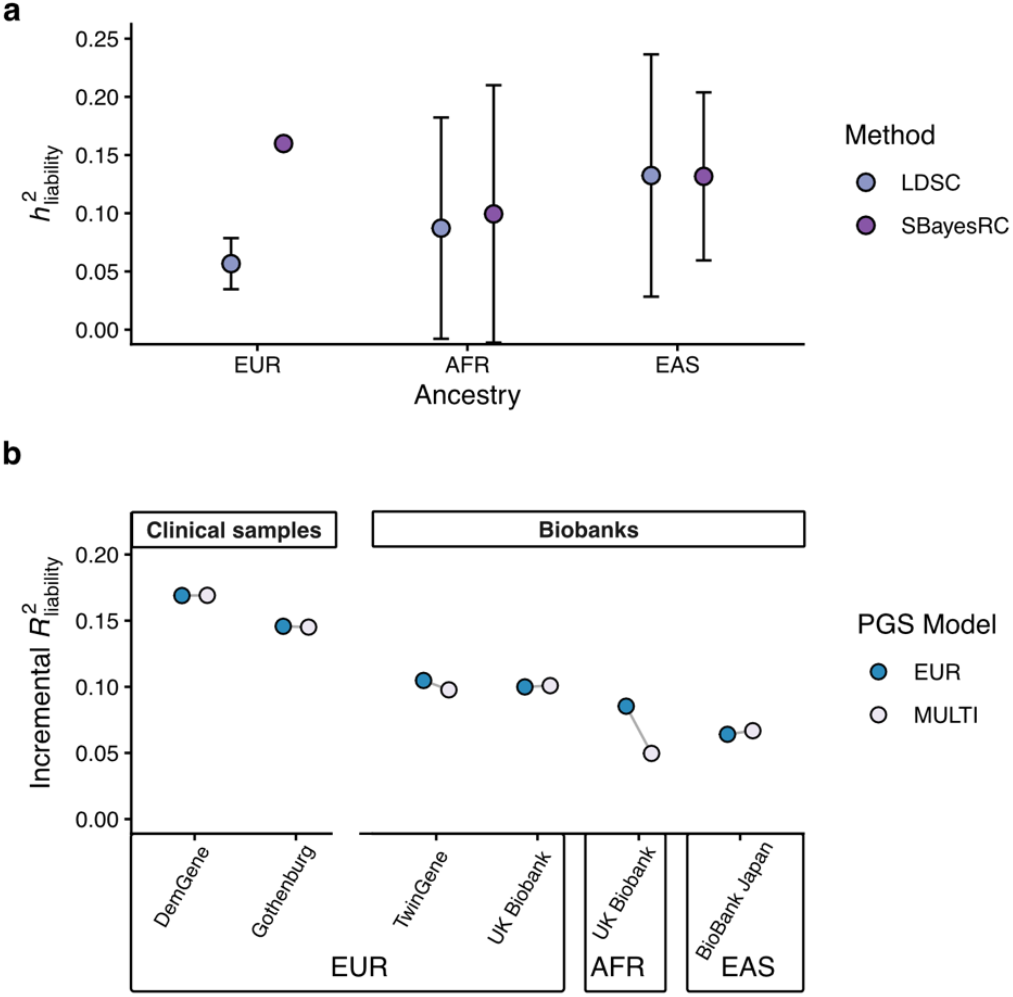
Heritability and polygenic prediction of AD. **a**, Summary-statistics based analysis of heritability of AD on the liability scale 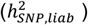 using LD Score Regression and SBayesRC. The error bars represent the 95% confidence intervals. **b**, Polygenic prediction performance in held-out clinical and biobank samples (excluding proxy cases). The base model included the first 10 principal components, and the model to be evaluated additionally included the PGS. We computed the incremental 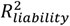 as the difference between the models. Two EUR clinical samples (DemGene and Gothenburg H70 Birth Cohort Studies and Clinical AD from Sweden) and two EUR biobank samples (TwinGene and UK Biobank) were available for prediction. One sample (UK Biobank) was available for prediction in AFR ancestry, and one (BioBank Japan) in EAS ancestry. We applied a leave-one-cohort-out approach, in which the PGS for each testing cohort was constructed using GWAS summary statistics that excluded that cohort.

The difference in 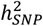 estimates between SBayesRC and LDSC was further investigated using simulations based on UK Biobank data, mimicking the genetic architecture of Alzheimer’s disease (**Methods**). SBayesRC without annotations (as no annotation effects were simulated) showed a small upward bias when fitting all SNPs as random effects, but produced unbiased estimates when excluding SNPs in LD with the APOE-ε4 variant (see **Supplementary Data 18**). In contrast, LDSC estimates were substantial downward biased, consistent with the result from the real data analysis. Based on the simulation findings, we conducted real data analysis (as reported above) using the same strategy, that is, fitting the APOE-ε4 variant as a fixed effect and all other SNPs outside the APOE LD block as random effects in SBayesRC. Based on these results and previous analyses^55,58,59^, we conclude that LDSC severely underestimates the 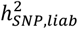 for AD.

We compared the predictive performance of a multi-ancestry (EUR, AFR, and EAS) PGS (see **Methods**) to that of a EUR-only PGS. We evaluated the PGS in hold-out samples of EUR, AFR, and EAS ancestry, as well as in clinical samples and biobanks (excluding proxy cases; see **Methods**). We excluded the AMR and SAS ancestries due to admixture, the unavailability of LD reference panels, and small sample sizes. The multi-PGS achieved a maximum incremental 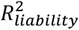 of 16.9% with an average of 12.8% in the EUR cohorts. The prediction in the two clinical samples (14.5% and 16.9%) was higher than in the two biobank samples (9.8% and 10.1%) (see **Figure 4b** and **Supplementary Data 19**). A PGS based only on EUR proxy cases achieved an average 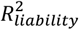 of 9.6% (see **Supplementary Figure 13**). We note the 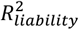 values represent point estimates without empirical standard errors.

The multi-ancestry PGS performed similarly to the EUR PGS in most cohorts, except in the AFR sample of the UK Biobank. This is likely due to heterogeneous APOE effect sizes across ancestries (see **Supplementary Figure 9**). Because the APOE effect size is smallest in AFR, the inclusion of PGSs based on ancestries with larger APOE effects, particularly EAS, negatively affects prediction in AFR. However, we also note that the AFR sample of the UK Biobank was particularly small (N = 550, see **Supplementary Data 20**).

As expected, both PGS performed worse in non-EUR cohorts. We note that the majority of the samples in the multi-PGS were of EUR ancestry. Furthermore, we found that the SBayesRC-based PGS is a better predictor than the combined *APOE*-e2 and *APOE-ε4* counts for all EUR cohorts (see **Methods** and **Supplementary Figure 14**).

When applying the PGS (UKB excluded) to unrelated individuals 65 and older of EUR ancestry in the UK Biobank, we found a 5.5 fold increase in case prevalence in the top 10% PGS group compared to the bottom 10% and a 3 fold increase when comparing the top 10% to the case prevalence of the whole sample **(Supplementary Figure 15**; **Supplementary Data 19**). This suggests that individuals with high PGS are at considerably higher risk of developing AD compared to the general population.

In conclusion, when compared to previous studies, we obtained higher estimates of 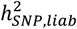 and demonstrated that a PGS derived from our meta-analysis explains a substantial and larger proportion of this heritability. Notably, the 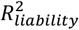 values based on SBayesRC^55^ exceeded the 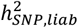 estimates obtained from LDSC, reinforcing that LDSC underestimates 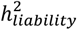 for AD.

## Discussion

We conducted the largest common variant analysis of AD to date and identified 127 independent loci, of which 48 were new. We applied multiple methodologies to prioritize genes in each locus, many of which were linked to pathways previously implicated in AD, most notably immune processes, microglia function, and amyloid catabolism.

Our cell-type analyses using RNAseq data showed enrichment in microglia, a well-established finding^4,5,60^. However, we also found that genes overexpressed in microglia and genes underexpressed in neurons in cases compared to controls were enriched for GWAS signal, expanding earlier work^4^. This suggests that AD risk variants may lead to overexpression of microglia genes and underexpression of neuronal genes. This is supported by previous transcriptomic and epigenomic studies that identified downregulation of neuronal functions and upregulation of innate immune response as relevant to AD^61,62^. Of the three types of neurons significantly underexpressing AD genes in cases (Sncg, Sst, and L6 IT Car3), Sncg and Sst neurons have been previously shown to be vulnerable early on in the AD disease process^63,64^. This suggests that the variants associated with AD in this GWAS may influence gene expression in vulnerable neuronal subtypes leading to neuronal cell death. Among the genes prioritized for the neuronal enrichment signal, we identified one linked to *App* processing (*APH1B*) and another implicated in post-brain-injury repair in zebrafish (*SPRED2*). These findings alone have limited ability to determine whether the neuronal enrichment reflects a primary or secondary disease process.

AD remains a challenging disease to treat. While new disease-modifying drugs have recently been approved, their effectiveness remains a subject of dispute^7,8^, and new avenues need to be explored that go beyond amyloid clearance. Here, we highlighted five potential drug targets among the new genes we identified. Two of these (*SYK*, and *AXL*) are involved in the immune response to Aβ. Existing treatments targeting these genes have shown promise in AD mouse models^16,20,65,66^. We further suggested that *AXL* and *SYK* along with the target of *SYK* (*BLNK*) were linked to AD through microglia. These genes, along with other known AD genes (*INPP5D, FCER1G*, and *PLCG2*)^48^ and a new gene prioritized in this study (*CEBPA)*^27^, have been suggested to play a role in microglial phagocytosis. Further research into these genes and microglial phagocytosis as drug targets for treating the microglial component of AD disease progression may identify them as complementary treatments to drugs targeting Aβ. Additionally, we identified that *UBE2V1* and *SPATA2* may be relevant to multiple neurodegenerative diseases. Further research targeting these genes may help address core mechanisms of neurodegeneration and provide therapeutic benefit across multiple conditions.

The SBayesRC estimate of 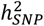 (16%) in EUR is substantially higher than the LDSC estimate (6%). The LDSC estimate was downward biased, as the APOE-ε4 variant alone explains 6% of the variance on the liability scale as shown in theory and empirical analyses, consistent with our simulation results. When running a standard SBayesRC model that fits all SNPs jointly, we observed a 2% inflation in 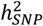 estimation in simulations and an estimate of 18% in the real data analysis. This upward bias is likely attributable to the use of logistic regression for GWAS, where the correlations between the estimated effects of the APOE-ε4 variant, which has a very large effect, and neighbouring SNPs in LD is not exactly proportional to their LD correlations. After removing SNPs within the same LD block as the APOE-ε4 variant, the SBayesRC estimates were unbiased in simulations and 2% lower in the real data analysis. Our approach therefore provides a conservative estimate of 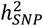, under the assumption that there is no additional genetic variation in LD with APOE. Our PGS prediction achieved an average incremental 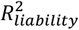 of 13%, exceeding the LDSC 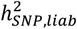 estimate (which is the theoretical upper limit of the 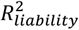) and further underscores that LDSC substantially underestimates 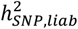 in AD.

There are several methodological differences between SBayesRC and LDSC that may explain the observed differences. SBayesRC explicitly models a mixture of SNP effect size distributions, accommodating variants with large effects, whereas LDSC’s regression framework implicitly assumes a highly polygenic architecture with normally distributed effect sizes^57,58,67^. This distinction is particularly consequential for AD, which exhibits a more oligogenic architecture due to the large-effect *APOE* locus. We expect this to be the principal methodological difference underlying the higher heritability estimates from SBayesRC. Additionally, unlike LDSC, SBayesRC incorporates functional annotations to inform variant prioritization and effect size distributions.

Our study marks the largest multi-ancestry genomic study of AD to date, encompassing 2.8 million individuals including 8673 cases, 15005 proxy cases, 105,525 controls and 116,426 proxy controls of non-EUR ancestry. Nonetheless, the proportion of non-EUR samples was still relatively low, which made the findings of ancestry-specific effects and the PGS prediction performance in non-EUR samples less precise. Future studies including more individuals of non-EUR ancestry are essential to improve equity in AD genomic research. To accelerate this process, we have made the summary statistics from our ancestry-specific meta-analyses publicly available, as well as sex- and phenotype-stratified (case-control vs. proxy) summary statistics, X chromosome summary statistics, and data excluding the UK Biobank and cohorts included in previous GWAS.

Our study included 74,141 proxy cases (40% of total cases) with the goal of boosting power, which yielded 32 additional genome-wide significant loci. The use of proxy cases is particularly valuable for AD, given its late-onset nature: biobanks predominantly recruit middle-aged participants who are often too young to have developed AD themselves but can report parental diagnoses through family history questionnaires. Genotyped individuals with an affected parent can be included as proxy cases, since they carry half of the parental genotype and therefore a portion of the risk variants. While a recent study suggests that including such AD proxy cases can introduce some statistical bias, this appears to be limited to genetic correlation analyses, particularly for educational attainment, with no observed impact on locus discovery, which is our primary aim. The proportion of proxy cases varied by ancestry: 37% in EUR samples, 63% in AFR, 62% in EAS, 68% in AMR, and 82% in SAS. This disparity reflects differential data accessibility: Non-EUR samples derived primarily from national biobanks with family history data and liberal data access policies, whereas established clinical networks in Europe facilitated direct case ascertainment in EUR populations from hospitals and memory clinics.

In conclusion, we identified 127 AD risk loci that implicate several new genes, reveal new molecular and cellular mechanisms with potential for future drug development, and improve PGS prediction and heritability estimates.

## Methods

### Quality control of individual-level data

Newly participating cohorts received an analysis plan detailing quality control, principal component analysis, imputation, phenotype definitions, GWAS models, and results formats (see **Supplementary Methods**). Briefly, pre-imputation quality control followed RICOPILI^68^ guidelines, namely removal of SNPs with a call rate lower than 0.98, palindromic SNPs with minor allele frequency higher than 0.3, invariant and multi-allelic SNPs, SNPs with Hardy-Weinberg-Equilibrium p-value in controls smaller than 1 × 10^6^, and SNPs with Hardy-Weinberg-Equilibrium p-value in cases smaller than 1 × 10^10^. Individuals were removed if their call rate was smaller than 0.98, their inbreeding coefficient was outside of the range -0.2 to 0.2, or where the genetic sex was not concordant with the reported sex. We note that many cohorts performed quality control independently from this analysis plan. Detailed descriptions and divergences are provided in the **Supplementary Methods**.

### Quality Control of GWAS summary statistics

Every set of summary statistics underwent a standardized quality control protocol. Because the majority of summary statistics were on genomic build 37, we used this as the consensus build. All other summary statistics on build 38 were lifted to 37 using the snp_modifyBuild() function in the bigsnpr package^69^. snp_modifyBuild() does not account for strand changes between genomic builds, which we manually corrected. We removed SNPs with missing values, indels, duplicated, multiallelic or monomorphic SNPs, SNPs with minor allele frequency below 0.001, SNPs with nonsense values, palindromic SNPs with minor allele frequency larger than 0.4, SNPs with allele frequency differences larger than 0.2 with a reference sample, or SNPs with INFO scores below 0.6 for dosage and 0.8 for hardcall data.

### Case and control definitions

We applied the following AD definition for most cohorts; however, there was heterogeneity in how AD was ascertained across studies. Details on the specific definition used in each cohort are provided in the **Supplementary Methods**. AD cases were defined based on ICD-10 codes (G30, F00*, F03) or clinical diagnosis. When possible, to minimize the inclusion of familial (Mendelian) AD cases, individuals with age-of-onset < 40 were excluded. If age-of-onset data were unavailable, we used age-at-assessment, prioritizing the earliest visit at which the individual was identified as a case. Detailed age and sex information can be found in **Supplementary Data 21**. Controls (coded as 0) were defined as individuals with no diagnosis or self-reported history of AD, dementia, or mild cognitive impairment (ICD-10: G30, F00*, F03, F06.7), nor any other neurodegenerative disorders (ICD-10: G20–G26, G31–G32, G35– G37, G10–G14, F02). Parental diagnoses were also considered, and individuals with a parent affected by any of these conditions were excluded from the study. Wherever possible, we aimed to include at least four times as many controls as cases in each GWAS to maximize power. This strategy resulted in several cohorts including controls below the typical age of onset, which risks misclassifying future cases as controls and may attenuate statistical power. However, published simulation work demonstrates that, at a population prevalence of 5%, a large set of unscreened controls yields greater power than a smaller set of screened controls at half the sample size^70^.

### Proxy-case analysis

Individuals with first-degree relatives with AD will, on average, carry half of the alleles that affect the risk of AD. As such, genotyped individuals can be used as proxies for their affected first-degree relatives, even though they are not affected themselves. In the present study, a subject was designated a proxy case (coded as 1) if either their mother or father is reported to have AD / Dementia. If the genotyped individuals are themselves AD cases, we excluded them from this analysis. We ran the GWAS separately in individuals who reported their mothers versus their fathers to have AD, and meta-analyzed the resulting summary statistics^71^. If genotyped individuals reported both their parents to have AD, half were randomly assigned to the paternal, and the other half to the maternal GWAS. Resulting GWAS effect sizes and standard errors were multiplied by 2 to correct for proxy status and transform them to the same scale as the case-control GWAS^72^. We found the proxy GWAS effect sizes after transformation to be well calibrated (See **Supplementary Figure 16**).

### GWAS analysis

A GWAS was run if at least 100 cases or 200 proxy-cases were available for analysis. When several ancestries were available, a GWAS was run separately in each ancestry. In the main analysis, we used the following regression model correcting for sex, age, principal components (PC), and other study-specific covariates:

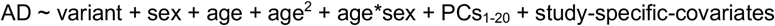

In the sex-stratified analyses, we use the following regression model:

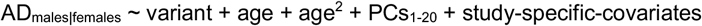

Not all cohorts had the same information available for every individual. As such, slightly different models were applied in some cohorts. Details for each cohort can be found in the **Supplementary Methods**.

### GWAS meta-analysis

We used metal (version 2020-05-05) to run all inverse-variance weighted fixed-effect meta-analyses. For the main analyses, we first ran ancestry- and phenotype-stratified (i.e., case-control vs. proxy) meta-analyses. Subsequently, we meta-analyzed all ancestries within the same phenotype definitions. Lastly, we meta-analyzed the case-control and proxy summary statistics within and across all ancestries. After having performed all meta-analyses on unfiltered summary statistics, the results were filtered for SNPs with at least 60% of the maximum effective sample size in a given set.

### Effective sample size

The effective sample size (*N*_*eff*_) is the size of a balanced case-control study (ratio 1:1) that would yield the same statistical power as the actual study with an imbalanced case-control ratio. It is calculated as the sum of *N*_*eff*_ for each cohort^10^, where 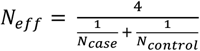. For samples with proxy-cases, *N*_*eff*_ was further divided by four^72^.

### FLAMES

We fine-mapped genome-wide significant loci with the PolyFUN (v1.0.0)^73^ implementation of SuSiE^74^. Because of the nature of large meta-analyses and the sharing restrictions present on raw genetic data, the generation of in-sample LD reference panels is infeasible. We therefore restricted fine-mapping to a single causal variant per locus. MAGMA^36^ scores were generated using the SNP-wise mean model with an ancestry-matched reference panel from the UKB and with a reference panel of all individuals included in the 1000 Genomes project (analysis implemented in FUMA v1.8.3). The UKB reference panels consisted of random sets of 10,000 individuals of each ancestry in the UK Biobank^75^. For ancestries with fewer individuals, all were used. For the multi-ancestry UKB reference panel, 10,000 individuals were selected such that the proportion of each ancestry matched that of the multi-ancestry GWAS. PoPS (v0.1)^76^ scores were generated for each ancestry, using their corresponding MAGMA scores as input. Credible set annotation and effector gene prediction were performed using FLAMES (v1.1.2)^12^. Credible sets were annotated with the MAGMA and PoPS scores matching the ancestry of the summary statistics from which the credible set was derived. Genes selected by FLAMES for each locus were reported from four FLAMES analyses: multi-ancestry summary statistics using 1000 Genomes and UKB reference panel, and EUR only summary statistics using the 1000 Genomes and UKB reference panel.

### Co-localization and Mendelian Randomization

To identify potential risk genes driven by GWAS signals in non-coding regions, we implemented a multi-faceted approach integrating molecular quantitative trait loci (QTL) colocalization analysis and Mendelian Randomization (MR)^77^. First, we defined the boundaries of each GWAS locus by identifying all SNPs in LD (r^2^ > 0.1) with the lead GWAS SNP. This LD analysis was based on the EUR population from the 1000 Genomes Project Phase 3, utilizing LDlink ^78^. Next, for each locus, we performed colocalization analysis to assess whether the GWAS signal shared a causal variant with molecular QTLs (eQTLs for gene expression, sQTLs for splicing, and pQTLs for protein abundance). We considered all genes for which QTL association data were available for the SNPs within the defined locus boundaries. This analysis was conducted using the coloc R package^13^ under the assumption of a single causal variant per signal. We leveraged a comprehensive set of QTL datasets from relevant human brain tissues or cell types^79–90^. Following colocalization, we performed two-sample Mendelian Randomization (MR) to infer a potential causal relationship between the gene’s expression/splicing/protein level and AD. For each gene that showed evidence of colocalization, the top associated eQTL/sQTL/pQTL from the respective study was used as the genetic instrument. A gene was considered a high-confidence candidate if it met three stringent criteria: 1) strong evidence of colocalization between the GWAS signal and the gene’s expression/splicing/protein level, defined as a posterior probability of a shared signal (PP.H4.abf) of 0.8 or greater; 2) a statistically significant MR result after correcting for multiple testing across all genes with PP.H4 ≥ 0.8 (False Discovery Rate < 0.05); and 3) the transcription start site (TSS) of the gene was located within 500kb of the lead GWAS SNP. We performed the same colocalization and MR analyses on the loci shared with ALS and PD using summary statistics from van Rheenen *et al*. (2021)^91^ and Nalls *et al*. (2019)^31^.

### Common and rare variant GWAS literature search

To identify which loci and genes have been reported in previous literature, we conducted a literature search of all GWAS of AD listed in the GWAS catalog, which yielded 206 studies (https://www.ebi.ac.uk/gwas/efotraits/MONDO_0004975; Accessed 7 October 2024). We then filtered these for common variant GWAS where AD was the primary phenotype, resulting in 49 studies (**Supplementary Data 3**). Next, we extracted all significant lead SNPs (*p* < 5×10^-8^) reported in these studies and the genes assigned to them. We defined 250kb windows around the lead SNPs and merged overlapping windows into single loci. This yielded a final set of 174 non-overlapping loci identified in previous AD GWAS (**Supplementary Data 3**). We were also interested in whether our common variant GWAS implicated the same genes as previous rare variant studies of AD. We created a list of rare variant studies of AD (largely based on two reviews^92,93^). We identified 32 studies that highlighted 25 genes (**Supplementary Data 4**).

### Drug–Gene Interaction Analysis

To investigate the potential translational relevance of genes prioritized in our Alzheimer’s disease (AD) genome-wide association study (GWAS), we performed a systematic drug–gene interaction analysis using the Drug–Gene Interaction Database (DGIdb)(ref=82), a curated compendium of interactions collated from multiple evidence sources. The input comprised prioritized effector genes derived from our genetic and post-GWAS annotation pipeline (see Gene Prioritization). These genes were queried against DGIdb to retrieve all reported drug–gene interaction pairs, yielding an initial set of 1,525 interactions (drug_gene_interactions_all.csv).

To focus on interactions of potential therapeutic significance, we applied the following criteria. First, we retained only drugs with documented approval by major regulatory agencies, specifically the U.S. Food and Drug Administration (FDA) or the European Medicines Agency (EMA); approval status was taken from DGIdb annotations and cross-referenced with authoritative regulatory records. Second, we filtered on DGIdb’s interaction confidence metric, retaining pairs with an interaction score ≥ 0.1 to exclude low-confidence associations while preserving interactions supported by at least minimal evidence. The final curated set comprised 220 approved drug–gene interactions (drug_gene_interactions_approved_SC0.1.csv), representing drugs with established regulatory status and evidence for interaction with prioritized AD-implicated genes.

For interpretative context, drugs in the curated set were assigned to broad mechanistic categories reflecting their primary mode of action or their relevance to biological processes implicated in AD (e.g., cholinergic signalling, amyloid homeostasis, lipid metabolism, vascular regulation, and neuroinflammation) based on documented mechanism-of-action information and literature review. Full interaction results and drug categorizations are provided in **Supplementary Data 11**.

### Polygenic prediction using SBayesRC

SBayesRC^55^ is a Bayesian hierarchical mixture model for polygenic prediction, jointly fitting genome-wide SNPs and incorporating various functional genomic annotations into the estimation of SNP effects. We applied SBayesRC to GWAS summary statistics of AD from different ancestry groups, along with LD matrices derived from reference samples of the same ancestry in the UK Biobank^75^, and 96 functional annotations from the BaselineLD model (v2.2)^94^. These annotations include evolutionary conservation, functional roles of DNA sequence (e.g., coding, promoter, and enhancers), and LD or MAF-related features. In SBayesRC, the annotations inform the probabilities that a SNP has zero, small, or large effects, with both annotation and SNP effects estimated using Markov chain Monte Carlo (MCMC). We ran SBayesRC using 4 independent MCMC chains, each with 5,000 iterations, discarding the first 3,000 as burn-in. The SNP effect estimates were obtained as posterior means across iterations and chains. Since SBayesRC accounts for LD between SNPs, all SNPs were included in the prediction equation. For cross-ancestry prediction, we first estimated SNP effects separately within each ancestry group, then linearly combined the ancestry-specific effect estimates using a tuning sample from the target ancestry and finally applied the combined SNP effects to the testing data (see **Supplementary Data 20** for the tuning and testing sample sizes).

We computed PGSs based on the SBayesRC posterior means using PLINK1.9 (version Linux 64-bit 6th June, 2021; command “--score <variant ID column> <effect allele column> <posterior mean beta> sum center”). To evaluate prediction, we used the coefficient of determination on the liability scale 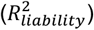. We converted the *R*^*2*^ from the observed scale to the liability scale based on the transformation introduced in Lee et al. (2012)^95^. The base model included the first 10 principal components, and the model to be evaluated additionally included the PGS. We computed the incremental 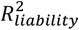 as the difference between the models. We note we did not include other covariates, such as age and sex, in the polygenic score analyses because doing so can lead to collider bias and reduce power in ascertained samples^96,97^ (the case ratio in the clinical samples was up to 43%).

We evaluated the PGSs in five cohorts: DemGene, Gothenburg H70 Birth Cohort Studies and Clinical AD from Sweden, TwinGene, UK Biobank, and BioBank Japan. DemGene and Gothenburg H70 Birth Cohort Studies and Clinical AD from Sweden are clinical samples, where AD status was ascertained in memory clinics in Norway and Sweden. TwinGene, UK Biobank, and BioBank Japan are biobanks with limited phenotypic appraisal. A sixth cohort available for prediction, STSA, was excluded because it did not clearly fall into either the clinical or biobank category, representing a mixture of both. We applied a leave-one-cohort-out approach, in which the PGS for each testing cohort was constructed using GWAS summary statistics that excluded that cohort. More details for each cohort can be found in the Supplementary Methods.

We compared the prediction of the PGS to APOE counts. APOE-ε4 dosage is fully determined by the count of the T allele at SNP 19:45411941:C:T (rs429358). Similarly, the number of T alleles at SNP 19:45412079:C:T (rs7412) determines the dosage of APOE-e2. We included both counts as predictors in a regression.

### SNP-based heritability estimation using SBayesRC

Besides SNP effects, SBayesRC provides estimates of genetic architecture parameters, including SNP-based heritability 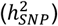. To estimate 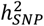 on the liability scale, we first converted GWAS marginal effects obtained from logistic regression (*b*_*logit*_) to the liability scale using *b*_*l*_ = *b*_*logit*_ × *K* × (1 − *K*)/*z*, where K is the population lifetime prevalence (set to 0.05), and z = dnorm(qnorm(K)). Given the allele frequency (*p*) in the LD reference samples, we estimated the sample size on the liability scale by 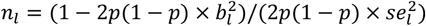 for each SNP. We then ran SBayesRC using *b*_*l*_, *n*_*l*_ and *p* as input. The SNP-based heritability 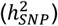 on the liability scale was calculated in each MCMC iteration as *β*′*Rβ*, where *β* is the vector of sampled SNP effects and R is the LD correlation matrix. As shown in the simulation study, a small inflation in the 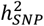 estimation could occur when including SNPs in LD with the APOE-ε4 tagging SNP (rs429358), probably due to a combination of the large effect of APOE-ε4 allele and the imperfect approximation of LD correlation to the correlation between logistic regression coefficient estimates. To avoid the potential inflation, we therefore fit rs429358 as a fixed effect and all other SNPs as random, excluding SNPs in the same LD block of rs429358 from the model. LD blocks were defined based on the quasi-independent LD blocks in the human genome^98^, merging small blocks to have a minimum size of 2cM. We estimated the total 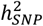, fixed-effect 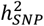, and random-effect 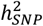. The posterior mean across MCMC iterations was reported as the estimate of 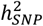, and the posterior standard error was calculated as the standard deviation of the MCMC samples. To assess convergence, we computed the potential scale reduction factor (PSRF)^99^ based on 4 independent chains.

### Simulation based on UK Biobank data

To investigate the difference in 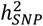 estimate between LDSC and SBayesRC, we simulated a phenotype whose genetic architecture mirrors that of Alzheimer’s disease and controls in 348,494 unrelated participants of the European ancestry in the UK Biobank with a disease prevalence of 5% (i.e. 17,424 cases and 331,070 controls) and an 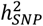 of 12% with the *APOE-ε4* tagging SNP (rs429358) accounting for 6% of the 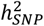 and another 1,000 SNPs randomly selected across the autosomes for the other 6% of the 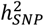. Under the additive model, individual liabilities were computed as the sum of allelic effects across all causal variants, plus Gaussian noise scaled to achieve the target heritability. The 5% of individuals with the highest liabilities were classified as cases. Simulations were replicated 5 times using the GCTA software version 1.95.1^100^. GWAS were performed on the simulated binary phenotype using logistic regression implemented in PLINK2^101^. We first converted GWAS marginal effects and standard errors from logistic regression and sample size to the liability scale using *b*_*l*_ = *b*_*logit*_ × (1 − *K*)/*z, se*_*l*_ *= se*_*logit*_ × *K* × (1 − *K)/z* and *n*_*l*_ = 348,494 × *z*^2^/ (*K* × (1 − *K*)) = 78,040 following Yang et al.^102^. Then, we ran SBayesR to estimate 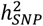 of the simulated phenotypes of the five replicates. We also ran SBayesR by considering the *APOE-ε4* tagging SNP (rs429358) as fixed effects and skipping SNPs with an LD r2 > 0.001 with rs429358 or skipping all variants in the same LD block containing rs429358 while taking all other variants as random effects. We used 10,000 unrelated individuals of European ancestry in the UK Biobank as an LD reference sample, same as in the empirical analysis. For LDSC, we first estimated 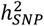 of the simulated pehnotypes using LD scores of the HapMap3 variants; since rs429358 is absent in the HapMap3 LD scores files, we also ran LDSC using the full LD scores (not restricted to the HapMap3 variants) prepared by the pan-UK Biobank project^103^ (https://pan-dev.ukbb.broadinstitute.org/).

### Global genetic correlation analysis using SBayesRC

We performed sex-stratified SBayesRC analyses and obtained posterior means of SNP effect sizes. During each MCMC iteration, we computed the correlation between these SNP effects in males and females, and estimated the posterior mean and standard deviation of the global genetic correlation as the mean and standard deviation of these correlations across all iterations. This approach may underestimate the true correlation because LD can lead to different but correlated SNPs being selected in males and females within a given iteration. Thus, the estimated correlation should be regarded as a lower bound of the true global genetic correlation.

### Local genetic correlation analysis

We aimed to use local genetic correlations with traits relevant for AD to help explain the associations of the new loci with AD. We first lifted over the locus boundaries of the new loci to build GRCh37 from GRCh38 using the UCSC liftover tool^104^. Next we queried the GWAS catalog^105^ (version e114_r2025-05-1) to find neurodegenerative phenotypes reported as having an association within the locus. We used the available summary statistics from EUR ancestry individuals for these traits for a local genetic correlation analysis with AD using LAVA^106^. For each summary statistics file, we removed SNPs with a sample size smaller than 60% of the maximum sample size. For dichotomous disease traits, we used the effective sample size (neff) as input in the summary statistic file, and defined the case and control sample sizes in the LAVA info files as 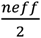. To account for potential sample overlap, we supplied the intercepts from cross-trait LDSC^57^ to LAVA. We used the European 1000 Genomes sample as an LD reference panel and defined the LAVA locus file based on the GWAS loci. We only computed local genetic correlations for locus pairs with sufficient genetic signal. To this end, we applied a 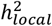 threshold of *p* < 0.05.

### Gene-set and Cell type analyses

We used MAGMA v1.10^36^ to perform gene-set enrichment, and cell type enrichment analyses. We performed these analyses using the gene-level associations from the FLAMES analysis using the UKB reference panel. As a sensitivity analyses, we also reported the P-value of all significant findings when using the gene level associations derived from using the 1000 Genomes data as a reference panel. The gene-level analysis results used for gene-set and cell type enrichment analyses were the same gene-level analysis results (snp-wise=mean) used for gene prioritization (FLAMES).

We performed gene-set enrichment analyses for all of the gene-sets included in MSigDB^107^ v2024.1.Hs C2 (curated), C5 GO (gene ontology), C7 (immunologic) gene-set groups, and the SynGO^108^ v20231201 gene-sets. The MSigDB gene-sets were downloaded as gene symbols and converted to Ensembl gene IDs using BioMart^109^. We excluded gene-sets with fewer than 10 genes; This resulted in a total of 18,782 tested gene-sets. We then selected gene-sets based on a false discovery rate of 5% using the Benjamini-Hochberg procedure. As a sensitivity analysis for the effect of *APOE*, we performed the gene-set analysis with the whole of chromosome 19 excluded. We performed pairwise conditional analysis with MAGMA to calculate how much of each association could be explained by the other gene-sets 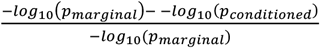.

This proportion of explained association was then used to cluster the gene-sets by edge betweenness (cluster_edge_betweenness in the R package igraph^110^). Only connections between gene-sets where more than 30% of the marginal association was explained by the other gene-set were included in the clustering. Clusters of gene-sets were assigned names based on the function of the gene-sets that make up the cluster.

We performed MAGMA gene property analysis using FUMA v1.7^111^ to highlight cell types relevant to AD. We tested whether the gene expression pattern of a cell type was significantly correlated with the association of the genes with AD. This regression model included the average gene expression for that tissue as a covariate. The association test was one-sided, only considering a positive relationship between gene expression and AD association. We tested this across 4 brain datasets: Siletti et al. (2023)^112^ hippocampus (datasets 59-69 level 2); Allen Human Brain Atlas^113^ (Allen_Human_MTG_level2); GSE168408 human prefrontal cortex^114^ (GSE168408_Human_Prefrontal_Cortex_level2_Adult); and PsychENCODE^115^ (PsychENCODE_Adult). We performed the full 3-step procedure outlined by FUMA to identify cell types associated with AD after conditioning on associations within and between datasets. We selected significant cell types in step 1 using Bonferroni correction for 337 cell type dataset pairs. As a sensitivity analysis for the effect of *APOE*, we performed gene property analysis excluding the whole of chromosome 19.

We also used MAGMA gene property analysis to highlight cell types where differentially expressed genes across AD cases and controls were enriched for AD GWAS signal. We extracted gene differential expression P-values per cell type from Nakatsuka et al. (2025)^43^ (SumRank up and down regulation from Supplementary File 3). We used the - −*log*_10_(*p*) of the differential up and down regulation analyses from each cell type as the gene property for the MAGMA gene property analysis. As with the FUMA cell type analysis, we tested whether more significant differential expression was associated with increased association in the AD GWAS. As a sensitivity analysis, we included the average association of each gene across all cell types as a covariate. Differential expression P-values of 0 were converted to 1×10^-10^ to avoid infinite values of –*log*_10_(*p*). The differential expression P-values were originally calculated with 10,000 permutations, so 1×10^-10^ was chosen as a P-value small enough to be beyond the calculation of the permutations but close to the boundary. Gene symbols were converted to Ensembl gene ID using BioMart^109^. As a sensitivity analysis for the effect of *APOE*, we performed the gene property analysis with the whole of chromosome 19 excluded.

We performed MAGMA gene-set enrichment analyses to determine if genes linked to specific microglia states were enriched for AD GWAS association. We extracted the gene-sets that were used to define the 12 microglia states from Sun et al. (2023)^116^ (extracted from **Supplementary Data 2**). Again, we removed chromosome 19 as a sensitivity analysis.

### METASOFT

To assess whether effect sizes were heterogeneous across ancestries, we applied METASOFT^11^ (version 2.0.1) to genome-wide significant loci identified in the primary meta-analysis. METASOFT is a random-effects meta-analysis framework specifically designed to detect and quantify heterogeneity in genetic associations across multiple studies or populations. Unlike fixed-effects approaches that assume a single true effect size shared across all studies, METASOFT’s RE2 (random-effects) model allows effect sizes to vary across populations and explicitly models this between-study heterogeneity using a random-effects variance component (τ^2^). The RE2 model estimates the probability that an effect exists in each individual study while simultaneously testing for overall association across all studies, making it particularly well-suited for identifying ancestry-specific or population-stratified genetic effects.

## Data availability

Summary statistics will be made available upon acceptance.

## Code availability

Analysis code can be downloaded from: https://github.com/euffelmann/paper-pgc_alzheimers

## Acknowledgements

This project was supported by the NWO Gravitation grant *BRAINSCAPES: a roadmap from neurogenetics to neurobiology* (grant no. 024.004.012), the European Research Council Advanced Grant (grant no. ERC-2018-ADG 834057), the Research Council of Norway (grants #344121, #324499, and #353629), the European Union’s Horizon 2020 Research and Innovation Action (grant #964874), NordForsk (grant #164218), and the NIMH (award 5R01MH124839-05). This research has been conducted using the UK Biobank Resource under Application no. 16406. Acknowledgements for each cohort can be found in the Supplementary Info.

## Author contributions

The following authors contributed to:

Project coordination

AH, AIC, AM, AMH, AZ, BMB, CAR, DR, DP, EAS, EU, FB, GS, GT, HMW, HZ, IK, ISk, ITM, JH, JHL, JMG, JMS, JZ, KB, KH, KL, KP, LA, MWL, MWa, MaF, OAA, OB, OF, PFO, RH, RZ, SH, SK, TL, TO, VF, VM, YO

Data collection

23andMe, AAS, ABK, AC, ACvH, AHS, AM, AR, AS, AZ, AdB, BA, BC, BEK, BH, BL, BMT, BOM, CCPM, CE, CJ, CM, ClB, DA, DBDS, DG, DPW, DS, EA, ErS, EstBB, EyS, FHD, FJW, GB, GBW, GHS, GJB, GS, GT, GW, HB, HE, HH, HU, HZ, HaS, HrS, IG, IR, ISa, ISk, ITM, Indiana, JL, JS, JV, KB, KK, KM, KN, KO, KP, KS, LKD, LaP, LeP, LifeLines, MA, MC, MG, MH, MSP, MT, MTB, MVP, MWL, MWa, MWe, MY, Mayo, MoF, MtS, NLP, NT, OBP, OF, PJV, PMBB, PS, QSL, RFS, RS, Regeneron, SBS, SD, SJvdL, SK, SRO, ST, ShB, SvB, TE, TF, TFH, TO, TT, TW, UCLA, VF, WMvdF, YO

GWAS Analysis

23andMe, AAS, CJ, ChB, DPW, DS, EM, EMG, EU, GT, JQT, KO, MRM, NT, PMV, RS, RW, SJvdL, ShB, TO, YO

Quality Control

AAS, AM, AN, AZ, ChB, DPW, HZ, ISk, KB, MWa, OF, QSL, SK, SN, TK, TO, VF

Post GWAS statistical analysis

ALvS, AM, CJ, ChB, DPW, DS, EU, JB, KO, MS, NYB, PMV, QSL, RS, SJvdL, TNP, TO, YO

Writing

AAS, AZ, DP, DPW, EAS, EM, EU, GS, HZ, ISk, JB, KB, MS, MWa, OAA, RW, SD, SK, ShB, TO, VF

## References

1. Nichols, E. et al. Estimation of the global prevalence of dementia in 2019 and forecasted prevalence in 2050: an analysis for the Global Burden of Disease Study 2019. Lancet Public Health 7, e105–e125 (2022).

2. Gatz, M. et al. Role of Genes and Environments for Explaining Alzheimer Disease. Arch. Gen. Psychiatry 63, 168–174 (2006).

3. Holland, D. et al. Beyond SNP heritability: Polygenicity and discoverability of phenotypes estimated with a univariate Gaussian mixture model. PLOS Genet. 16, e1008612 (2020).

4. Wightman, D. P. et al. A genome-wide association study with 1,126,563 individuals identifies new risk loci for Alzheimer’s disease. Nat. Genet. 53, 1276–1282 (2021).

5. Bellenguez, C. et al. New insights into the genetic etiology of Alzheimer’s disease and related dementias. Nat. Genet. 54, 412–436 (2022).

6. Zhang, Q. et al. Risk prediction of late-onset Alzheimer’s disease implies an oligogenic architecture. Nat. Commun. 11, 4799 (2020).

7. Digma, L. A., Winer, J. R. & Greicius, M. D. Substantial Doubt Remains about the Efficacy of Anti-Amyloid Antibodies. J. Alzheimer’s Dis. 97, 567–572 (2024).

8. Kwon, D. Debate rages over Alzheimer’s drug lecanemab as UK limits approval. Nature https://doi.org.10.1038/d41586-024-02720-y (2024) doi:10.1038/d41586-024-02720-y.

9. Baker, E. et al. What does heritability of Alzheimer’s disease represent? PLOS ONE 18, e0281440 (2023).

10. Grotzinger, A. D., Fuente, J. de la, Privé, F., Nivard, M. G. & Tucker-Drob, E. M. Pervasive Downward Bias in Estimates of Liability-Scale Heritability in Genome-wide Association Study Meta-analysis: A Simple Solution. Biol. Psychiatry 93, 29–36 (2023).

11. Han, B. & Eskin, E. Random-Effects Model Aimed at Discovering Associations in Meta-Analysis of Genome-wide Association Studies. Am. J. Hum. Genet. 88, 586–598 (2011).

12. Schipper, M. et al. Prioritizing effector genes at trait-associated loci using multimodal evidence. Nat. Genet. 57, 323–333 (2025).

13. Giambartolomei, C. et al. Bayesian test for colocalisation between pairs of genetic association studies using summary statistics. PLoS Genet. 10, e1004383 (2014).

14. Hartwig, F. P., Davies, N. M., Hemani, G. & Davey Smith, G. Two-sample Mendelian randomization: avoiding the downsides of a powerful, widely applicable but potentially fallible technique. Int. J. Epidemiol. 45, 1717–1726 (2016).

15. Brokate-Llanos, A. M. et al. Ribonucleotide reductase inhibition improves the symptoms of a Caenorhabditis elegans model of Alzheimer’s disease. G3 GenesGenomesGenetics 14, jkae040 (2024).

16. Stuchbury, G., Ko, A. & Dymock, B. A brain penetrant small-molecule SYK inhibitor for the treatment of Alzheimer’s and neuroinflammatory diseases. Alzheimers Dement. 19, e082900 (2023).

17. Wang, S. et al. TREM2 drives microglia response to amyloid-β via SYK-dependent and - independent pathways. Cell 185, 4153-4169.e19 (2022).

18. Kerr, F. et al. Direct Keap1-Nrf2 disruption as a potential therapeutic target for Alzheimer’s disease. PLOS Genet. 13, e1006593 (2017).

19. Lawlor, B. et al. Nilvadipine in mild to moderate Alzheimer disease: A randomised controlled trial. PLOS Med. 15, e1002660 (2018).

20. Chen, G., Ozturk, G., Porta, S. & Lee, V. M.-Y. Evaluate the Efficacy of the Axl inhibitor Bemcentinib in the 5xFAD mouse model of Alzheimer’s Disease. Alzheimers Dement. 20, e089525 (2024).

21. Bayer, T. A. Pyroglutamate Aβ cascade as drug target in Alzheimer’s disease. Mol. Psychiatry 27, 1880–1885 (2022).

22. Coimbra, J. R. M., Moreira, P. I., Santos, A. E. & Salvador, J. A. R. Therapeutic potential of glutaminyl cyclases: Current status and emerging trends. Drug Discov. Today 28, 103644 (2023).

23. Zhang, H. et al. The Retromer Complex and Sorting Nexins in Neurodegenerative Diseases. Front. Aging Neurosci. 10, (2018).

24. Einem, B. von et al. The Golgi-Localized γ-Ear-Containing ARF-Binding (GGA) Proteins Alter Amyloid-β Precursor Protein (APP) Processing through Interaction of Their GAE Domain with the Beta-Site APP Cleaving Enzyme 1 (BACE1). PLOS ONE 10, e0129047 (2015).

25. Cai, X., Xing, J., Long, C. L., Peng, Q. & Humphrey, M. B. DOK3 Modulates Bone Remodeling by Negatively Regulating Osteoclastogenesis and Positively Regulating Osteoblastogenesis. J. Bone Miner. Res. 32, 2207–2218 (2017).

26. Peng, Q., Long, C. L., Malhotra, S. & Humphrey, M. B. A Physical Interaction Between the Adaptor Proteins DOK3 and DAP12 Is Required to Inhibit Lipopolysaccharide Signaling in Macrophages. Sci. Signal. 6, ra72–ra72 (2013).

27. Kim, S.-M. et al. TREM2 promotes Aβ phagocytosis by upregulating C/EBPα-dependent CD36 expression in microglia. Sci. Rep. 7, 11118 (2017).

28. Chung, J. et al. Genome-wide pleiotropy analysis of neuropathological traits related to Alzheimer’s disease. Alzheimers Res. Ther. 10, 22 (2018).

29. Didonna, A. & Opal, P. The promise and perils of HDAC inhibitors in neurodegeneration. Ann. Clin. Transl. Neurol. 2, 79–101 (2015).

30. Jansen, I. E. et al. Genome-wide meta-analysis for Alzheimer’s disease cerebrospinal fluid biomarkers. Acta Neuropathol. (Berl.) 144, 821–842 (2022).

31. Nalls, M. A. et al. Identification of novel risk loci, causal insights, and heritable risk for Parkinson’s disease: a meta-analysis of genome-wide association studies. Lancet Neurol. 18, 1091–1102 (2019).

32. Chen, K. et al. Identifying risk loci for FTD and shared genetic component with ALS: A large-scale multitrait association analysis. Neurobiol. Aging 134, 28–39 (2024).

33. Liu, H. et al. Ubiquitin–proteasome system in the different stages of dominantly inherited Alzheimer’s disease. Alzheimers Dement. 21, e70243 (2025).

34. Xu, N. et al. Ube2v1 Positively Regulates Protein Aggregation by Modulating Ubiquitin Proteasome System Performance Partially Through K63 Ubiquitination. Circ. Res. 126, 907–922 (2020).

35. Masola, V., Greco, N., Tozzo, P., Caenazzo, L. & Onisto, M. The role of SPATA2 in TNF signaling, cancer, and spermatogenesis. Cell Death Dis. 13, 977 (2022).

36. de Leeuw, C. A., Mooij, J. M., Heskes, T. & Posthuma, D. MAGMA: Generalized Gene-Set Analysis of GWAS Data. PLOS Comput. Biol. 11, e1004219 (2015).

37. Verheijen, J. & Sleegers, K. Understanding Alzheimer Disease at the Interface between Genetics and Transcriptomics. Trends Genet. 34, 434–447 (2018).

38. Cissé, M. & Checler, F. Eph receptors: New players in Alzheimer’s disease pathogenesis. Neurobiol. Dis. 73, 137–149 (2015).

39. Pocernich, C. B. & Butterfield, D. A. Elevation of glutathione as a therapeutic strategy in Alzheimer disease. Biochim. Biophys. Acta BBA - Mol. Basis Dis. 1822, 625–630 (2012).

40. Laskovs, M., Partridge, L. & Slack, C. Molecular inhibition of RAS signalling to target ageing and age-related health. Dis. Model. Mech. 15, dmm049627 (2022).

41. Li, S. & Kim, H.-E. Implications of Sphingolipids on Aging and Age-Related Diseases. Front. Aging 2, (2022).

42. Musardo, S. et al. The development of ADAM10 endocytosis inhibitors for the treatment of Alzheimer’s disease. Mol. Ther. 30, 2474–2490 (2022).

43. Nakatsuka, N. et al. A Reproducibility Focused Meta-Analysis Method for Single-Cell Transcriptomic Case-Control Studies Uncovers Robust Differentially Expressed Genes. 2024.10.15.618577 Preprint at 10.1101/2024.10.15.618577 (2025).

44. Kreft, K. L. et al. Abundant kif21b is associated with accelerated progression in neurodegenerative diseases. Acta Neuropathol. Commun. 2, 144 (2014).

45. Rodríguez-Giraldo, M. et al. Astrocytes as a Therapeutic Target in Alzheimer’s Disease– Comprehensive Review and Recent Developments. Int. J. Mol. Sci. 23, 13630 (2022).

46. Kim, M. et al. Maf links Neuregulin1 signaling to cholesterol synthesis in myelinating Schwann cells. Genes Dev. 32, 645–657 (2018).

47. Papuć, E. & Rejdak, K. The role of myelin damage in Alzheimer’s disease pathology. Arch Med Sci 16, 345–341 (2020).

48. Sierksma, A. et al. Novel Alzheimer risk genes determine the microglia response to amyloid-β but not to TAU pathology. EMBO Mol. Med. 12, EMMM201910606 (2020).

49. Kimura, K. et al. Immune checkpoint TIM-3 regulates microglia and Alzheimer’s disease. Nature 641, 718–731 (2025).

50. Wilson, E. N. & Andreasson, K. I. TAM-ping down amyloid in Alzheimer’s disease. Nat. Immunol. 22, 543–544 (2021).

51. Kozlova, A. et al. PICALM Alzheimer’s risk allele causes aberrant lipid droplets in microglia. Nature 646, 1178–1186 (2025).

52. Serneels, L. et al. γ-Secretase Heterogeneity in the Aph1 Subunit: Relevance for Alzheimer’s Disease. Science 324, 639–642 (2009).

53. Lim, F. T., Ogawa, S. & Parhar, I. S. Spred-2 expression is associated with neural repair of injured adult zebrafish brain. J. Chem. Neuroanat. 77, 176–186 (2016).

54. Disouky, A. et al. Human hippocampal neurogenesis in adulthood, ageing and Alzheimer’s disease. Nature https://doi.org.10.1038/s41586-026-10169-4 (2026) doi:10.1038/s41586-026-10169-4.

55. Zheng, Z. et al. Leveraging functional genomic annotations and genome coverage to improve polygenic prediction of complex traits within and between ancestries. Nat. Genet. 56, 767–777 (2024).

56. Jansen, I. E. et al. Genome-wide meta-analysis identifies new loci and functional pathways influencing Alzheimer’s disease risk. Nat. Genet. 51, 404–413 (2019).

57. ReproGen Consortium et al. An atlas of genetic correlations across human diseases and traits. Nat. Genet. 47, 1236–1241 (2015).

58. Evans, L. M. et al. Comparison of methods that use whole genome data to estimate the heritability and genetic architecture of complex traits. Nat. Genet. 50, 737–745 (2018).

59. Liu, S. et al. Alzheimer disease is (sometimes) highly heritable: Drivers of variation in heritability estimates for binary traits, a systematic review. 2025.04.29.25326648 Preprint at 10.1101/2025.04.29.25326648 (2025).

60. Bellenguez, C., Grenier-Boley, B. & Lambert, J.-C. Genetics of Alzheimer’s disease: where we are, and where we are going. Curr. Opin. Neurobiol. 61, 40–48 (2020).

61. Srinivasan, K. et al. Alzheimer’s Patient Microglia Exhibit Enhanced Aging and Unique Transcriptional Activation. Cell Rep. 31, (2020).

62. Mathys, H. et al. Single-cell transcriptomic analysis of Alzheimer’s disease. Nature 570, 332–337 (2019).

63. Gabitto, M. I. et al. Integrated multimodal cell atlas of Alzheimer’s disease. Nat. Neurosci. 27, 2366–2383 (2024).

64. Almeida, V. N. Somatostatin and the pathophysiology of Alzheimer’s disease. Ageing Res. Rev. 96, 102270 (2024).

65. Nitsch, L., Schneider, L., Zimmermann, J. & Müller, M. Microglia-Derived Interleukin 23: A Crucial Cytokine in Alzheimer’s Disease? Front. Neurol. 12, 639353 (2021).

66. vom Berg, J. et al. Inhibition of IL-12/IL-23 signaling reduces Alzheimer’s disease–like pathology and cognitive decline. Nat. Med. 18, 1812–1819 (2012).

67. Nan, F., Azriel, D. & Schwartzman, A. Partitioning Fraction of Variance Explained into Strong Localized Effects and Weak Diffuse Effects. 2026.01.06.697735 Preprint at 10.64898/2026.01.06.697735 (2026).

68. Lam, M. et al. RICOPILI: Rapid Imputation for COnsortias PIpeLIne. Bioinformatics 36, 930–933 (2020).

69. Privé, F., Aschard, H., Ziyatdinov, A. & Blum, M. G. B. Efficient analysis of large-scale genome-wide data with two R packages: bigstatsr and bigsnpr. Bioinformatics 34, 2781– 2787 (2018).

70. Peyrot, W. J., Boomsma, D. I., Penninx, B. W. J. H. & Wray, N. R. Disease and Polygenic Architecture: Avoid Trio Design and Appropriately Account for Unscreened Control Subjects for Common Disease. Am. J. Hum. Genet. 98, 382–391 (2016).

71. Marioni, R. E. et al. Assessing the genetic overlap between BMI and cognitive function. Mol. Psychiatry 21, 1477–1482 (2016).

72. Liu, J. Z., Erlich, Y. & Pickrell, J. K. Case–control association mapping by proxy using family history of disease. Nat. Genet. 49, 325–331 (2017).

73. Weissbrod, O. et al. Functionally informed fine-mapping and polygenic localization of complex trait heritability. Nat. Genet. 52, 1355–1363 (2020).

74. Zou, Y., Carbonetto, P., Wang, G. & Stephens, M. Fine-mapping from summary data with the “Sum of Single Effects” model. PLOS Genet. 18, e1010299 (2022).

75. Sudlow, C. et al. UK Biobank: An Open Access Resource for Identifying the Causes of a Wide Range of Complex Diseases of Middle and Old Age. PLOS Med. 12, e1001779 (2015).

76. Weeks, E. M. et al. Leveraging polygenic enrichments of gene features to predict genes underlying complex traits and diseases. Nat. Genet. 55, 1267–1276 (2023).

77. Evans, D. M. & Davey Smith, G. Mendelian Randomization: New Applications in the Coming Age of Hypothesis-Free Causality. Annu. Rev. Genomics Hum. Genet. 16, 327– 350 (2015).

78. Machiela, M. J. & Chanock, S. J. LDlink: a web-based application for exploring population-specific haplotype structure and linking correlated alleles of possible functional variants. Bioinforma. Oxf. Engl. 31, 3555–3557 (2015).

79. Wingo, A. P. et al. Sex differences in brain protein expression and disease. Nat. Med. 29, 2224–2232 (2023).

80. Jang, B. et al. SingleBrain: A Meta-Analysis of Single-Nucleus eQTLs Linking Genetic Risk to Brain Disorders. medRxiv 2025.03.06.25323424 (2025) doi:10.1101/2025.03.06.25323424.

81. Panousis, N. I. et al. Gene expression QTL mapping in stimulated iPSC-derived macrophages provides insights into common complex diseases. 2023.05.29.542425 Preprint at 10.1101/2023.05.29.542425 (2023).

82. de Klein, N. et al. Brain expression quantitative trait locus and network analyses reveal downstream effects and putative drivers for brain-related diseases. Nat. Genet. 55, 377– 388 (2023).

83. Lopes, K. de P. et al. Genetic analysis of the human microglial transcriptome across brain regions, aging and disease pathologies. Nat. Genet. 54, 4–17 (2022).

84. Kosoy, R. et al. Genetics of the human microglia regulome refines Alzheimer’s disease risk loci. Nat. Genet. 54, 1145–1154 (2022).

85. Jerber, J. et al. Population-scale single-cell RNA-seq profiling across dopaminergic neuron differentiation. Nat. Genet. 53, 304–312 (2021).

86. Humphrey, J. et al. Integrative transcriptomic analysis of the amyotrophic lateral sclerosis spinal cord implicates glial activation and suggests new risk genes. Nat. Neurosci. 26, 150–162 (2023).

87. Humphrey, J. et al. Long-read RNA sequencing atlas of human microglia isoforms elucidates disease-associated genetic regulation of splicing. Nat. Genet. 57, 604–615 (2025).

88. Haglund, A. et al. Cell state-dependent allelic effects and contextual Mendelian randomization analysis for human brain phenotypes. Nat. Genet. 57, 358–368 (2025).

89. Fujita, M. et al. Cell subtype-specific effects of genetic variation in the Alzheimer’s disease brain. Nat. Genet. 56, 605–614 (2024).

90. Bryois, J. et al. Cell-type-specific cis-eQTLs in eight human brain cell types identify novel risk genes for psychiatric and neurological disorders. Nat. Neurosci. 25, 1104–1112 (2022).

91. van Rheenen, W. et al. Common and rare variant association analyses in amyotrophic lateral sclerosis identify 15 risk loci with distinct genetic architectures and neuron-specific biology. Nat. Genet. 53, 1636–1648 (2021).

92. Hoogmartens, J., Cacace, R. & Van Broeckhoven, C. Insight into the genetic etiology of Alzheimer’s disease: A comprehensive review of the role of rare variants. Alzheimers Dement. Diagn. Assess. Dis. Monit. 13, e12155 (2021).

93. Khani, M., Gibbons, E., Bras, J. & Guerreiro, R. Challenge accepted: uncovering the role of rare genetic variants in Alzheimer’s disease. Mol. Neurodegener. 17, 3 (2022).

94. Gazal, S. et al. Linkage disequilibrium–dependent architecture of human complex traits shows action of negative selection. Nat. Genet. 49, 1421–1427 (2017).

95. Lee, S. H., Goddard, M. E., Wray, N. R. & Visscher, P. M. A better coefficient of determination for genetic profile analysis. Genet. Epidemiol. 36, 214–224 (2012).

96. Choi, S. W., Mak, T. S.-H. & O’Reilly, P. F. Tutorial: a guide to performing polygenic risk score analyses. Nat. Protoc. 1–14 (2020) doi:10.1038/s41596-020-0353-1.

97. Pirinen, M., Donnelly, P. & Spencer, C. C. A. Including known covariates can reduce power to detect genetic effects in case-control studies. Nat. Genet. 44, 848–851 (2012).

98. Berisa, T. & Pickrell, J. K. Approximately independent linkage disequilibrium blocks in human populations. Bioinformatics 32, 283–285 (2016).

99. Gelman, A. & Rubin, D. B. Inference from Iterative Simulation Using Multiple Sequences. Stat. Sci. 7, 457–472 (1992).

100. Yang, J., Lee, S. H., Goddard, M. E. & Visscher, P. M. GCTA: A Tool for Genome-wide Complex Trait Analysis. Am. J. Hum. Genet. 88, 76–82 (2011).

101. Chang, C. C. et al. Second-generation PLINK: rising to the challenge of larger and richer datasets. GigaScience 4, (2015).

102. Yang, J., Wray, N. R. & Visscher, P. M. Comparing apples and oranges: equating the power of case-control and quantitative trait association studies. Genet. Epidemiol. 34, 254–257 (2010).

103. Karczewski, K. J. et al. Pan-UK Biobank genome-wide association analyses enhance discovery and resolution of ancestry-enriched effects. Nat. Genet. 57, 2408–2417 (2025).

104. Perez, G. et al. The UCSC Genome Browser database: 2025 update. Nucleic Acids Res. 53, D1243–D1249 (2025).

105. Cerezo, M. et al. The NHGRI-EBI GWAS Catalog: standards for reusability, sustainability and diversity. Nucleic Acids Res. 53, D998–D1005 (2025).

106. Werme, J., van der Sluis, S., Posthuma, D. & de Leeuw, C. A. An integrated framework for local genetic correlation analysis. Nat. Genet. 54, 274–282 (2022).

107. Liberzon, A. et al. Molecular signatures database (MSigDB) 3.0. Bioinformatics 27, 1739–1740 (2011).

108. Koopmans, F. et al. SynGO: An Evidence-Based, Expert-Curated Knowledge Base for the Synapse. Neuron 103, 217-234.e4 (2019).

109. Durinck, S. et al. BioMart and Bioconductor: a powerful link between biological databases and microarray data analysis. Bioinformatics 21, 3439–3440 (2005).

110. Csárdi, G. et al. igraph for R: R interface of the igraph library for graph theory and network analysis. Zenodo 10.5281/zenodo.14736815 (2025).

111. Watanabe, K., Umićević Mirkov, M., de Leeuw, C. A., van den Heuvel, M. P. & Posthuma, D. Genetic mapping of cell type specificity for complex traits. Nat. Commun. 10, 3222 (2019).

112. Siletti, K. et al. Transcriptomic diversity of cell types across the adult human brain. Science 382, eadd7046 (2023).

113. Hodge, R. D. et al. Conserved cell types with divergent features in human versus mouse cortex. Nature 573, 61–68 (2019).

114. Herring, C. A. et al. Human prefrontal cortex gene regulatory dynamics from gestation to adulthood at single-cell resolution. Cell 185, 4428-4447.e28 (2022).

115. Wang, D. et al. Comprehensive functional genomic resource and integrative model for the human brain. Science 362, eaat8464 (2018).

116. Sun, N. et al. Human microglial state dynamics in Alzheimer’s disease progression. Cell 186, 4386-4403.e29 (2023).

