## Supplementary Info for "Genomic analyses reveal new insights into Alzheimer’s disease"

### Table of Contents

*Supplementary Notes* ... *3*

**Supplementary Note 1: Alzheimer's Disease proxy-cases** ... **3**

**Supplementary Note 2: Previously reported loci** ... **5**

**Supplementary Note 3: Exclusion of cohorts that did not correct for batch or array** ... **7**

**Supplementary Note 4: Novel loci gene prioritization** ... **8**

**Supplementary Note 5: Effector gene comparison to previous literature** ... **18**

**Supplementary Note 6: Novel genes as potential drug targets** ... **21**

**Supplementary Note 7: Exclusion of F03 individuals.** ... **23**

**Supplementary Note 8: Loci shared with other neurodegenerative diseases** ... **25**

**Supplementary Note 9: Gene-set analyses** ... **27**

**Supplementary Note 10: Microglia state analyses** ... **28**

**Supplementary Note 11: Genes prioritized through specific cell types** ... **29**

**Supplementary note 12: APOE- ε4 heritability under a liability threshold model** ... **32**

*Supplementary Methods* ... *34*

**Cohorts** ... **34**

All of Us (AoU) ... 34
deCODE Genetics ... 35
HUSK ... 36
Lifelines ... 36
Eli Lilly, Johnson & Johnson, and PROTECT ... 39
Tohoku Medical Megabank (TMM) ... 41
BioBank Japan (BBJ) ... 42
UK Biobank ... 43
DemGene ... 45
VUMC ... 46
Penn Medicine Biobank (PMBB), Mount Sinai Million Health Discoveries Program, INDIANA-CHALASANI, UCLA, Mayo Clinic-RGC Project Generation, Colorado Center for Personalized Medicine – RGC Collaboration, & GHS-RGC DiscovEHR collaboration ... 47
BioVU (Vanderbilt University Medical Center), Gothenburg H70 Birth Cohort Studies and Clinical AD from Sweden, Gr@ce, TwinGene, 23andMe Research Institute, STSA ... 49
HUNT ... 50
IGAP, Shigemizu et al. (2021) ... 52
Million Veteran Program (MVP) ... 52
FinnGen ... 52
Estonian Biobank ... 53

### Supplementary Notes

#### Supplementary Note 1: Alzheimer's Disease proxy-cases

Individuals with first-degree relatives with Alzheimer's disease (AD) will, on average, carry half of the alleles that affect the risk of AD. If these healthy individuals are genotyped, they can be used in a GWAS-by-proxy (GWAX) to identify said risk-affecting alleles, and only a simple scale transformation needs to be applied (see **Methods**) to correct for proxy status. The genetic correlation between the EUR AD-case and proxy-case GWAS was 0.99 (s.e. = 0.14, $p$ = 3e-13). Despite such near-perfect genetic correlation, it has been observed that GWAX can bias cross-phenotype genetic correlation analyses[1]. Specifically, the negative genetic correlation between AD and educational attainment has been shown to flip to a small positive genetic correlation when proxy cases are included in the AD GWAS. Here, we confirm this observation in the EUR GWAS. While not significant, we observe a similar trend in the EAS GWAS. This observed bias is likely due to ascertainment biases, where proxy-cases systematically differ from AD-cases in more phenotypes than just AD-status, and not due to the GWAX methodology per se. Proxy cases are individuals who knew the terms dementia/AD and could remember that their parents had the condition, and may thus, on average, be more highly educated than the average control, which would lead to a positive genetic correlation with Educational Attainment. No bias has been reported in basic GWAS analyses for locus discovery[1], which remains the main objective of our study. Additionally, we found that the inclusion of proxy cases in the meta-analysis did not significantly change the genetic correlation between AD and other neurodegenerative diseases (**Supplementary Results**: Loci shared with other neurodegenerative diseases).

Across the 127 loci identified, 95 were significant in either the multi-ancestry or EUR only analysis that excluded proxy cases and proxy controls. The inclusion of proxy data resulted in an additional 32 significant loci, of which 24 were novel. Of the 48 novel loci, half (24) were only significant with the inclusion of the proxy data. All 48 of the novel loci were at least suggestive ($P<1x10^{-5}$) without the inclusion of proxy data and none were significant in the proxy only data, which suggests that the increased number of significant loci with the inclusion of the proxy data is due to the increased sample size and not due to proxy specific associations. We expect the novel loci to be particularly sensitive to increased sample size because loci that have not been previously identified are likely to have lower effect sizes and thus require larger sample sizes for identification. This was reflected in the data where the median absolute log-odds of lead SNPs in novel loci was 0.036 compared to 0.052 for previously identified loci.

We additionally compared the effect sizes of the lead variants from the 127 significant loci in the case control and proxy only datasets and found a strong correlation (0.98) (**Supplementary Figure 16**). The best fit linear regression model had an intercept of 0.00061 and a slope of 1.069, which shows that the effect sizes of the proxy and case control datasets were extremely similar.

In the EUR GWAS, we identified 93 loci when excluding proxy cases and 11 loci in a proxy-only GWAS; 10 of the 11 proxy loci overlapped with the case-control loci, and the one non-overlapping locus did not reach significance in the combined GWAS. The high genetic correlation between proxy and case-control phenotypes (rg = 0.99, s.e. = 0.14, $p = 3\times10^{-13}$) further supports the validity of proxy inclusion in EUR. In the AFR GWAS, two loci reached genome-wide significance in the case-control GWAS, while one locus was identified in the proxy-only GWAS (APOE). The genetic correlation could not be computed with LDSC due to insufficient sample size. Results were similar in EAS, with two loci in the case-control GWAS and only APOE in the proxy-only GWAS. Although the point estimate of the genetic correlation was low (rg = 0.24), the estimate carries substantial uncertainty (95% CI: −0.40, 0.90), precluding meaningful interpretation. No genome-wide significant loci were identified in AMR or SAS.

#### Supplementary Note 2: Previously reported loci

We identified 118 significant loci in our primary analysis (all ancestries, including proxy cases). 9 additional loci were significant in the European ancestry only analyses (including proxy) and not in the primary analysis (*TMEM163*, *MAP3K13*, *GPAM*, *MCF2L*, *KIAA0125*/IGH gene cluster, *GLCE*, *MCTP2*, *MNT*/*SGSM2*/*DPH1*/*RAP1GAP2*, *AC104532.2*/*VMAC*). This resulted in 127 loci across all subsets of analyses (combined vs. separate ancestries, including proxies vs. excluding proxies). We overlapped these loci with those previously reported in Alzheimer's disease GWAS and rare variant studies (**Supplementary Data 3** and **4**). Across our 127 loci, 48 were not previously reported. Across the 174 loci reported in the common variant previous literature, 78 contained a genome-wide significant ($P<5x10^{-8}$) variant in our primary analysis, and 93 contained a suggestive variant ($P<1x10^{-5}$) (**Supplementary Data 3**). Many of the studies included in previous literature contained a large number of overlapping samples with this study, so these 93 overlapping loci cannot be considered independently replicated. Additionally, the loci from previous literature were defined around the lead variant with a static window (250 kb up and downstream) so overlapping loci may represent distinct loci in close proximity.

We aimed to assess previous GWAS meta-analysis findings using a version of the meta-analysis that used datasets that were not included in previous GWAS. We meta-analyzed the cohorts in our current analysis that did not have overlapping samples with the Wightman et al. (2021)[2] and Bellenguez at al. (2022)[3] cohorts (i.e., AllOfUs, Tohoku Medical Megabank (TMM), Eli Lilly + Johnson&Johnson + PROTECT, Estonian biobank, Genentech, HUSK, Shigemizu et al. (2021), Lifelines, the African-American samples of the Million Veteran Program, GHS, Indiana-Chalasani, Mayo Clinic, Mount Sinai Million Health Discoveries Program, UCLA, Penn Medicine BioBank, Colorado, and the non-EUR sample of the UK Biobank). This resulted a multi-ancestry cohort of 38,581 cases, 978,642 controls, 21,218 proxy cases, and 157,663 proxy controls (Max Neff=145,267). Next, we identified which of the 174 loci reported above were identified in Wightman et al. (2021) and Bellenguez et al. (2022). This resulted in 79 unique loci (**Supplementary Data 6**). Of the 79 loci from Wightman et al. (2021) and Bellenguez et al. (2022), 17 contained a genome-wide significant ($P<5x10^{-8}$) variant in the independent dataset meta-analysis, 31 contained a suggestive variant ($P<1x10^{-5}$), and all 79 contained a nominal variant ($P<0.05$). The 17 regions with a genome significant variant were: *CR1*, *NCK2*, *BIN1*, *HLA*, *TREM2*, *CD2AP*, *TMEM106B*, *ZCWPW1*, *CLU*, *SHARPIN*, *MS4A*, *PICALM*, *SORL1*, *MAPT*, *TSPOAP1*, *ABCA7*, and *APOE*. The additional 15 regions with a suggestive but not genome-wide significant variant were: *PRKD3*, *COX7C*, *TNIP1*, *USP6NL*, *CELF1*, *TPCN1*, *SLC24A4*, *ADAM10*, *APH1B*, *SNX1*, *NTN5*, *LILRB2*, *RBCK1*, and *APP*.

We also aimed to replicate the specific variants reported in Bellenguez et al. (2022) using this independent dataset. We found that 60 of 83 variants were present in the independent set, all of which had the same effect direction as reported in Bellenguez et al. (2022) (**Supplementary Data 7**). 12 of these variants were significant ($P<5x10^{-8}$), 17 were suggestive ($P<1x10^{-5}$), and 53 were nominally significant ($P<0.05$). The seven variants that were present in the independent sample but were not nominally significant were in the following loci: *ADAM17*, *HS3ST5*, IGH cluster, *SCIMP*/*RABEP1*, *MYO15A*, *LILRB2*, and *ADAMTS1*. All of these loci had a nominal variant in the independent sample, suggesting that there may be an association signal in these loci, but with a different lead variant. The *LILRB2* locus contained a suggestive variant in the independent sample (rs2781753; $P=4.15x10^{-7}$), which suggests that there is relatively strong evidence for this locus in the independent sample. Additionally, we found that the effect sizes of lead SNPs correlated strongly ($r = 0.98$; **Supplementary Figure 8**). Overall, we found that many of the loci identified from previous GWAS replicated in an independent dataset. We have made the summary statistics from this independent cohort meta-analysis publicly available for use as a replication cohort (**Data availability statement**).

#### Supplementary Note 3: Exclusion of cohorts that did not correct for batch or array

Three cohorts did not include array or batch as covariates in their GWAS: deCODE, Eli Lilly + Johnson & Johnson + PROTECT, and VUMC. Although this could in principle introduce false-positive associations, inflation metrics indicated well-controlled test statistics in all three, with LDSC intercepts ranging from 0.96 to 1.06 and mean chi-square values from 0.99 to 1.10. Excluding these cohorts reduced the effective sample size by 59,035 and resulted in the loss of 29 genome-wide significant loci. All lost loci were close to the significance threshold in the full analysis (minimum p-value: $1.05\times10^{-9}$) and remained near the significance threshold after exclusion (p-value range: $5.38\times10^{-8}$ to $1.79\times10^{-6}$). This pattern is consistent with a loss of statistical power rather than the removal of false-positive signals.

#### Supplementary Note 4: Novel loci gene prioritization

To select the likely effector gene in the novel loci, we first identified any genes highlighted through an effector gene predictor (FLAMES), the closest gene to the lead SNP, and colocalisation and MR analyses with QTL data (**Supplementary Data 9)**. We then searched previous literature for evidence connecting any of these genes to AD. Finally, we used this evidence to select as few genes as possible to represent the locus as the most likely effector genes (**Supplementary Data 2)**.

In locus 1, two genes (*HP1BP3* and *EIF4G3*) were identified through colocalisation and MR analyses with QTL data, one of which (*EIF4G3*) was the closest gene to the lead SNP. *EIF4G3* was prioritized by FLAMES using the 1000 Genomes reference panel. EIF4G3 has been previously reported as having lower gene expression in AD cases[4]. Knockdown of *HP1BP3* has been associated with cognitive deficits in mice[5]. Based on this evidence, it is difficult to select a single effector gene for this locus.

In locus 2, only one gene (*LAPTM5*) was identified. This gene was prioritized by FLAMES and identified through colocalization and MR analyses with QTL data. The expression of *LAPTM5* was found to have changed in transgenic mouse models of AD and after amyloid deposition[6]. Another study has also linked *LAPTM5* to AD through co-expression network analysis[7]. Based on this evidence, we selected *LAPTM5* as the likely effector gene for this locus.

In locus 3, no gene was prioritized through FLAMES or colocalization and MR analyses with QTL data. *PLEKHO1* was the closest gene to the lead SNP in this locus. We were unable to find any literature supporting the role of *PLEKHO1* in AD. Based on this evidence, we selected *PLEKHO1* as the likely effector gene for this locus but the evidence is not strong.

In locus 6, two genes were prioritized (*CACNA1S* and *KIF21B*). *CACNA1S* was the closest gene to the lead SNP, and *KIF21B* was prioritized through the colocalization and MR analyses with QTL data and was prioritized by FLAMES using the 1000 Genomes reference panel. This region has been identified as shared between AD and MS using GWAS summary statistics[8]. *CACNA1S* is a target of the drug Nilvadipine (https://go.drugbank.com/drugs/DB06712), which was included in a phase 3 trial for AD treatment[9]; however, a previous trial found that Nilvadipine did not reduce cognitive decline[10]. *KIF21B* has been linked to MS and increased gene expression in AD[11,12] and has been linked to neurodevelopmental disorders[13]. Based on this evidence, it is difficult to select a single effector gene for this locus.

In locus 15, only one gene (*ERC2*) was identified. FLAMES prioritized this gene and was the closest gene to the lead SNP. *ERC2* has been found to have increased protein abundance in AD and MCI patients[14] and the gene expression levels has been suggested as a biomarker for AD[15]. Based on this evidence, we selected *ERC2* as the likely effector gene for this locus.

In locus 16, two genes were identified (*NCK1* and *SLC35G2*). *NCK1* was prioritized by FLAMES and, through colocalization and MR analyses with QTL data, was identified as the closest gene to the lead SNP. *SLC35G2* was prioritized through colocalization and MR analyses with QTL data. We were unable to find literature connecting either gene to AD. Based on this evidence, we selected *NCK1* as the likely effector gene for this locus.

In locus 17, only one gene (*MBNL1*) was identified. This gene was prioritized by FLAMES and through colocalization and MR analyses with QTL data, and was the closest gene to the lead SNP. We were unable to find literature connecting either gene to AD. Based on this evidence, we selected *MBNL1* as the likely effector gene for this locus.

In locus 20, only one gene (*MAP3K13*) was identified. FLAMES prioritized this gene and was the closest gene to the lead SNP. We were unable to find literature connecting either gene to AD. Based on this evidence, we selected *MAP3K13* as the likely effector gene for this locus.

In locus 21, three genes were identified (*RNF168, UBXN7,* and *PCYT1A*). *RNF168* was the closest gene to the lead SNP and was prioritized by FLAMES using the 1000 Genomes reference panel. *UBXN7* and *PCYT1A* were identified through colocalization and MR analyses with QTL data. *UBXN7* has been suggested to be a potential transcriptomic biomarker for AD[16]. *PCYT1A* was found to have increased protein abundance in AD brains[17]. Based on this evidence, it is difficult to select a single effector gene for this locus.

In locus 25, three genes were identified (*FNIP2*, *C4orf45*, and *RAPGEF2*). *RAPGEF2* was prioritized by FLAMES (UKB reference panel) and through colocalization and MR analyses with QTL data. *FNIP2* was prioritized through colocalization and MR analyses using QTL data and was prioritized by FLAMES using the 1000 Genomes reference panel. *C4orf45* was the closest gene to the lead SNP. *C4orf45* was identified as pleiotropic for AD and snoring using a cross-trait meta-analysis of GWAS data[18]. *RAPGEF2* levels were found to be higher in transgenic AD mouse models, and silencing *RAPGEF2* expression blocked Aβ oligomer-induced synapse loss in oligomeric Aβ-treated hippocampal neurons[19]. Our MR analysis suggested that more mRNA expression in neurons increases AD risk, which is in line with evidence that more *RAPGEF2* mediates Aβ induced synaptic loss. Based on this evidence, we selected *RAPGEF2* as the likely effector gene for this locus.

In locus 28, two genes were identified (*CEP120* and *SNX2*). Both were prioritized by FLAMES; *CEP120* was the closest gene to the lead SNP, and *SNX2* was prioritized through colocalization and MR analyses with QTL data. SNX2 is in a family of genes (sorting nexins) relevant for endosomal trafficking of app[20]. Based on this evidence, it is difficult to select a single effector gene for this locus.

In locus 32, two genes were identified (*FAM193B* and *DOK3*). *FAM193B* was the closest gene to the lead SNP and *DOK3,* as prioritized through colocalization and MR analyses with QTL data. *FAM193B* was reported to have a male-specific effect on AD in a gene-based GWAS[21]. *DOK3* was found to have lower gene expression in mice carrying *ABCA7* mutations[22]. *DOK3* is also involved in the *TREM2* pathway[23] and interacts with another gene identified in this GWAS (*SYK*)[24]. Based on this evidence, we selected *DOK3* as the likely effector gene for this locus.

In locus 37, two genes were identified (*SUPT3H* and *RUNX2*). *RUNX2* was prioritized by FLAMES and, through colocalization and MR analyses with QTL data, was identified as the closest gene to the lead SNP. *SUPT3H* was prioritized through colocalization and MR analyses with QTL data. We were unable to find literature connecting either gene to AD. Based on this evidence, we selected *RUNX2* as the likely effector gene for this locus.

In locus 39, two genes were identified (*FAM135A* and *C6orf57*). *FAM135A* was the closest gene to the lead SNP and was prioritized by FLAMES using the 1000 Genomes reference panel. Both genes were prioritized through colocalization and MR analyses with QTL data. We were unable to find literature connecting either gene to AD. Based on this evidence, it is difficult to select a single effector gene for this locus.

In locus 40, only one gene (*RGS17*) was identified. This gene was prioritized by FLAMES and through colocalization and MR analyses with QTL data, and was the closest gene to the lead SNP. We were unable to find literature connecting either gene to AD. Based on this evidence, we selected *RGS17* as the likely effector gene for this locus.

In locus 41, two genes were identified (*MAFK* and *INTS1*). *MAFK* was prioritized by FLAMES through colocalization and MR analyses with QTL data. *INTS1* was prioritized through colocalization and MR analyses using QTL data. *INTS1* was differentially expressed in AD patients[25]. Based on this evidence, we selected *MAFK* as the likely effector gene for this locus.

In locus 42, two genes were identified (*PMS2* and *EIF2AK1*). *EIF2AK1* was the closest gene to the lead SNP, and both genes were prioritized through colocalization and MR analyses using QTL data. We were unable to find literature connecting either gene to AD. Based on this evidence, it is difficult to select a single effector gene for this locus.

In locus 45, two genes were identified (*HDAC9* and *SNX13*). *SNX13* was the closest gene to the lead SNP and was prioritized through colocalization and MR analyses with QTL data. *HDAC9* was prioritized by FLAMES. *HDAC9* has been previously linked to neurofibrillary tangle (NFT) and cerebral amyloid angiopathy (CAA) through GWAS[26]. *HDAC9* inhibition has been linked to improved memory in an AD mouse model[27]. Based on this evidence, we selected *HDAC9* as the likely effector gene for this locus.

In locus 50, only one gene (*DOCK4*) was identified. This gene was prioritized by FLAMES and through colocalization and MR analyses with QTL data, and was the closest gene to the lead SNP. *DOCK4* has been identified as pleiotropic for AD and diastolic blood pressure based on GWAS evidence[28]. Based on this evidence, we selected *DOCK4* as the likely effector gene for this locus.

In locus 55, three genes were identified (*ODF1*, *UBR5*, and *RRM2B*). *ODF1* was the closest gene to the lead SNP. *UBR5* was prioritized through FLAMES using the 1000 Genomes reference panel. RRM2B was prioritized through colocalization and MR analyses with QTL data. When an *RRM2B* homolog was targeted by gemcitabine, it led to reduced paralysis in Aβ1-42 C. *elegans* model[29]. Based on this evidence, it is difficult to select a single effector gene for this locus.

In locus 57, two genes were identified (*TG* and *SLA*). *SLA* and TG were the joint closest genes to the lead SNP. *SLA* was prioritized by FLAMES. We were unable to find literature connecting either gene to AD. Based on this evidence, we selected *SLA* as the likely effector gene for this locus.

In locus 59, two genes were identified (*IFNA8* and *IFNE*). *IFNA8* was the closest gene to the lead SNP. *IFNE* was prioritized through colocalization and MR analyses with QTL data. We were unable to find literature connecting either gene to AD. Based on this evidence, it is difficult to select a single effector gene for this locus.

In locus 60, two genes were identified (*GNAQ* and *PSAT1*). *GNAQ* was prioritized by FLAMES and was the closest gene to the lead SNP. *PSAT1* was prioritized through colocalization and MR analyses with QTL data. Based on this evidence, we selected *GNAQ* as the likely effector gene for this locus.

In locus 61, only one gene (*SYK*) was identified. This gene was prioritized by FLAMES and through colocalization and MR analyses with QTL data, and was the closest gene to the lead SNP. *SYK* activation was found to occur following Aβ deposition and the formation of tau pathological species in mice models of AD[30]. Targeted deletion of SYK was linked to higher Aβ deposition, aggravated neuropathology, and cognitive defects in an AD mouse model[31]. When inhibited by a drug (QED-701), a mouse model showed lower neuroinflammation[32]. *TREM2* activates microglia response through *SYK* and the CLEC7A-SYK pathway has been suggested as a potentially druggable pathway based on a mouse model carrying the p.R47H *TREM2* mutation[33]. Our MR analysis suggested that mRNA expression was protective. Based on this evidence, we selected *SYK* as the likely effector gene for this locus.

In locus 62, three genes (*ZNF484*, *FGD3,* and *IPPK*) were identified. *ZNF484* was the closest gene to the lead SNP. *FGD3* and *IPPK* were prioritized through colocalization and MR analyses using QTL data. We were unable to find literature connecting either gene to AD. Based on this evidence, it is difficult to select a single effector gene for this locus.

In locus 68, only one gene (*GPAM*) was identified. FLAMES prioritized this gene and was the closest gene to the lead SNP. We were unable to find literature connecting either gene to AD. Based on this evidence, we selected *GPAM* as the likely effector gene for this locus.

In locus 72, 9 genes were identified (*SNX32*, *CFL1*, *EFEMP2*, *FIBP*, *CCDC85B*, *RNASEH2C*, *BANF1*, *EIF1AD*, and *YIF1A*). *CFL1* was the closest gene to the lead SNP. All of the genes were prioritized through colocalization and MR analyses using QTL data. *CFL1* encodes cofilin, which has been linked to AD in various AD models[34]. *EFEMP2* was found to have decreased gene expression in AD cases[35]. *FIBP* was also found to have decreased gene expression in AD cases[36]. A microglia eQTL for *FIBP* colocalized with AD GWAS signal[37]. *RNASEH2C* has been linked to Aicardi–Goutières syndrome, a progressive neurological disorder linked with neuroinflammation[38]. *BANF1* and *EIF1AD* were linked to tau aggregation in a CRISPRi screen[39,40]. Based on this evidence, it is difficult to select a single effector gene for this locus.

In locus 75, 4 genes were identified (*NINJ2*, *WNK1*, *RAD52*, and *ERC1*). *NINJ2* was prioritized by FLAMES using the UKB reference panel. *WNK1* was the closest gene to the lead SNP and was prioritized by FLAMES using the 1000 Genomes reference panel. *WNK1*, *RAD52*, and *ERC1* were prioritized through colocalisation and MR analyses with QTL data. Genetic variants in *NINJ2* have been linked to AD[41] and *NINJ2* knockout in a cell model was linked to neuroinflammation[42]. Based on this evidence, we selected *NINJ2* as the likely effector gene for this locus.

In locus 76, 6 genes were identified (*SPRYD4*, *GLS2*, *RBMS2*, *IL23A*, *ANKRD52*, and *RPL41*). *GLS2* was the closest gene to the lead SNP. All of these genes were prioritized through colocalisation and MR analyses with QTL data. *GLS2* was found to have lower gene expression in specific cortical layers of AD[43]. *IL23A* encodes the alpha subunit (p19) of Interleukin 23 (IL23)[44]. Knockout of the p19 subunit in an APPPS1 mouse model caused increased Aβ plaque load[44]. However, these results were later refuted by the original authors[45]. *ANKRD52* was found to be differential expressed in female AD patients[46]. Based on this evidence, we selected *GLS2* and *ANKRD52* as potential effector genes.

In locus 78, 4 genes were identified (*HIP1R*, *VPS37B*, *DENR*, and *SBNO1*). *VPS37B* was the closest gene to the lead SNP. All of these genes were prioritized through colocalisation and MR analyses with QTL data. VPS37B was identified as a hub gene for differential AD gene expression[47]. Based on this evidence, it is difficult to select a single effector gene for this locus.

In locus 79, 6 genes were identified (*ATP6V0A2*, *TCTN2*, *GTF2H3*, *DDX55*, *ZNF664*, and *MPHOSPH9*). *TCTN2* was the closest gene to the lead SNP. All of these genes were prioritized through colocalisation and MR analyses with QTL data. We were unable to find literature connecting either gene to AD. Based on this evidence, it is difficult to select a single effector gene for this locus.

In locus 80, 2 genes were identified (*MCF2L* and *LAMP1*). *MCF2L* was prioritized by FLAMES and through colocalisation and MR analyses with QTL data, and was the closest gene to the lead SNP. *LAMP1* was prioritized through colocalisation and MR analyses with QTL data. Increased methylation in *MCF2L* was associated with Braak stages[48]. *LAMP1* was found to accumulate at amyloid beta plaques in APP mouse model[49,50]. Based on this evidence, we selected MCF2L as the likely effector gene for this locus

In locus 85, only one gene (*RASGRP1*) was identified. This gene was prioritized by FLAMES and through colocalisation and MR analyses with QTL data, and was the closest gene to the lead SNP. We were unable to find literature connecting either gene to AD. Based on this evidence, we selected *RASGRP1* as the likely effector gene.

In locus 86, two genes (*PLCB2* and *IVD*) were identified. *PLCB2* was prioritized by FLAMES and through colocalisation and MR analyses with QTL data, and was the closest gene to the lead SNP. *IVD* was prioritized through colocalisation and MR analyses with QTL data. *PLCB2* was found to be differentially expressed in AD[51]. Based on this evidence, we selected *PLCB2* as the likely effector gene.

In locus 90, two genes (*GLCE* and *PAQR5*) was identified. *PAQR5* was prioritized by FLAMES using the 1000 Genomes reference panel. *GLCE* was prioritized by FLAMES using the UKB reference panel and through colocalisation and MR analyses with QTL data, and was the closest gene to the lead SNP. *GLCE* is involved in heparan sulfate proteoglycan biosynthetic process which has been implicated in Alzheimer's disease[52]. Based on this evidence, we selected *GLCE* as the likely effector gene.

In locus 92, only one gene (*MCTP2*) was identified. This gene was prioritized by FLAMES and through colocalisation and MR analyses with QTL data, and was the closest gene to the lead SNP. *MCTP2* was suggestively associated with AD in Korean APOE4 carriers[53]. Based on this evidence, we selected *MCTP2* as the likely effector gene.

In locus 94, 5 genes (*EARS2*, *GGA2*, *DCTN5*, *COG7*, and *SCNN1B*) were identified. *EARS2* was the closest gene to the lead SNP. All of these genes were prioritized through colocalisation and MR analyses with QTL data. *GGA2* was found to interact with *BACE1* in a cell model which then affects *APP* processing[54,55]. Based on this evidence, we selected *GGA2* as the likely effector gene.

In locus 98, 3 genes (*PMFBP1*, *HPR*, and *DHODH*) were identified. *PMFBP1* was the closest gene to the lead SNP. All of these genes were prioritized through colocalisation and MR analyses with QTL data. *DHODH* was prioritized by FLAMES using the UKB reference panel. *PMFBP1* was found to have decreased gene expression in a tau mouse model[56]. *PMFBP1* affects polyamine metabolism, which has been linked to neurodegeneration[57]. Based on this evidence, we selected *PMFBP1* as the likely effector gene.

In locus 101, only one gene (*C16orf95*) was identified. This gene was prioritized by FLAMES and was the closest gene to the lead SNP. *C16orf95* was associated with phosphorylated tau in the CSF in a GWAS[58]. In our local genetic correlation analysis, this locus had a non-significant negative correlation (rg=-0.31, P=0.12). Based on this evidence, we selected *C16orf95* as the likely effector gene.

In locus 104, 4 genes (*MNT, SGSM2, DPH1, and RAP1GAP2*) were identified. *MNT* was the closest gene to the lead SNP. All of these genes were prioritized through colocalisation and MR analyses with QTL data. We were unable to find literature connecting any of these genes to AD. Based on this evidence, it is difficult to select a single effector gene for this locus.

In locus 113, 3 genes (*AC104532.2*, *RFX2*, *VMAC*) were identified. *AC104532.2* and VMAC were the closest genes to the lead SNP. *RFX2* was prioritized through colocalisation and MR analyses with QTL data. *AC104532.2* was found to have upregulated gene expression in *GRN* mutation carriers and FTLD-TDP-A patients[59]. Based on this evidence, it is difficult to select a single effector gene for this locus.

In locus 114, 3 genes (*DNM2*, *SLC44A2*, and *KEAP1*) were identified. *DNM2* was the closest gene to the lead SNP. All of these genes were prioritized through colocalisation and MR analyses with QTL data. Lower gene expression of *DNM2* was linked to Aβ secretion[60] and this gene was identified in a GWAS of AD in Japanese non APOE4 carriers[61]. *KEAP1* has been suggested to be a target for potential drug development for AD[62], where inhibition prevented neuronal toxicity in response to Aβ42[63]. Based on this evidence, it is difficult to select a single effector gene for this locus.

In locus 115, 5 genes (*LRRC25, SSBP4, PIK3R2, MAST3,* and *ARRDC2*) were identified. *LRRC25* was the closest gene to the lead SNP. All of these genes were prioritized through colocalisation and MR analyses with QTL data. Increased protein abundance of *LRRC25* was found in AD brains[64] and an association between decreased expression of *LRRC25* and cognitive impairment was identified through TWAS[65]. Based on this evidence, it is difficult to select a single effector gene for this locus.

In locus 116, 4 genes (*YJEFN3, GATAD2A, TMEM161A,* and *DDX49*) were identified. *GATAD2A* was the closest gene to the lead SNP. All of these genes were prioritized through colocalisation and MR analyses with QTL data. *DDX49* can interact with *APP* in mice[66]. Based on this evidence, it is difficult to select a single effector gene for this locus.

In locus 117, 2 genes (*CEP89* and *CEBPA*) were identified. *CEP89* was prioritized by FLAMES and through colocalisation and MR analyses with QTL data, and was the closest gene to the lead SNP. *CEBPA* was prioritized through colocalisation and MR analyses with QTL data. *TREM2* was found in mice to increase the expression of *CEBPA* (C/EBPα) which promotes *CD36* in microglia and potentially influences Aβ clearance[67]. Based on this evidence, it is difficult to select a single effector gene for this locus.

In locus 118, 4 genes (*AXL, TMEM91, TGFB1,* and *ATP5SL*) were identified. *AXL* was prioritized by FLAMES and through colocalisation and MR analyses with QTL data, and was the closest gene to the lead SNP. All of these genes were prioritized through colocalisation and MR analyses with QTL data. *AXL* was suggested to be relevant for microglia clearance in AD in a mouse model[68]. *AXL* was targeted by a drug with the intention for AD treatment but the drug failed at lowering *AXL* levels in mice[69]. *AXL* CSF levels at baseline have been associated with decreased Aβ CSF levels over time[70]. *AXL* activation has been linked to decreased Aβ in mice[71]. An *AXL* mouse knockout model causes microglia to not respond correctly to Aβ[72]. Increased *TGFB1* expression has been linked to decreased amyloid burden in mice[73,74]. Based on this evidence, we selected *AXL* as the likely effector gene.

In locus 124, 2 genes (*SMOX* and *ADRA1D*) were identified. *SMOX* was prioritized by FLAMES and through colocalisation and MR analyses with QTL data, and was the closest gene to the lead SNP. Both of these genes were prioritized through colocalisation and MR analyses with QTL data. *SMOX* expression levels were found to be lower in LOAD patients[75]. The expression levels of *ADRA1D* have been linked to AD status[76]. Based on this evidence, we selected *SMOX* as the likely effector gene.

In locus 125, 3 genes (*SPATA2, UBE2V1,* and *RNF114*) were identified. *RNF114* was the closest gene to the lead SNP. *SPATA2* and *UBE2V1* were prioritized through colocalisation and MR analyses with QTL data. *RNF114* had higher protein abundance in extracellular vesicles of EOAD patients[77]. *UBE2V1* has been suggested to regulate protein aggregation[78,79]. *SPATA2* is a part of a signaling network that regulates apoptosis[80]. This locus was identified as a shared GWAS locus for ALS and FTD[81]. Our local genetic correlation analyses identified a nominally significant positive genetic correlation with ALS (rg=0.47, P=0.0060). Based on this evidence, we selected *UBE2V1* and SPATA2 as the likely effector gene.

#### Supplementary Note 5: Effector gene comparison to previous literature

We used a tool for effector gene prediction (FLAMES), which tried to nominate an effector gene in each locus based on variant annotation, pathway convergence, and finemapping. We were able to nominate one or more genes for 92/127 loci. Of the 60 loci where we predicted a gene and a gene was nominated in previous literature, there were 10 loci where FLAMES did not predict the gene that was nominated in previous literature.

In locus 8, FLAMES nominated *QPCT* whereas previous literature nominated *PRKD3*[3,82] and *NDUFAF7*[82]. *QPCT* encodes glutaminyl cyclase[83] which catalyzes the formation of pyroglutamate on position 3 of Aβ (AβpE3-*X*)[84]. This specific modification of Aβ is the target of donanemab (an approved anti-amyloid antibody). *PRKD3* has been linked to two aspects of AD pathology; lipid metabolism[85] and neuroinflammation[86]. *NDUFAF7* is involved in the mitochondrial complex 1 and has been suggested to contribute to AD through ROS production[87]. Based on this evidence, *QPCT* is most likely to be the effector gene.

In locus 22, FLAMES nominated *FGFRL1* whereas previous literature nominated *IDUA*[3,88]. *IDUA* has been reported in a Parkinson's disease associated locus[89] but Bellenguez *et al.* (2022)[3] reported that these loci did not colocalise. *FGFRL1* was differentially expressed in cell models carrying APP mutations[90] and other fibroblast growth factors have been implicated in AD[91]. *IDUA* is involved in lysosome function and a recent preprint identified that haploinsufficiency of *IDUA* led to increased plaque burden insoluble $A\beta_{42}$ in a mouse model[92]. Based on this evidence, it is difficult to nominate a single gene as the effector gene.

In locus 23, FLAMES nominated *CLNK*. This gene has been reported in previous GWAS but Schwartzentruber *et al.* (2021)[93] suggested *HS3ST1* because it was linked to cellular uptake of tau[94]. In our FLAMES analysis, *CLNK* had a more support from the variant to gene annotations (0.43 vs 0.13) and pathway convergence (0.63 vs 0.20). *CLNK* functions within the immune system[95] and many of the genes associated with AD in this study are involved in immune functions. This is likely why *CLNK* was prioritized by FLAMES. Based on this evidence it is difficult to select an effector gene within this locus.

In locus 26, FLAMES nominated *FAM105B* (also called *OTULIN*) whereas previous literature nominated *ANKH*[3]. *ANKH* has been linked to excessive mineralisation[96] and potentially inflammation[97]. *FAM105B* has been linked to AD through a TWAS study[98] and a recent preprint found that a knockout of *FAM105B* (*OTULIN*) was associated with a reduction in tau[99] . Based on this evidence, *FAM105B* (*OTULIN*) is more likely to be the effector gene.

In locus 27, FLAMES nominated *AC008394.1* and *RASA1* whereas previous literature nominated *COX7C*[3,82]. *AC008394.1* is uncharacterised so does not have any previous literature linking it to AD. Increased *RASA1* levels in CSF have been linked to increased risk of AD[100] but the primary role of *RASA1* is in vascular development[101]. *COX7C* is involved in the mitochondrial respiratory chain and has been found to be differentially expressed between AD cases and controls[102]. Based on this evidence, it is difficult to nominate a single gene as the effector gene.

In locus 43, FLAMES nominated *RPA3-AS1* whereas previous literature nominated *UMAD1* and *ICA1*[3,88]. *RPA3-AS1* is a long non-coding RNA that has been linked with cancer[103]. *UMAD1* has been suggested to be involved in membrane remodeling[104]. *ICA1* may influence *APP* processing[105]. Based on this limited evidence, *ICA1* is most likely to be the effector gene but further research is needed.

In locus 47, FLAMES nominated *ELMO1* and *GPR141* whereas previous literature nominated *NME8*[3,106] and *EPDR1*[3]. *ELMO1* is a part of a complex involved in internal signalling for microglia to prepare for engulfing apoptotic cells[107]. *ELMO1* was included at the downstream edge of the locus but no variants in LD with the lead SNP of the locus were present in *ELMO1*. *GPR141* is was upregulated in the microglia of AD cases[108] and is involved in immune response[109]. While variants around *NME8* have been associated with AD[110] and AD biomarkers[111], we were unable to find functional studies connecting *NME8* to AD. *EPDR1* was found to be less expressed in AD patients[112]. Overall, *ELMO1* and *GPR141* have the most convincing evidence connecting them to AD pathology.

In locus 52, FLAMES nominated *FDFT1* whereas previous literature nominated *CTSB*[3,88]. *FDFT1* is involved in cholesterol biosynthesis and has been linked to AD through differential expression analysis[113,114]. *CTSB* has also been linked to AD through differential expression[115] and mouse models[116]. Based on this evidence, it is difficult to select a single effector gene for this locus.

In locus 82, FLAMES nominated *SLC24A4*. This gene has been reported in previous GWAS but Schwartzentruber *et al.* (2021)[93] suggested *RIN3* is the likely causal gene in locus due to colocalization with eQTLs in brain tissue and its presence in AD relevant pathways (early endocytic pathway). In our FLAMES analysis, *RIN3* and *SLC24A4* had strong support from the pathway convergence (PoPs raw score was 1.11 and 0.75 respectively) but *SLC24A4* has slightly more support from the variant to gene annotations (XGBoost score was 0.32 vs 0.15). Based on this evidence, it is difficult to select a single effector gene for this locus.

In locus 84, FLAMES nominated *KIAA0125* whereas previous literature nominated the IGH gene cluster[3,88,117]. The expression of *KIAA0125* was observed to decrease as the ratio of Aβ42/Aβ40 increased[118]. The IGH gene cluster encodes proteins necessary for antibodies[119]. We were unable to find any experimental evidence connecting the IGH gene cluster to AD but multiple genetic risk factors have immune system functions so the IGH genes may also be relevant to AD. Based on this evidence, it is difficult to select a single effector gene for this locus.

In locus 89, FLAMES nominated *FAM96A* whereas previous literature nominated the *SNX1*[3]. *FAM96A* has been identified as differentially expressed between AD cases and controls in the cortex in women[120]. *SNX1* is a sorting nexin which forms complexes with other sorting nexins to mediate endosomal trafficking[20]. Deficiencies in these proteins have been linked to impaired trafficking of *app* leading to increased burden of Aβ. Based on this evidence, *SNX1* is likely to be the effector gene for this locus.

In locus 107, FLAMES using the UKB reference panel nominated *ITGA2B* whereas previous literature and FLAMES using the 1000 Genomes panel nominated *GRN*[2,3,88]. *ITGA2B* codes for integrin αIIbβ3 which binds to Aβ and is linked to increased Aβ fibril formation[121]. *GRN* encodes progranulin which is strongly associated with frontotemporal dementia and multiple neurodegenerative processes[122]. Based on this evidence, it is difficult to select a single effector gene for this locus.

#### Supplementary Note 6: Novel genes as potential drug targets

Across the 48 novel loci, we identified 5 genes that have been tested as drug targets for AD (*CACNA1S*, *RRM2B*, *SYK*, *KEAP1*, and *AXL*). *CACNA1S* encodes a subunit of a calcium channel and is a target of Nilvadipine https://go.drugbank.com/drugs/DB06712. Nilvadipine is a calcium channel blocker used for treatment of hypertension[10]. Experimental work in transgenic mice showed that Nilvadipine can lower Aβ40 and Aβ42 levels. However, a clinical trial published in 2018 showed no reduction in cognitive decline. A limitation of that trial was that it included current cases so it is unknown how Nilvadipine would affect AD development prior to plaque build up. While our GWAS identified the *CACNA1S* locus, another gene in that locus (*KIF21B*) has been linked to MS and increased gene expression in AD brains[11,12] so it is uncertain whether *CACNA1S* is the effector gene in that locus.

*RRM2B* encodes a subunit of a ribonucleotide reductase which is involved in DNA repair of mitochondrial DNA[123]. A study inhibited a homolog of *RRM2B* in an Aβ model of *C. elegans* with gemcitabine and found that treatment improved the outcome (reduction of paralysis caused by build up of Aβ)[29]. Gemcitabine is a chemotherapeutic agent used to treat cancer and we were unable to find other literature to support the potential of Gemcitabine for AD so based on current evidence the risk of treatment for AD is likely to outway the reward.

*SYK* encodes a pro-inflammatory signalling kinase that mediates the role of *TREM2* for microglia response to Aβ plaques[33]. Interestingly, the direction of effect for *SYK* is mixed in the literature. Some studies suggest that increasing *SYK* activity is linked to decreased AD pathology[31,33] while others suggest that reduction of *SYK* activity is linked to decreased AD pathology[30,32]. One of these studies used an antibody for *CLEC7A* to activate *SYK* which led to partial rescue of the microglial responses to Aβ[33]. Another used a *SYK* inhibitor (QED-701) and found decreased neuroinflammation in a mouse model[32]. While the exact mechanism is not clear, *SYK* does seem to be an interesting drug target for further study.

*KEAP1* encodes an inhibitor of Nrf2, which is a transcription factor that activates genes to respond to oxidative stress[63]. Activators of Nrf2 have been tested as treatment for AD in mouse models but off-target effects have been reported. Inhibition of *KEAP1* has been suggested as a potential mechanism to increase Nrf2 activity. *KEAP1* has shown promise in AD cell and animal models[63] and various compounds have been suggested as potential drugs for targeting *KEAP1* for AD treatment[62]. We were unable to find any clinical data for drug targeting *KEAP1* but this gene represents an interesting candidate for further research.

*AXL* encodes a TAM receptor tyrosine kinase which was found to be relevant for microglia response to Aβ in mice[72]. A drug (bemcentinib) targeting *AXL* was tested in an AD mouse model but this drug did not cause a reduction in *AXL* levels and no improvement in the phenotype of the mice was observed[69]. While that specific drug did not show efficacy, AXL remains as an interesting candidate for further research.

#### Supplementary Note 7: Exclusion of F03 individuals.

F03 is an ICD-10 code for unspecified dementia, which may capture individuals with Alzheimer's Disease (AD) alongside those with other neurodegenerative conditions. Inclusion of F03 individuals may bias pleiotropy analyses with other neurodegenerative diseases. To test for potential bias, we excluded 13 cohorts that used F03 as part of their AD case definition: All of Us, UK Biobank, Penn Medicine Biobank (PMBB), Mount Sinai Million Health Discoveries Program, INDIANA- CHALASANI, UCLA, Mayo Clinic-RGC Project Generation, Colorado Center for Personalized Medicine – RGC Collaboration, GHS-RGC DiscovEHR collaboration, HUNT, Estonia biobank, BioVU, Copenhagen Hospital Biobank and Danish Blood Donor Study, collectively contributing 33,072 cases to the primary meta-analysis. These same cohorts also screened controls for the absence of other neurodegenerative disorders. Exact counts of individuals diagnosed exclusively via F03 are unavailable for most cohorts; however, in the UK Biobank, 2,361 out of 5,772 cases (40.9%) carried only F03 ICD-10 codes. Extrapolating this proportion, we estimate that approximately 13,526 cases across these seven cohorts were diagnosed with F03 only. We performed a sensitivity meta-analysis excluding all 13 cohorts and re-evaluated local genetic correlation and cell-type enrichment analyses.

At the *UBE2V1/SPATA2* locus, the local genetic correlation between AD (EUR, excluding proxy cases) and amyotrophic lateral sclerosis (ALS) strengthened in the sensitivity analysis, increasing from $rg = 0.47$ ($p = 0.006$) to $rg = 0.62$ ($p = 0.002$). At the *TMEM163* locus, the local genetic correlation between AD and Parkinson's Disease remained at $rg = -1.00$, with a slightly stronger association ($p = 0.0004$ versus $p = 0.0006$ in the primary analysis). These results indicate that the pleiotropic signals at both loci are not driven by the inclusion of F03-defined cases.

In the differential cell-type enrichment analysis, associations for three neuronal cell types were attenuated relative to the primary analysis: Sncg neurons ($p = 0.00018$ vs. $p = 0.0036$), Sst neurons ($p = 0.00021$ vs. $p = 0.00034$), and L6_IT_Car3 neurons ($p = 0.0012$ vs. $p = 0.088$). Applying a Bonferroni correction for the number of cell types tested ($\alpha = 0.05/42 = 0.00119$), the Sst neuron association remained significant, while the Sncg neuron and the L6_IT_Car3 neuron association was no longer significant. Given the substantial reduction in sample size by excluding seven cohorts, this attenuation may be explained by reduced statistical power rather than bias introduced by F03-defined cases.

Taken together, these sensitivity analyses support the conclusion that including F03-defined cases did not bias the pleiotropy analyses, but that it slightly reduced the observed cell-type enrichment, likely reflecting reduced statistical power

#### Supplementary Note 8: Loci shared with other neurodegenerative diseases

Of the 127 loci identified in this study, 18 have been previously associated with Parkinson's disease (PD), frontotemporal dementia (FTD), or amyotrophic lateral sclerosis (ALS) (**Supplementary Data 12**). There were 14 loci shared with PD, 4 loci shared with FTD, and 3 loci shared with ALS. Two loci were shared across more than 2 neurodegenerative diseases; the *HLA* region was associated with AD, PD, FTD, and ALS, and locus 52 (*CTSB*) was associated with AD, PD, and FTD. We were interested in whether the known neurodegenerative loci were associated with AD due to the inclusion of other neurodegenerative disease cases in the proxy AD datasets. In the meta-analysis of datasets that did not contain proxy AD phenotypes, the lead SNPs of the 18 loci had p-values between $2x10^{-5}$ and $1x10^{-26}$. This suggests that case misspecification in the proxy AD datasets did not drive the associations of these loci. Additionally, the LDSC genetic correlations between AD and PD were 0.18 (SE=0.052, P=0.0004) using the combined dataset and 0.17 (SE=0.055, P=0.0023) when the proxy datasets were excluded, which suggests that the inclusion of the proxy datasets was not causing the GWAS results to be more similar to PD GWAS. The same was true for ALS; 0.21 (SE=0.064, P=0.0009) using the combined dataset and 0.24 (SE=0.068, P=0.0005) when the proxy datasets were excluded. Overall, the genetic overlap between AD, ALS, FTD, and PD was unlikely due to the inclusion of proxy cases.

We further explored these shared loci using colocalization and local genetic correlation analyses to identify whether these loci shared the same causal variants. We were able to find summary statistics[124,125] for 14 of the 18 loci and then tested these loci using coloc[126] and LAVA[127] (**Supplementary Data 12**). We found that the *UBE2V1/SPATA2* locus had a positive genetic correlation between ALS and AD ($r_g$=0.47, P=0.0060) and colocalized (PP.H4=0.95), with the most evidence for the lead SNP of this locus as the shared variant (20:48578734:A:T/rs113558364; SNP.PP.H4=0.39). This suggests that this locus is shared between ALS and AD and that there is evidence for rs113558364 being the shared causal variant. The *GBA1* locus colocalized between AD and PD (PP.H4=0.99), with the lead SNP of the locus the most likely shared variant (1:155135036:A:G/rs35749011; SNP.PP.H4=0.97). The *TMEM163* locus had a strong negative genetic correlation ($r_g$=-1, P= 0.00057) and is likely driven by distinct variants (PP.H3=0.66). These results suggest that *UBE2V1/SPATA2* is a shared locus between AD and ALS, *GBA1* is a shared locus between AD and PD, but the *TMEM163* locus is likely to have opposite effects in AD and PD through different causal variants.

To test whether these overlapping loci may account for the global genetic correlations between AD and PD, and AD and ALS, we repeated the global genetic correlation analysis after excluding these loci. The global genetic correlations changed only minimally for PD (0.15 vs. 0.17) and remained unchanged for ALS (0.24).

We were interested in whether these loci influenced gene expression in the same cell types across diseases. We performed colocalization and Mendelian randomization analysis with multiple QTL datasets to determine whether disease-associated variants influenced expression. We identified that the PD-associated variants in the *TMEM163* locus were linked with increased expression of *TMEM163* in microglia and macrophages, but the AD-associated variants were linked with decreased expression in neurons (**Supplementary Data 12)**. This suggests that the effect of the distinct causal variants in this locus may be mediated through different cell types in AD and PD. We identified that the AD and ALS variants in the *UBE2V1/SPATA2* locus were linked with increased expression of *SPATA2* in brain tissue and *UBE2V1* in neurons. This suggests that the effect of this locus is likely mediated through the same cell type in AD and ALS, with evidence for both *SPATA2* and *UBE2V1* being the effector gene.

#### Supplementary Note 9: Gene-set analyses

Using MAGMA v1.10 (SNP-wise mean model), we identified 72 significant MSigDB and SynGO gene sets at a false discovery rate of 5% (Benjamini-Hochberg) (**Supplementary Data 13**). We performed pairwise conditional analyses on these genes and clustered them based on the proportion of marginal association explained by the other gene set for all pairs of gene sets. This resulted in 13 clusters and 21 gene sets which were not included in any cluster (**Supplementary Data 13**). Of these 13 clusters, 12 had functions that could be easily connected to four processes identified in previous AD GWAS[2,3,128] (lipid binding, immune activation, amyloid catabolism, and cell adhesion). In addition to these processes, there was 1 cluster (astrocytes/cell fibres) with a group of gene sets that did not suggest a clear biological process. The gene sets in this cluster contained *PSEN1*, *PSEN2*, and *APP*, so it is likely that this cluster was associated with AD through biological processes related to amyloid catabolism. Across the clustered gene sets we identified 4 broad biological mechanisms: lipid binding, immune activation, amyloid catabolism, and cell-cell adhesion. We performed the same gene-set analysis using a meta-analysis of cohorts not included in Wightman et al. (2021) or Bellenguez et al. (2022) (see section "Previously reported loci"). Restricting to these cohorts reduced the effective sample size by a factor of 3 (~415k to ~145k). The gene sets related to amyloid catabolism and B cell activation were identified at a false discovery rate of 5% (**Supplementary Data 13**). The reduced sample size may explain the lack of association with gene sets related to cell-cell adhesion and lipid binding.

In addition to the 13 clusters of gene sets, there were 21 gene sets that did not form clusters. Of these 21 gene sets, there was one gene-set (ROVERSI_GLIOMA_COPY_NUMBER_UP) where 85% of the gene-set association was due to chromosome 19 which suggests that this association was largely due to the extremely strong *APOE* association. The remaining 20 gene sets highlighted immune activation, EphrinB reverse signaling, oxidative stress due to glutathione metabolism, RAS signaling, senescence through sphingolipid metabolism, nuclear envelope lumen, and synapse endocytosis. EphrinB reverse signaling[129], oxidative stress due to glutathione metabolism[130], RAS signaling[131], senescence through sphingolipid metabolism[132], and synapse endocytosis[133] have been connected to AD pathology in previous literature but not through GWAS gene set analysis.

#### Supplementary Note 10: Microglia state analyses

In our previous analysis, we found that microglia express AD-related genes more highly than other brain cell types, with increased levels in affected cases. We next performed MAGMA gene set analysis to identify whether genes defining microglia states were enriched for AD GWAS association. We used the microglia state definitions defined in Sun et al. (2023)[134] as gene sets. We found that no microglia state was significant after Bonferroni correction for 12 states ($P<0.05/12$) (**Supplementary Figure 11**; **Supplementary Data 15**). There were five states with nominal associations: Inflammatory_II, Homeostatic, Inflammatory_III, Stress_signature, Glycolytic. All of these states except the homeostatic are related to inflammation (inflammatory and Glycolytic[135]) or response to misfolded proteins (Stress_signature). When using the LD reference data from 1000 Genomes, the Inflammatory_II state was associated ($P=6.72 \times 10^{-5}$). While the gene set associations were not significant when using the UK Biobank LD reference data, our results suggest that microglia states related to inflammation and stress are characterized by genes associated with AD.

#### Supplementary Note 11: Genes prioritized through specific cell types

We performed three types of analyses that connected the GWAS results to specific cell types. First, we used colocalization and MR in QTL data of specific cell types to prioritize genes. Next, we used MAGMA gene property analysis to identify that microglia overexpress genes linked to AD compared to the average expression in the brain datasets. Finally, we used differential gene expression data to identify that microglia of cases overexpress genes linked to AD compared to controls and neurons of cases under express genes linked to AD compared to controls. Using these results, we have prioritized genes associated with AD that may have cell type specific influences on AD.

We aggregated the significant colocalization and MR results per cell type to identify 24 loci where only a single cell type was implicated (**Supplementary Table 9**). Across 24 loci, 35 genes were prioritized, in 8 loci multiple genes were prioritized for the same cell type. Only one gene was linked to astrocytes, *KIF21B* in locus 6, which colocalised with an eQTL in astrocytes. Our MR analysis indicated that increased expression of *KIF21B* was linked with AD risk. Increased expression of *KIF21B* has been previously linked to astrocyte activation and AD[11]. Abnormal astrocyte activation has been linked with increased Aβ production and release[136]. This suggests that overexpression of *KIF21B* may influence AD through abnormal astrocyte activation leading to increased Aβ pathology. One gene (*MAF*) was linked to oligodendrocytes with decreased expression of *MAF* linked to increased AD risk in our MR analysis. Knockout of *MAF* has been associated with decreased myelination[137] and decreased myelin has been linked to AD[114]. We speculate that decreased *MAF* expression may cause decreased myelin production and make neurons more susceptible to the effects of AD pathology. Interestingly, genes that regulate myelin have been implicated in transcriptomic analyses of AD[138]. In this analysis, we chose to focus on loci where only a single cell type was implicated to aid translation of findings to experimental models; however, an absence of colocalization in other cell types does not mean that the gene only impacts AD through a single cell type. Future experimental research is required to confirm the specificity of the findings.

Of the 23 genes that were only linked to macrophages or microglia, five (*MBNL1*, *SYK*, *BIN1*, *DOK3*, *BLNK*) had the same effect direction in our MR analyses and in literature. Decreased expression of *MBNL1* was linked with increased AD risk in our MR analysis and overexpression was linked to M2 type microglia polarization and ischemic stroke recovery[139]. The protective effect of overexpression of *MBNL1* in microglia in stroke recovery may also mitigate AD risk. Decreased expression of *SYK* was linked to increased AD risk in our MR analysis and previous literature identified that deletion of *SYK* in microglia led to exacerbated Aβ deposition, aggravated neuropathology, and cognitive defects in a mouse model[31]. Increased expression of *DOK3* was linked to increased AD risk in our MR analyses and overexpression of *DOK3* has been linked to AD previously[140]. *DOK3* interacts with known AD genes *TREM2* and *INPP5D*, which suggests that further genes relevant to AD may be found in this network. Overexpression of *BLNK* was linked to AD risk in our MR analysis and increased expression of *BLNK* has been observed after Aβ exposure[141]. *BLNK* along with 4 other genes identified in this GWAS (*SYK*, *INPP5D*, *FCER1G*, and *PLCG2*) have been suggested to play a role in FC gamma receptor-mediated phagocytosis[141]. These results highlight genes that are involved in microglia activation and phagocytosis.

To prioritize genes driving the microglia association in the cell type analysis of non-diseased individuals, we overlapped the top decile of microglia expressed genes (log2 fold change vs dataset average) from the most significant dataset from each study (67_Siletti_Hippocampus.HiB.RostralCA1-2_Human_2022_level2, GSE168408_Human_Prefrontal_Cortex_level2_Adult, PsychENCODE_Adult, Allen_Human_MTG_level2). We then restricted the overlapping overexpressed genes to genes with a significant gene level AD association (MAGMA gene P<0.05/18439). This resulted in 30 genes that were overexpressed in microglia and have been associated with AD in the GWAS (**Supplementary Data 14**). All of these genes were present in one of the loci reported in **Supplementary Data 2** with *PLCB2*, *DOK3* and *AXL* being in loci new to this study. Interestingly, 7 genes (*PICALM*[142], *BLNK*[141], *HAVCR2*[143], *FCER1G*[141], *AXL*[68], *INPP5D*[141], and *PLCG2*[141]) have been linked to microglial phagocytosis in literature. This suggests that other genes involved in microglial phagocytosis may be good candidates for future study.

To prioritize genes driving the microglia association in the differential gene expression cell type analysis of cases vs controls, we identified genes that were nominally overexpressed (P<0.05) and had a significant gene level AD association (MAGMA gene P<0.05/18439). This resulted in 31 genes that were overexpressed in microglia of cases and have been associated with AD in the GWAS (**Supplementary Data 16**). Of these, 12 (*SPI1*, *CASS4*, *TREM2*, *MS4A6A*, *APOE*, *APOC1*, *BIN1*, *SORL1*, *AXL*, *SIGLEC9*, *PILRA*, *CD14*) were prioritized in the cell type analysis of non-disease individuals. This suggests that these 12 genes are active in microglia generally and are over active in AD cases. One gene (*MALT1)* was prioritized despite not falling within a GWAS associated locus. The most significant variant within this gene was just below significance (rs1059442: $P=7.27 \times 10^{-8}$); however, there was sufficient signal within this gene to meet the gene level significance threshold. *MALT1* has been linked to myeloid activation[144] and therefore may play a role in microglial activation in AD.

To prioritize genes driving the neuron association in the differential gene expression cell type analysis of cases vs controls, we overlapped the nominally under expressed genes (P<0.05) from the 3 significant neuron types (Sncg_Neuron, Sst_Neuron, and L6_IT_Car3_Neuron). We then restricted the overlapping overexpressed genes to genes with a significant gene level AD association (MAGMA gene P<0.05/18439). This resulted in 3 genes (*ZKSCAN1*, *APH1B*, *SPRED2*) that were under expressed in all three neuron types and were associated with AD in the GWAS (**Supplementary Data 16**). *APH1B* encodes a part of the γ-secretase complex that cleaves *App*[145] and may be linked to AD through misprocessing of *App* in neurons. Decrease in *SPRED2* has been observed after brain injury in a zebrafish model and has been suggested to lead to neuro-regeneration[146]. The association of *SPRED2* in the GWAS may be due to variants impacting *SPRED2* expression leading to impaired neuronal response to neurodegeneration.

Overall, we prioritized specific genes and biological processes relevant to AD by combining AD GWAS results with QTL and gene expression data. These genes and biological processes were mediated by 4 cell types: astrocytes, oligodendrocytes, microglia, and neurons. Astrocyte activation was identified through an association with *KIF21B*. Myelin production in oligodendrocytes was identified through an association with *MAF*. Microglial phagocytosis was identified through *INPP5D, SYK*, *BLNK*, *HAVCR2*, *PICALM*, *AXL*, *FCER1G*, and *PLCG2*. Repair after damage in neurons was identified through *SPRED2*.

#### Supplementary note 12: APOE- ε4 heritability under a liability threshold model

We assume a population prevalence of Alzheimer's disease of $K$ = 0.05. The *APOE*-ε4 allele frequency in our study was $p$ = 0.1786 and and per-allele log-odds ratio was β = 1.0037 (odds ratio $OR = e^{\beta} \approx 2.728$). Under a multiplicative model on the odds scale, the heterozygote and homozygote odds ratios relative to the ε4-noncarrier baseline are $OR$ and $OR^2$, respectively.

Let $K_{00}$, $K_{01}$ and $K_{11}$ denote the probability of disease given 0, 1, and 2 copies of ε4, respectively. Under the multiplicative-odds model, $K_{01}$ and $K_{11}$ are determined by $K_{00}$:

$$K_{01} = OR \cdot \text{odds}(K_{00}) / [1 + OR \cdot \text{odds}(K_{00})],$$

$$K_{11} = OR^2 \cdot \text{odds}(K_{00}) / [1 + OR^2 \cdot \text{odds}(K_{00})]$$

where odds($x$) = $x/(1-x)$. Assuming Hardy–Weinberg equilibrium, the genotype frequencies are $(1-p)^2$, $2p(1-p)$, and $p^2$ for the three genotypes, and the overall prevalence is

$$K = (1-p)^2K_{00} + 2p(1-p)K_{01} + p^2K_{11}.$$

Assuming K = 0.05, we solved numerically by one-dimensional root-finding (*uniroot* in R) to obtain K_00, and then $K_{01}$ and $K_{11}$ from the equations above.

Under the liability threshold model, disease occurs when an individual's latent liability exceeds a genotype-specific threshold $t_g = \Phi^{-1}(1 - K_g)$, where Φ is the standard-normal cumulative distribution function. Equivalently, each genotype can be assigned a mean liability $u_g = t_K - t_g$, where $t_K = \Phi^{-1}(1 - K)$ is the common threshold corresponding to the overall prevalence.

| Genotype | Frequency | Risk (K) | Mean liability (u) |
|---|---|---|---|
| ε4 noncarrier (00) | 0.6753 | 0.0306 | −0.227 |
| ε4 heterozygote (01) | 0.2936 | 0.0793 | 0.235 |
| ε4 homozygote (11) | 0.0319 | 0.1903 | 0.768 |

We define the additive and dominance components at a biallelic locus as $a = (u_{11} - u_{00})/2$ and $d = u_{01} - (u_{00} + u_{11})/2$. The average effect of allele substitution is

$$\alpha = a + (1 - 2p)\, d,$$

and the additive and dominance variance contributed by the locus is

$$V_A = 2p(1 - p)\, \alpha^2$$

$$V_D = (2p(1 - p)*d)^2$$

Assuming the residual liability is standard-normal with variance $V_E$ = 1, the liability-scale heritability ($h^2_{liab}$) attributable to *APOE*-ε4 is

$$h^2_{liab} = V_A / ((V_A + V_D) + V_E).$$

With the inputs above, we get $a$ = 0.498, $d$ = −0.035, and α = 0.475, giving $V_D$ = 0.0001, $V_A$ = 0.0662 and $h^2_{liab}$ **= 6.2%**. In other words, the *APOE*-ε4 allele alone accounts for roughly 6.2% of the variance in Alzheimer's disease liability under a prevalence of 5%.

A simple linear transformation of the per-allele log-odds ratio to the liability scale yields a very similar result. We first convert the log-odds ratio using $b_{liab}$ = $b_{logit} \times K \times (1 - K)/z$, where $z$ is the height of the standard-normal density at the threshold $t_K$ corresponding to the overall prevalence (i.e., dnorm(qnorm(K)) in R). Using the values above, this gives $b_{liab}$ = 0.462 (= $\alpha^2$) and $V_A$ = $2p(1 - p)\alpha^2$ = 0.063, yielding $h^2_{liab}$ = 5.9%. This closely matches the value of 6.2% obtained above.

### Supplementary Methods

#### Cohorts

##### All of Us (AoU)

This study used the data from the AoU Research Program Curated Data Repository (Controlled Tier Dataset version 7), accessed via the AoU Researcher Workbench. AD cases were identified using ICD-10 codes G30, F00*, and F03; individuals with a first AD diagnosis before age 40 were excluded. Controls were defined as individuals without any diagnostic codes for AD, mild cognitive impairment, or other neurodegenerative diseases, including ICD-10 codes G30, F00*, F03, F06.7, G20–G26, G31–G32, G35–G37, F02, and G10–G14.

Genetic ancestry inference was performed by AoU using genetic similarity between each individual and global population reference panels to assign ancestry groupings for all WGS samples. Individuals of East Asian (EAS), Middle Eastern (MID), and South Asian (SAS) were excluded from the analysis due to fewer than 100 AD cases in each group. Genotyping was performed using the Illumina Global Diversity Array (GDA). The APOE-e4 variant was not present on this array and was not imputed. GWAS were conducted using PLINK v1.90. For genotyping QC, we included variants with call rate ≥ 0.98 and MAF ≥ 0.01. Hardy-Weinberg equilibrium (HWE) test was performed in each ancestry group separately, variants were excluded if HWE p-values were $< 1\times10^{-6}$ in controls or $< 1\times10^{-10}$ in cases within each ancestry group. For individual QC, we excluded individuals with low call rate (>2%), inbreeding coefficient (FHET) outside ±0.20, and close genetic relatedness (kinship coefficient ≥ 0.044). Association analyses included the first 20 principal components (calculated separately within each ancestry group), age, sex, $age^{2,}$ and age*sex as covariates. The final case-control dataset consisted of 1,621 AD cases and 190,991 controls, with a total of 77,856 males and 114,756 females. The age range of participants was 17.6-113.7 years old, with a mean age of 71.5 years old for cases and 50.8 years old for controls.

We derived AD-by-proxy status based on responses to two questions from the Personal and Family Health History: (1) “Have you or anyone in your family ever been diagnosed with the following brain and nervous system conditions?” and (2) “Including yourself, who in your family has had dementia (includes Alzheimer's, vascular, etc.)?”. Participants who responded “None” to the first question were coded as 0 (no family history of brain or nervous system conditions). Any participants who identified either their mother or father in response to the second question were coded as 1. GWAS for AD-by-proxy were conducted separately for individuals reporting maternal versus paternal history of dementia, within each ancestry group. The final proxy GWAS included 3,613 maternal cases and 4,562 controls, and 2,084 paternal cases and 4,608 controls. Participant ages ranged from 19.31 to 104.19 years, with a mean age of 66.2 years for cases and 56.1 years for controls.

Acknowledgement: We gratefully acknowledge AoU participants for their contributions, without whom this research would not have been possible. We also thank the National Institutes of Health's All of Us Research Program for making available the participant data examined in this study.

**deCODE Genetics**

Approval for this study was obtained from the National Bioethics Committee and the Icelandic Data Protection Authority (VSN-19-129). Written informed consent was obtained from participants or their legal guardians prior to blood sample collection, and all sample identifiers were encrypted in accordance with the regulations of the Icelandic Data Protection Authority. The Icelandic Alzheimer's cohort (N=9,559, male=3,621, female=5,938, mean yob=1,928.4 (SD 11.2)) has been previously described[147]. Briefly, a subset of patients was diagnosed with definite, probable, or possible AD based on the NINCDS-ADRDA criteria[148]. For another subset, diagnoses were made according to the International Classification of Diseases, 10th Revision (ICD-10), using codes F00 or G30, or individuals who were prescribed donepezil (Aricept). The case group was compared to population controls (male=132,044, female=124,954, mean yob=1953.7 (SD 26,7)). AD onset data were calculated from the first available Alzheimer's diagnosis or Aricept/Donepezil prescription. The mean age at onset was 80.4 years with a standard deviation of 7.8 years.

The sample was genotyped using Illumina: HumanHap, Omni and Infinium Global Screening SNP arrays as previously described[147]. BeadStudio (Illumina; version 2.0) was used to call genotypes. To account for the relatedness and stratification within our case and control sample sets, we applied the method of genomic control based on chip markers. For the AD GWAS, the correction factor based on the genomic control was 1.32. This elevated value reflects the well-documented inflation of test statistics in the Icelandic population due to relatedness among individuals[149]. After correction, the LD Score regression intercept was 0.96, and the mean chi-square was 0.99, indicating no residual inflation.

Acknowledgement: The authors are grateful to the participants and their relatives. We also thank the staff at the recruitment center (Þjónustumiðstöð Rannsóknarverkefna), the Memory Clinic at the National Hospital of Iceland, and the deCODE Genetics core facilities.

**HUSK**

The Norwegian Hordaland Health Study (HUSK) (36,000) is a population-based cohort study from the western part of Norway[150]. The first participants (n=18,000) were included in 1992-93 (born in 1925-27 and 1950-52); 18,000 additional participants were recruited in 1997 (born 1953-57). The Norwegian HUSK AD cohort (N=624, male=214, female=410, mean yob=1931.6 (SD 10.6) and controls (N=15,329, male=7,240, female=8,089, mean yob=1944.2 (SD 11.0) were obtained from the HUSK Study (REC#2018/915). The cases (ICD-10 codes) were identified using the Norwegian Patient Registry (NPR) in 2024. The mean age at onset was 80.2 years with a standard deviation of 10.2 years[147]. The sample was genotyped using Illumina Global Screening Array (version 3) at deCODE, Iceland. For the Norwegian HUSK AD GWAS, the correction factor based on the genomic control was 1.03.

**Lifelines**

The Lifelines cohort is a collection of health-related data, measurements, and biological samples from a population cohort. The cohort consists of over 167,000 individuals, including children, adults, and elderly people. Lifelines participants were initially recruited through general practitioners in Groningen, Friesland, and Drenthe. Invited participants were patients between 25 and 50 years old; these individuals were then asked to invite their family members (parents, partner, children, parents-in-law). Approximately 21k additional individuals registered directly for baseline participation via the Lifelines website. At baseline, 9% of participants (~15k) were 9 years or younger, 84% (~140k) were between 18 and 64, and 7% (~12k) were 65+. Approximately 58% of participants are female and 42% are male. The vast majority were born in the Netherlands (97%), and 98% are of European ancestry. The Lifelines protocol was approved by the UMCG Medical Ethics Committee under number 2007/152. All participants signed an informed consent form. Additional cohort Information is available here https://wiki.lifelines.nl/doku.php and https://wiki.lifelines.nl/doku.php?id=cohort. We accessed the data through project ov20_0073.

Maternal and paternal dementia proxy phenotypes were defined using self-reported questionnaire data from the baseline (2007–2013) and follow-up (2011–2014) assessments. Phenotypes were based on responses indicating dementia in the mother or father from fields dementia_mother_fam_q_1_a/b and dementia_father_fam_q_1_a/b. This yielded 8,401 maternal cases versus 116,784 controls, and 5,070 paternal cases versus 120,115 controls. Covariates included sex and age (from follow-up), 20 ancestry-specific genetic principal components (PCs), APOE ε4 allele count (estimated from rs429358 genotypes), and estimated parental age at death. Parental age was derived from reported birth and death years. Birth years were inferred by subtracting the participant's age from 2015. Implausible values (<16 or >60 years difference from offspring) or inconsistencies across assessments were set to missing. Death years were capped between 1900 and 2015; for living parents, 2015 was used. Ages were calculated as the difference between death and birth years and rounded to the nearest integer.

Three genotyping batches were analyzed separately: the GWAS cohort (CytoSNP-12v2 array), UGLI1 (GSA MultiEthnic Disease array), and UGLI2 (FinnGen Thermo Fisher Axiom array).

For the GWAS cohort, variants with call rate <0.95, MAF <0.01, or HWE P $<1\times10^{-3}$ were removed. Samples with call rate <0.80 or sex mismatches were excluded. Imputation was performed using SHAPEIT2 and Minimac with GoNL and 1000 Genomes reference panels. Full details are available at Lifelines GWAS QC.

For UGLI1, duplicate and tri-allelic variants, indels, sex-discordant samples, and those with high homozygosity were removed. Variant and sample call rate thresholds were 99%; variants with MAF <0.01 or HWE P $<1\times10^{-6}$ were excluded. Imputation was conducted through the Sanger imputation service using the Haplotype Reference Consortium panel. Details are available at UGLI1 QC.

UGLI2 followed a similar protocol with some differences: MAF threshold was 0.02 and HWE threshold was P $<1\times10^{-10}$. Pre-imputation alignment used HRC tools, and imputation was also conducted through the Sanger service using the HRC panel. Full details are available at UGLI2 QC.

Ancestry and relatedness were inferred after additional QC: removal of low-call rate variants and samples, strand ambiguous or multi-allelic variants, and variants in the MHC (chr6:25–35 Mb) or chr8 inversion (chr8:7–13 Mb). After pruning with PLINK (--indep-pairwise 200 100 0.2), samples were merged with the 1000 Genomes dataset. Individuals more than 4 standard deviations from the European mean along PC1 or PC2 were removed. Related individuals were removed using a KING cutoff of 0.177 within each batch (via PLINK v2.0) and across batches (via KING 2.2). Further details are provided in the UGLI QC report.

We performed three regression models per phenotype and dataset using different covariates:

1. Sex + Age + Age² + ParentalAge + ParentalAge² + APOE ε4 + PCs1–20
2. Sex + Age + Age² + ParentalAge + ParentalAge² + PCs1–20
3. Sex + PCs1–20

This resulted in 18 total analyses (3 models × 2 phenotypes × 3 datasets). Regressions were run using *regenie* v3.4 in a two-step procedure. Step 1 used covariates and genetic relationships to generate phenotype predictions based on cleaned genotype data (removing variants with MAF <0.01, MAC <100, genotyping rate <0.99, or HWE P $<1\times10^{-15}$). Step 2 tested association between imputed genotypes and phenotypes, adjusting for Step 1 predictions. We used BGEN files for autosomal analysis in the GWAS and UGLI2 cohorts. For UGLI1, we converted VCFs to BGEN using qctool v2.2.0 with 8-bit encoding. For the X chromosome in UGLI datasets, VCFs were converted to PLINK pgen format using hard-call thresholds (male: 0/2, female: 0/1/2). Only variants with INFO ≥0.3 and MAC >5 were included; post-analysis filters excluded variants with MAF <0.005 and X chromosome variants with INFO <0.8.

Acknowledgements: The Lifelines Biobank initiative has been made possible by funding from the Dutch Ministry of Health, Welfare and Sport, the Dutch Ministry of Economic Affairs, the University Medical Center Groningen (UMCG, the Netherlands), the University of Groningen, and the Northern Provinces of the Netherlands. The generation and management of GWAS genotype data for the Lifelines Cohort Study is supported by the UMCG Genetics Lifelines Initiative (UGLI). UGLI is partly supported by a Spinoza Grant from NWO, awarded to Cisca Wijmenga. The authors wish to acknowledge the services of the Lifelines Cohort Study, the contributing research centers delivering data to Lifelines, and all the study participants.

**Eli Lilly, Johnson & Johnson, and PROTECT**

The following three samples were combined for a GWAS, as Eli Lilly, Johnson & Johnson only contained cases. The PROTECT study contributed controls. The samples from the 3 datasets were matched based on principal components before being imputed together to minimize risk of false positives due to differing genotyping platforms. After pre-imputation QC, each of the 3 datasets were restricted to overlapping variants, Then, samples across the 3 datasets were matched based on the first two principal components, with any outliers being removed. Finally, the matched sample were merged before being imputed together.

Eli Lilly

This dataset includes 3,944 AD cases: 2,140 females and 1,804 males. Participants were aged 53–93 years (mean = 73) and recruited from 11 countries, primarily in Western Europe, with data collection taking place prior to 2017. Cases were defined as individuals aged ≥55 years with probable AD, meeting NINCDS/ADRDA diagnostic criteria. Additional inclusion criteria included a Modified Hachinski Ischemia Scale (MHIS) score ≤4, a Mini-Mental State Examination (MMSE) score between 16 and 26, and a Geriatric Depression Scale (GDS) short-form score ≤6. Participants also underwent either MRI or CT imaging within the past two years, with no findings inconsistent with AD. Approximately 50% of individuals had supporting evidence of amyloid pathology via florbetapir PET or cerebrospinal fluid (CSF) analysis. Individuals with major or unstable illnesses or requiring prohibited medications were excluded, as outlined in the study protocol. Participants were genotyped in four batches: three batches using the Illumina HumanOmni5M version 3 array and one batch (~50% of samples) using the Illumina Global Screening Array + Multi-disease panel version 2. Genome-wide association analyses were performed using PLINK v2.00a6LM AVX2 AMD (9 Jun 2024) with logistic regression. Covariates in the primary model included sex, age, age$^2$, age × sex, and the first 20 ancestry-specific principal components. Alternative covariate sets were used in additional models as described in the protocol. Variants with minor allele frequency <0.5% were excluded. Related individuals were removed such that all pairwise kinship coefficients were <0.05. Genetic ancestry was assessed using self-reported data, with outliers removed based on principal component analysis (PC1 vs. PC2).

Acknowledgements

Participants and care-givers for participating in AD clinical trials

Johnson & Johnson

This dataset includes 1,052 AD cases: 688 females and 364 males. Participants were aged 45–90 years (mean = 71) and recruited from 13 countries, with approximately 60% from Eastern Europe, 30% from Western Europe, and 10% from Southern Europe. Data collection occurred prior to 2022. Cases were defined as outpatients diagnosed with mild to moderately-severe Alzheimer's disease (probable or possible), based on criteria from the National Institute of Neurological and Communicative Disorders and Stroke and Alzheimer's Disease Related Disorders Association (NINCDS-ADRDA) or the DSM-IV. Exclusion criteria were applied as described in the study protocol. Genotyping was performed using the GDAneuroBooster array. GWAS was conducted jointly with the Eli Lilly dataset using the same protocol. Logistic regression was performed in PLINK v2.00a6LM with sex, age, $age^2$, age × sex, and the first 20 genetic principal components as covariates. Related individuals were excluded (kinship coefficient < 0.05), and variants with MAF <0.5% were removed. Genetic ancestry outliers were excluded based on principal component analysis (PC1 vs. PC2).

PROTECT

This dataset comprises 4,792 cognitively normal controls, consisting of 3,389 females and 1,403 males. Participants were aged 50–105 years (mean = 63) and were recruited in the UK between 2015 and 2022 as part of the PROTECT study (https://www.protectstudy.org.uk/). Cognitive status was assessed through annual computerized cognitive testing. Controls were defined as cognitively normal individuals. Exclusion criteria were applied as specified in the study protocol. Genotyping was performed using a customized version of the Illumina Global Screening Array v1. A GWAS was conducted jointly using the Eli Lilly and Johnson & Johnson datasets, employing the same analysis protocol. Logistic regression was performed in PLINK v2.00a6LM, including sex, age, $age^2$, age × sex, and the first 20 genetic principal components as covariates. Related individuals were excluded (kinship coefficient < 0.05), and variants with MAF <0.5% were removed. Genetic ancestry was assessed using self-reported data, with outliers excluded based on principal component analysis (PC1 vs. PC2).

Acknowledgment: The PROTECT Study is supported by the National Institute for Health and Care Research (NIHR) Exeter Biomedical Research Centre; an MRC Proximity to Discovery: Industry Engagement Fund (MC_PC_17189) grant awarded to Dr Creese; and by the NIHR Collaboration for Leadership in Applied Health Research and Care South West Peninsula. The views expressed are those of the author(s) and not necessarily those of the NIHR or the Department of Health and Social Care.

**Tohoku Medical Megabank (TMM)**

TMM is a population-based prospective cohort that enrolled participants from Miyagi and Iwate Prefectures in the Tohoku region, in the northeastern part of Japan[151]. In this study, we conducted analyses on participants recruited between 2013 and 2017. Genotyping of study participants was conducted using a custom SNP array for the Japanese population (that is, Japonica Array v.2[152]). For genotype quality control, we excluded the variants meeting any of the following criteria: call rate of <99%, P value for Hardy–Weinberg equilibrium of $<1.0 \times 10^{-5}$ and MAF < 0.01. The quality-controlled genotype data were pre-phased by using SHAPEIT2 software and imputed using IMPUTE4 software with a combined reference panel of 1000 Genomes Project phase 3 version 5 genotype data (n = 2,504) and Japanese whole-genome sequencing data (3.5KJPNv2; n = 3,552). After imputation, variants with an imputation INFO Score of <0.7 were excluded. Quality control of the study participants was performed with the following exclusion criteria: (1) outliers from East Asian ancestry clustering based on the projection PCA with samples of 1000 Genomes Project Phase 3 data; (2) genotype call rate of <95%; (3) individuals under 40 years of age; (4) without phenotype or covariate information.

A proxy phenotype is defined based on self-reported family history of dementia in questionnaires. Of the 60,363 participants, 421 who reported dementia in both parents, 1,525 in only their father, 4,499 in only their mother, and 53,918 control participants reported no parental dementia. We assigned half of the controls and participants who reported dementia in both parents to the paternal proxy and half to the maternal proxy. The total age range is between 40-93, the mean age of the maternal proxy-cases is 62.99, that of their controls is 61.75, and that of the paternal proxy-cases is 59.98, and that of their controls is 61.80. There was no information about parental age in the TMM cohort. We conducted proxy-GWAS for dementia using REGENIE, with covariates including age, $age^2$, sex, genotyping batch, and the top 20 principal components. Further details of the TMM cohort design and analyses of genotype and phenotype information are described elsewhere[151,153].

We acknowledge the participants and investigators of TMM. The TMM Project is supported by grants from the Reconstruction Agency, from the Ministry of Education, Culture, Sports, Science and Technology (MEXT), and the Japan Agency for Medical Research and Development (AMED) [JP20km0124005 and JP20km0424601].

**BioBank Japan (BBJ)**

BBJ is a patient-based cohort mainly comprising participants of Japanese ancestry from cooperating hospitals across Japan[154]. BBJ recruited approximately 200,000 individuals with at least one of 47 target diseases from twelve Japanese medical institutions and collected DNA, serum samples, and clinical information between 2003 and 2007. Genotyping of BBJ participants was conducted using the Illumina HumanOmniExpressExome BeadChip or a combination of the Illumina HumanOmniExpress and HumanExome BeadChips. A summary of genotyping platforms and quality control criteria for study participants and variants is described elsewhere[154,155]. Genotype dosages were imputed using minimac3 software with 1000 Genomes Project Phase 3 version 5 genotype data (n = 2,504) and Japanese whole-genome sequencing data (n = 1,037)[156]. After imputation, variants with an imputation quality of $R^2 < 0.3$ or minor allele frequency (MAF) < 1% were excluded.

The clinical status of Alzheimer's disease was defined based on both the doctor's diagnosis and medical records at the cooperating hospitals. We excluded participants under the age of 40 and outliers from East Asian ancestry clustering based on PCA. Of the 156,538 participants, 197 were male cases, 85,605 were male controls, 207 were female cases, and 70,529 were female controls. The age range is between 40-105, and the mean age of the male cases is 78.1, that of male controls is 67.9, that of female cases is 79.6, and that of female controls is 67.7.

Further details of the BBJ cohort design and analyses of genotype and phenotype information are described elsewhere[154,155].

We acknowledge the participants and investigators of BBJ that was supported by AMED. All participants provided written informed consent as approved by the ethical committees of the RIKEN Center for Integrative Medical Sciences and the Institute of Medical Sciences at the University of Tokyo.

**UK Biobank**

We created diagnosed and proxy AD phenotypes using ICD code information reported in field 41270 and illnesses of parent information (fields 20110 and 20107). First, any individual reporting the following ICD codes was considered a case: G30, F00, F03, and F06.7. Then, the age of onset information (fields p131036, p130836, p130842) was used to remove any cases with an age of onset <40. If age of onset information was not available, age at first assessment (field 21003 instance 0) was used instead. To maximize cases, the cases were not trimmed to unrelated individuals. This led to the inclusion of an additional 968 cases. To check whether these related cases were familial AD cases, we used the exome sequencing data to check the additional cases for pathogenic and likely pathogenic variants in *APP*, *PSEN1*, and *PSEN2* as defined by ALZFORUM (https://www.alzforum.org/mutations/). Only 1 of the 968 additional cases had the risk allele for one of these variants (*PSEN1* p.A246E). This suggests that the risk of these additional cases influencing the GWAS results towards familial AD was low.

Next, proxy maternal and paternal cases were defined by reporting an "Alzheimer's disease/dementia" diagnosis in fields 20110 and 20107, respectively. Any individuals included as diagnosed cases were removed from the proxy case list, along with any relatives with a kinship coefficient > 0.117 (defined by KING integrated in Plink v2). Next, maternal and paternal ages were calculated by taking the oldest reported age (<120) in fields 1845, 3526, 2946, and 1807. The cases were split into maternal and paternal cases, and individuals with the oldest reported age of < 40 were removed. The individuals who reported that both parents were cases were randomly split in half and assigned to either the maternal or paternal phenotype.

Controls for the diagnosed case sample were defined by any individual not reporting any of the following ICD codes or a parent diagnosed with "Alzheimer's disease/dementia": G30*, F00*, F03, F06.7 G20-G26, G31-32, G35-G37, F02, G10-14. Controls were selected using the R package MatchIt to identify the best matching controls based on age and sex, with age defined as the maximum value reported in field 21003 and sex defined in field 31. Four controls were selected for every case. Finally, proxy controls were defined as individuals who did not report that their parents had "Alzheimer's disease/dementia" and who did not report "Do not know". All individuals related to or overlapping with the diagnosed cases, controls, and proxy cases were removed. Any proxy controls reporting their parents' oldest age as <16 or >120 were removed. Then, half of the proxy controls were randomly split in half and assigned to the maternal and paternal phenotypes. This resulted in three phenotypes: one case-control analysis using diagnosed cases where related individuals were included; one maternal phenotype where no individuals were related to each other or to any other individuals included in the case-control or paternal phenotype; similarly, for the paternal phenotype. Phenotypes were created using R v4.2.1-foss-2022a.

Covariates were also created to be used in regression analyses. Age, age$^2$, maternal age, maternal age$^2$, paternal age, paternal age$^2$, sex, array01, PCs1-20, APOE4 dosage, and age*sex were used as covariates in at least one of the analyses. Age was defined as the max value reported in field 21003, age$^2$ was the square of that value, maternal age was defined as the oldest reported age (<120) in fields 2946, and 1807, paternal age$^2$ was the square of that value, paternal age was defined as the oldest reported age (<120) in fields 1845 and 3526, maternal age$^2$ was the square of that value, sex was defined in field 31, array01 was created by setting all values of field 22000 >1 as 1, APOE4 dosage was defined by the number C alleles reported for rs429358, age*sex was defined by the multiplication of centered age (scale(age, center=T,scale=F) in R) multiplied by sex, and PCs1-20 were defined within ancestry using FlashPCA2. Phenotypes were created using R v4.2.1-foss-2022a. Counts of the C allele of rs429358 were calculated using Plink v1.9 (--recode A).

We used genetic data from 436,803 UKB participants of European, South Asian, and African ancestry. These genotypes were used to empirically assign individuals to ancestral continental populations using the 1000 Genomes reference panel. Only these three ancestries had sufficient (proxy) cases to meet our analysis criteria (100 cases or 200 proxy cases). The UKB is a large population-based biobank that includes 503,325 individuals. Individuals were selected for participation between 2006 and 2010. Invited individuals were between 40 and 69 years old, registered with the National Health Service, and living within 25 miles of one of the study research centres. Various data were collected from the individuals, including questionnaire answers, medical records, and genetic data. Of the 436,803 participants included in this analysis, 241,878were female (55.4%) and 194,925were male (44.6%). The median oldest age reported in field 21003 was 59. All participants provided written informed consent. The UKB received ethical approval from the National Research Ethics Service Committee North West-Haydock (reference 11/NW/0382). All study procedures were in accordance with the World Medical Association for medical research. Access to the UK Biobank data was obtained under application number 16406.

We performed 11 regression analyses for each ancestry group using different phenotypes and covariates:

1) AD_status~ Sex+array+Age+Age$^2$+Age*Sex+*APOE*e4+PCs1-20
2) AD_status~ Sex+array+Age+Age$^2$+Age*Sex+PCs1-20
3) AD_status_Females~ array+Age+Age$^2$ +PCs1-20

4) AD_status_Males~ array+Age+Age$^2$ +PCs1-20
5) AD_status~ Sex+array +PCs1-20
6) Maternal_AD_status~Sex+array+Age+Age$^2$+MotherAge+MotherAge$^2$+ *APOE*e4+PCs1-20
7) Maternal_AD_status~Sex+array+Age+Age$^2$+MotherAge+MotherAge$^2$+PCs1-20
8) Maternal_AD_status~Sex+array+PCs1-20
9) Paternal_AD_status~Sex+array+Age+Age$^2$+FatherAge+FatherAge$^2$+ *APOE*e4+PCs1-20
10) Paternal_AD_status~Sex+array+Age+Age$^2$+FatherAge+FatherAge$^2$+PCs1-20
11) Paternal_AD_status~Sex+array+PCs1-20

This resulted in a total of 33 analyses. We performed the regression using regenie v3.1.3, with the standard two-step procedure. Step 1 created predictions based on the covariates and the genetic relationships between individuals based on genotype data described above, after additional variant QC procedures (removal of variants with MAF<0.01, MAC<100, genotyping rate <0.99, and Hardy-Weinberg equilibrium P-values <1x10$^{-15}$). Step 2 performed the regression analysis between the variants and phenotypes, adjusting for the predictions from Step 1. We used plink hard call files as input for Step 2. Firth correction was used for variants with P-values less than 0.01 (--firth --approx --pThresh 0.01). Only variants with an INFO score of 0.9 or larger were included and only variants with a MAC>5 were included. After analysis, variants with MAF<0.005 were removed.

**DemGene**

The DemGene cohort was recruited from a network of clinical sites collecting cases from Norwegian memory clinics, hospitals and nursing homes based on standardized examination of cognitive, functional, and behavioral measures and data on progression of real-world patients in Norway[157,158]. For the current study we included 1,449 cases and 2,328 controls from the Norwegian Register of persons with Cognitive Symptoms (NorCog[159]), the Progression of Alzheimer's Disease and Resource use (PADR[160]), the Dementia Study of Western Norway (DemVest[161]), the AHUS study, the Dementia Disease Initiation study (DDI[162]), the Dementia Study in Rural Northern Norway (NordNorge[163]), the Nursing Home study[164], the TrønderBrain study[165], and the Oslo Parkinson's Disease study[166]. These Norwegian cases were diagnosed according to the recommendations from the National Institute on Aging–Alzheimer's Association (NIA/AA) (AHUS, DDI), the NINCDS-ADRDA criteria (DemVest and TrønderBrain), or the ICD-10 research criteria (NorCog, PADR, NordNorge). The current sample also included European cohorts: Brno, Strasbourg, Stockholm, Halle. The controls were screened with standardized interviews and cognitive tests. To increase the statistical power of our association analysis, the controls were combined with additional population controls from Norwegian blood donor samples (Oslo University Hospital, Oslo) and controls from the Thematically Organized Psychosis (TOP) Study (between 25-65 years). Controls of the TOP Study were of Caucasian origin without a history of moderate/severe head injury, neurological disorder, mental retardation, and were excluded if they or any of their close relatives had a lifetime history of a severe psychiatric disorder, a history of medical problems thought to interfere with brain function, or significant illicit drug use.

DemGene participants were genotyped at deCODE Genetics using customized versions of Illumina GSA v1 and v3 arrays, as well as the Human Omni Express-24 v1.1 array (Illumina Inc., San Diego, CA, USA). The Halle cohort was genotyped using Affy 6.0, Illumina HumanHap 300, Illumina Omni1-Quad, and Illumina Human OmniExpress 12. The regression included batch as a covariate, and each batch was genotyped on a single array.
The current study was approved by REC# 2014/631.

Acknowledgements: We thank all patients participated in DemGene, and clinicians involved in the study and data collection. We gratefully acknowledge support from the Research Council of Norway AgeCare, RCN#344121, PreciMENT Nordforsk #164218, MultiMENT RCN#324499, NIMH Award 1R01MH124839.

**VUMC**

Participants were drawn from 9 studies, including 2,965 from the Amsterdam Dementia Cohort (ADC); 799 from the Parelsnoer initiative (PSI) and university medical hospitals in the Netherlands (395 from Erasmus Medical center; 9 from LUMC; 40 from the Amsterdam UMC location AMC; 157 from MUMC; 67 from Radboudmc; 84 from UMC Utrecht and 47 from UMCG)[167]; 97 from Twin 60 ++ project[168]; 69 from the 90+ Study; 478 from Dutch Brain Bank[169]; 48 from PROGRESS PD[170]; 1,654 from the Longitudinal Aging Study Amsterdam (LASA), 188 partners of participants of the 100-Plus Study[171]; 74 from the European Medical Information Framework (EMIF) project[172]; Details on the different studies can be found in their respective references. Probable AD was defined according to NINCDS–ADRDA/NIA–AA criteria, supplemented with biomarker evidence when available for the ADC, or based on the neuropathological diagnosis of AD[173,174]. Control subjects were participants cognitively normal at presentation (PSI, Progress PD, Twin 60 ++ project, LASA, 100-Plus Study), no clear neuropathology (NBB) or with subjective cognitive decline (ADC)[175], the latter means patients presented with cognitive problems, but performed within normal limits in neuropsychological assessments. All samples were prepared and analyzed at a single site, and in total, we included 6,848 European-ancestry participants collected from 1992 to 2023 for the current study: 2,812 AD cases and 4,036 controls (52.5% female). The cases had a mean age of 69.6 years (range: 32–100 years), while the controls had a mean age of 62.6 years (range: 18.2–102 years).

All the participants and/or their legal guardians gave written informed consent for participation in the clinical and genetic studies. DNA was extracted following standard operating procedures; thereafter, all DNA samples passing standard quality control were genotyped on the Illumina Infinium Global Screening Array (GSA) v1.0, v1.0 (with custom content), or v3.0. Variant- and sample-level QC was applied within each array, and datasets were combined at the genotype level. Details are described in Tesi et al., (2024)[96]. We then imputed the combined dataset with the TOPMed reference panel. Outliers based on principal-component analysis and one member of each related pair were excluded to ensure population homogeneity and sample independence. Association testing was performed in PLINK 2.0, adjusting for age, age$^2$, age × sex, sex, and the first 20 principal components, with additional sex-stratified and sensitivity analyses (i.e., excluding age as a covariate and controlling for APOE genotype).

##### Penn Medicine Biobank (PMBB), Mount Sinai Million Health Discoveries Program, INDIANA-CHALASANI, UCLA, Mayo Clinic-RGC Project Generation, Colorado Center for Personalized Medicine – RGC Collaboration, & GHS-RGC DiscovEHR collaboration

Three cohorts, PMBB, Mount Sinai Million Health Discoveries Program and INDIANA-CHALASANI were genotyped using the illumina Global Screening Array genotyping chip. GHS participants genotyped using either the Illumina Infinium OmniExpressExome or the Global Screening Array. The remaining cohorts, UCLA, MAYO-CLINIC and COLORADO were genotyped using targeted genomic sequencing using the twist diversity SNP panel, followed by the multipoint refinement using GLIMPSE[176]. Within each cohort, standard quality-control procedures (see below) were followed to retain only high-quality genotyped variants (see below), which were then used for imputing common variants using the TOPMed[177] LD reference panel.

Stringent and standardized pre-imputation quality control was performed for each cohort. Briefly, the following filters and checks were implemented including, removal of variants with missingness > 0.01; removal of samples with missingness >0.1; removal variants with significant deviations from Hardy-Weinberg equilibrium (p<1e-15). For this study, post-imputation QC included retaining common variants (minimum MAF > 1%) and MaCH[178] $R^2$>0.1. For cohorts for which genotype by sequencing was used, a specialized multipoint refinement (GLIMPSE) based variant caller was used to increase the call rate and concordance in spite of differential coverage across sites.

Association analyses in each study were performed using the genome-wide Firth logistic regression test implemented in REGENIE[179] v3.2.8. In step 1 of REGENIE, we included the variants derived from TOPMed imputation with MAF>1%, 10% missingness, Hardy-Weinberg equilibrium test - and linkage disequilibrium (LD) pruning (1,000 variant windows, 100 variant sliding windows and ). The association model used in REGENIE included as covariates (1) age, age squared, sex, age-x-sex, and age squared-x-sex; (2) 10 ancestry-informative principal components (PCs) derived from the analysis of a stricter set of LD-pruned common variants from the imputed data; (3) for the analysis of GHS, we additionally included genotyping batch (4 batches) as covariates; (4) for the analysis of Mount Sinai Million Health Discoveries Program, we additionally included two covariates F_MISS and miss_bin, where F_MISS was the direct measure of missingness rates observed in imputation and miss_bin was binarization of the F_MISS variable at missingness = 0.0017; (5) for the analysis of UPENN-PMBB, we additionally included one batch indicator.

We used the broad Alzheimer’s Disease definition: cases were those who had ICD-10 G30 diagnosis, or dementia in AD (F00), or other primary non-demyelinating, non-vascular degenerative conditions (G310|G311|F00|F02|F03), and controls were those who had no neurodegenerative disorders (G20-G26, G30-G32, G35-G37).

Data availability:

**Geisinger Health System**

Regeneron can make individual-level genomic data available to qualified academic noncommercial researchers through the REGN pre-clinical Research portal at https://regeneron.envisionpharma.com/vt_regeneron/ under a data access agreement.

**PMBB, Mayo Clinic, CCPM, and UCLA**

Academic, non-commercial researchers interested in reproducing the results reported in this manuscript may request access to individual-level data via a data access agreement by reaching out to the associated biobank.

Acknowledgements:
We acknowledge the Penn Medicine BioBank (PMBB) for providing data and thank the patient-participants of Penn Medicine who consented to participate in this research program. We would also like to thank the Penn Medicine BioBank team and Regeneron Genetics Center for providing genetic variant data for analysis. The PMBB is approved under IRB protocol# 813913 and supported by Perelman School of Medicine at University of Pennsylvania, a gift from the Smilow family, and the National Center for Advancing Translational Sciences of the National Institutes of Health under CTSA award number UL1TR001878.
We would like to thank all patient-participants engaged in the MyCode Community Health Initiative and the MyCode Research Team. We would also like to acknowledge the members of the Geisinger-Regeneron DiscovEHR Collaboration who have been critical in the generation of the data used for this study.
Mayo Clinic-RGC Project Generation were supported in part by Mayo Clinic's Center for Individualized Medicine.
Biospecimens and associated data used in this study were obtained from the biobank at the Colorado Center for Personalized Medicine (CCPM) at the University of Colorado Anschutz Medical Campus (CU AMC). All samples and data were collected under Institutional Review Board (IRB) approved protocol (#15-0461) with appropriate informed consent from participants. Research using these materials was conducted in accordance with the ethical guidelines and regulations governing human subjects research, upholding the principles of respect for persons, beneficence, and justice.

**BioVU (Vanderbilt University Medical Center), Gothenburg H70 Birth Cohort Studies and Clinical AD from Sweden, Gr@ce, TwinGene, 23andMe Research Institute, STSA**

Detailed information for these cohorts can be found in Jansen et al (2019)[180] and Wightman et al (2021)[2].

Acknowledgement

We would like to thank the research participants and employees of 23andMe Research Institute for making this work possible

**HUNT**

The Trøndelag Health Study (HUNT) is a Norwegian population-based cohort study[181,182]. The HUNT study is still ongoing and currently consists of four surveys (HUNT1 [1984-1986], HUNT2 [1995-1997], HUNT3 [2006-2008], and HUNT4 [2017-2019]). All residents of North-Trøndelag aged 20 years or older were invited to participate in HUNT1-3, while all residents aged 20 or older from the former Trøndelag county were invited to take part in HUNT4. The data came from various sources, including self-reported information, health measurements, biological samples, and genotyping; more details can be found at the HUNT databank online https://hunt-db.medisin.ntnu.no/hunt-db/. Additionally, participant data were linked across different local and national registries, the HUNT database, and the HUNT biobank, using each participant's Norwegian Identification Number.

To date, 240,000 individuals have participated in at least one of the HUNT surveys and 88,000 have been genotyped[183]. Participants were genotyped using Illumina HumanCoreExome arrays (HumanCoreExome12 v1.0, HumanCoreExome12 v1.1, or UM HUNT Biobank v1.0). Participants were excluded if they had a call rate below 99%, contamination over 2.5% as estimated with BAF Regress, large chromosomal copy number variants, lower call rates in technical duplicate pairs or twins, uncommon sex chromosomal configurations (other than XX or XY), or discrepancies with reported gender. The remaining individuals were analyzed using Genome Studio following a standard quality control protocol https://www.illumina.com/techniques/microarrays/array-dataanalysis-experimental-design/genomestudio.html. The BLAT[184] tool was used to map the genomic position, strand orientation, and reference allele genotyped variants, using the Genome Reference Consortium Human genome build 37 http://genome.ucsc.edu. Additionally, variants were excluded if they had a call rate <99%, another assay identified a higher call rate, the probe sequences did not map to the reference genome, cluster separation <0.3, Gentrain score <0.15, or the Hardy-Weinberg equilibrium deviation from unrelated samples of European ancestry had a p-value <0.0001. To harmonize the three arrays, variants with frequency differences >15% between the datasets or variants monomorphic in one dataset and with MAF > 1% in one of the other datasets were removed. Ancestry was inferred using an online singular value decomposition and shrinkage adjustment algorithm (FRAPOSA)[185], projecting the genotype samples into the space of the principal components of the Human Genome Diversity Project (HGDP) reference panel (938 unrelated individuals; downloaded from http://csg.sph.umich.edu/chaolong/LASER/).

Eagle2 v2.3[186] was then used to phase the data, and imputation was performed with the Haplotype Reference Consortium (HRC) reference panel[187] using Minimac4 v1.0 software https://genome.sph.umich.edu/wiki/Minimac4.

Cases were defined using ICD-10 codes G30*, F00*, F03*. Controls were defined as not having ICD-10 codes G30*, F00*, F03*. Data about ICD-10 codes have been available since 1997. The age for cases was defined as the age at the first time the AD was diagnosed, while the age for controls was the age at the last time they were seen at one of the HUNT surveys. Additionally, we excluded cases with age at the first diagnosis <40 years and controls with age< 70 years. The analysis was restricted to individuals of European ancestry. Individuals without genotyping data or information about their age were excluded.

Scalable and Accurate Implementation of GEneralized mixed model (SAIGE) version 1.0.3[188] was used. Depending on the model, adjustment was performed by including sex, age, $age^2$, age-sex interaction, genotyping batch, the first 20 genetic principal components and/or APOE ε4 dosage (APOE status was defined using two dummy variables: 0[reference]/2 copies of the C allele at rs429358 and 0[reference]/1 copy of the C allele at rs429358). Additionally, variants with imputation INFO score < 0.3 and with minor allele count (MAC) <50 were excluded.

In total, 3,924 cases (age range [42.0-103.0]; age mean=81.1) and 19,410 controls (age range [70.1-103.3]; age mean=77.9) have been included in the main analysis. In the sex-specific analysis, 2,372 female cases (age range [42.0-103.0]; age mean=81.6) and 10,340 female controls (age range [70.1-103.3]; age mean=78.3), and 1,552 male cases (age range [44.0-101.0]; age mean=80.3) and 9,070 male controls (age range [70.1-101.1]; age mean=77.5) have been included.

Acknowledgements: The Trøndelag Health Study (HUNT) is a collaboration between HUNT Research Center (Faculty of Medicine and Health Sciences, NTNU, Norwegian University of Science and Technology), Trøndelag County Council, Central Norway Regional Health Authority, and the Norwegian Institute of Public Health. The genotyping in HUNT was financed by the National Institutes of Health, University of Michigan, the Research Council of Norway, the Liaison Committee for Education, Research and Innovation in Central Norway, and the Joint Research Committee between St Olavs hospital and the Faculty of Medicine and Health

Sciences, NTNU. The HUNT Center for Molecular and Clinical Epidemiology (formely the K.G. Jebsen Center for Genetic Epidemiology) was financed by Stiftelsen Kristian Gerhard Jebsen; Faculty of Medicine and Health Sciences, NTNU, Norway. We thank HUNT participants for donating their time, samples, and information to help others; clinicians and other employees at Nord-Trøndelag Hospital Trust for their support and for contributing to data collection

Ethics: The current study is approved by the Regional Committee for Medical and Health Research Ethics (ref. 2017/1031).

##### IGAP, Shigemizu et al. (2021)

The summary statistics from these cohorts are described in detailed in their respective publications (IGAP[189], Shigemizu[190]).

##### Million Veteran Program (MVP)

The MVP summary statistics used are available at dbGAP phs001672 and described in detail in Sherva et al (2022)[191]. ML was supported by MVP000 as well as MVP grant (MVP015/VA BLR&D I01BX004192) and the continuation grant (MVP040/BLR&D I01BX005749). This publication does not represent the views of the Department of Veteran Affairs or the United States Government.

##### FinnGen

We downloaded the GWAS summary statistics for Alzheimer's Disease (wide definition; G6_AD_WIDE) from the FinnGen Freeze 10 release on January 17, 2024. Details can be found here: https://risteys.finngen.fi/endpoints/G6_AD_WIDE, https://r10.finngen.fi/pheno/G6_AD_WIDE, and https://finngen.gitbook.io/documentation/methods/phewas. The FinnGen study is a large-scale genomics initiative that has analyzed over 500,000 Finnish biobank samples and correlated genetic variation with health data to understand disease mechanisms and predispositions. The project is a collaboration between research organizations and biobanks within Finland and international industry partners.

Acknowledgement
We want to acknowledge the participants and investigators of the FinnGen study.

**Estonian Biobank**

The Estonian Biobank (EstBB) is a large data-rich population-based biobank, covering approximately 20% of the adult population in Estonia (N=~210,000)[192]. All EstBB participants have signed an informed consent form and provided blood samples for genotyping. Electronic health records are regularly retrieved by linking to the national health databases and registries, such as the National Health Insurance Funds (NHIF) database, cause of death register, and hospital records.

Cases were defined as individuals with an ICD-10 diagnosis of F00* (Dementia in Alzheimer's disease) or G30* (Alzheimer’s disease). Individuals with an AD diagnosis before the age of 30 were excluded (n = 25). Controls were defined as individuals without any of the following ICD-10 diagnoses: F01* (Vascular dementia), F02* (Dementia in other diseases classified elsewhere), F03* (Unspecified dementia), F05.1 (Delirium superimposed on dementia), F10.6 (Amnesic syndrome), F10.73 (Residual and late-onset psychotic disorder [dementia]), G31* (Other degenerative diseases of the nervous system, not elsewhere classified), and I67.3 (Progressive vascular leukoencephalopathy).

The GWAS included 533 cases and 194,928 controls. The analysis was performed using SAIGE version 0.43.1, with gender, birth year, and the first 10 principal components included as covariates. Data analysis was carried out in part in the High-Performance Computing Center of the University of Tartu.

The activities of the EstBB are regulated by the Human Genes Research Act, which was adopted in 2000 specifically for the operations of the EstBB. Individual-level data analysis in the EstBB was carried out under ethical approval 1.1-12/624 from the Estonian Committee on Bioethics and Human Research (Estonian Ministry of Social Affairs) using data according to the release application 6-7/GI/18759 from the Estonian Biobank.

Acknowledgement: We acknowledge all the participants of the Estonian Biobank. The research was conducted using the Estonian Center of Genomics/Roadmap II funded by the Estonian Research Council (project number TT17). EstBB was funded by the Estonian Research Council grant PSG615 and Estonian Centre of Excellence for Well-Being Sciences, and by grant TK218 from the Estonian Ministry of Education and Research.

#### Copenhagen Hospital Biobank (CHB) and The Danish Blood Donor Study (DBDS)

Danish samples and data were obtained in collaboration with CHB [193] and DBDS [194]. CHB is a research biobank, which contains samples obtained during diagnostic procedures on hospitalized and outpatients in the Danish Capital Region hospitals. Data analysis within CHB was performed under the '"Developing the basis for personalized medicine in degenerative and episodic brain disorders' protocol, approved by the Regional Scientific Ethical Committee and the Capital Region Data Protection Office (H-21058057 and P-2022-392). DBDS is a nationwide study of ~170,000 blood donors, approved by The Danish Data Protection Agency (P-2019-99) and the National Committee on Health Research Ethics (NVK-1700407).

CHB and DBDS contributed a total of 12,031 Alzheimer's disease (AD) cases and 210,602 controls. The proportion of females was 52.1% in cases and 58.5% in controls. AD diagnoses were ascertained through the Danish National Patient Register using ICD-10 codes (G30, F00, F03), with age of onset ≥40 years. Controls were free of major neurological disorders (ICD-10 codes: F06.7, G20–26, G31–32, G35–37, G10–14). The mean age was 82.7 years (range 42–107) for cases and 64.0 years (range 17–109) for controls. Data collection began in 2009 at Copenhagen University Hospital – Rigshospitalet and in 2012 across other hospitals in the Capital Region of Denmark and is ongoing.

Genotyping was performed on the Illumina GSA chip, with imputation using the deCODE panel (≈50,000 individuals, including 10,800 Danes) on GRCh38. GWAS analyses were conducted in Regenie v3.3 with QC thresholds of MAF > 0.005 and INFO > 0.3, and covariates including sex, chip batch, age, $age^2$, age×sex, and PCs 1–20. Ancestry was assigned using FlashPCA 2.0; individuals within three standard deviations of the mean of those with both parents born in Denmark were retained.

Acknowledgement: We wish to express special gratitude to the individual participants in the Copenhagen Hospital Biobank. We also thank those who worked on generating these resources through data and sample collection, genotyping, and analysis.

Ethics: The study was approved by the National Scientific Ethical Committee and the Capital Region Data Protection Office (H-21058057 and P-2022-392)

Funding: Copenhagen Hospital Biobank (CHB) sample collection was facilitated by the infrastructure of Bio- and Genome Bank Denmark. Furthermore, CHB is supported by the Department of Clinical Immunology, Rigshospitalet, Copenhagen University Hospital, Copenhagen, Denmark and by grants from Novo Nordisk Foundation (NNF23OC0082015, NNF17OC0027594) and Rigshospitalet Research Council (Framework grant). The Danish Blood Donor Study (DBDS) is funded by an annual grant from Bio- and Genome Bank Denmark. The initiation of DBDS was supported by the Danish Administrative Regions (02/2611) and the Danish Council for Independent Research (09–069412). Additionally, the DBDS is funded by the Novo Nordisk Foundation (NNF23OC0082015, NNF17OC0027864, and NNF17OC0027594). The work was further funded by the Research Fund at Sygeforsikringen Danmark (2021-0245).

**Genentech**

This study comprised 2,211 cases (978 males, 1,233 females) and 11,726 controls (5,797 males, 5,929 females), with data collected globally between 2011 and 2020. Cases were clinically adjudicated within clinical trials and met the NINCDS-ADRDA criteria for Alzheimer's Disease, supported by biomarker evidence of amyloid pathology. Controls were selected from individuals enrolled in studies unrelated to neurological or neurodegenerative diseases. The age range for controls was 50-89 years (mean age: 60.6 years), while for cases, it was 50-90 years (mean age: 70.8 years).

Participants were genotyped either by array or whole genome sequencing. High-quality array genotype data were imputed using BEAGLE 5.1 with hg38 HapMap genetic maps and a Genentech reference panel consisting of over 25,000 individuals from diverse ancestries. Imputed datasets were merged with whole-genome sequencing data, focusing on variants present on both platforms. Genetic ancestry was estimated using ADMIXTURE 1.3 with reference samples from Phase 3 of the 1000 Genomes Project; analyses were restricted to individuals with a European ancestry assignment of >70%. Variants exhibiting differential missingness between cases and controls were excluded, and only those with an overall call rate exceeding 95% were retained. Genome-wide association analysis was conducted using REGENIE, adjusting for covariates including sex, age, age squared, age-by-sex interaction, principal components, study, and APOE genotype.

Acknowledgement: We thank our colleagues from the Genentech Department of Human Genetics, with special acknowledgement of Natalie Bowers and Vipin Menon for facilitating data access. We are also deeply grateful to the study teams, investigators, and the patients who generously contributed their samples and data for this research.

### Supplementary Figures

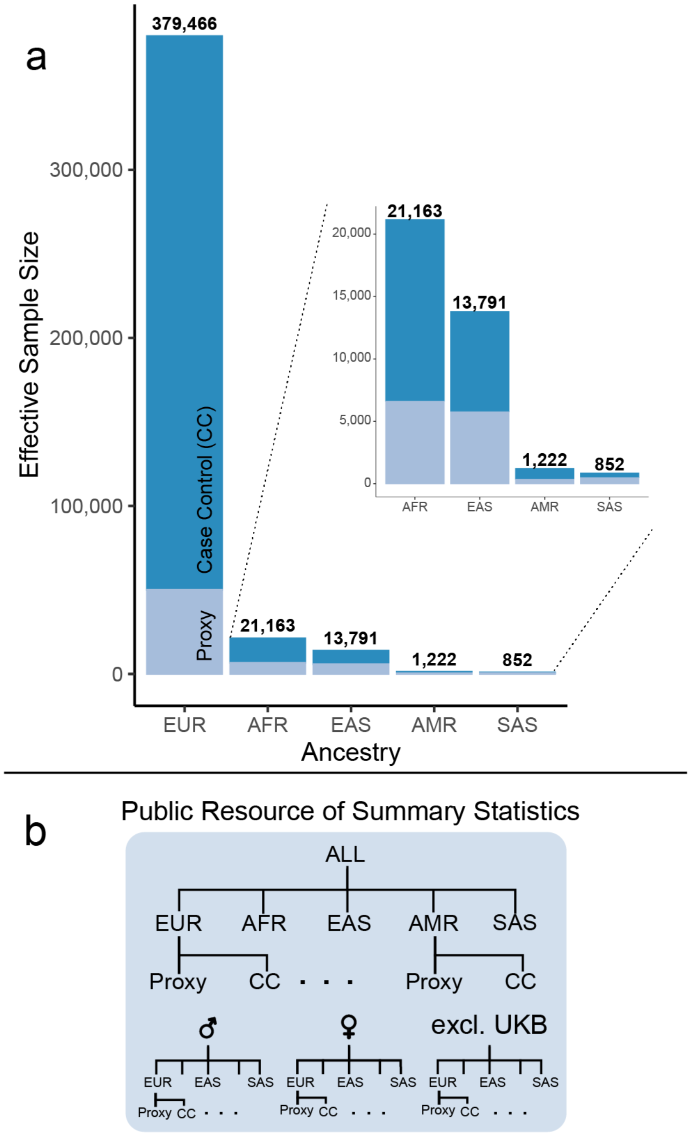

**Supplementary Figure 1. Sample size and sumstats. a,** Overview of the effective sample size for each ancestry and the proportion cases-control samples to proxy samples. **b**, Overview of the combined and stratified summary statistics that have been made publicly available.

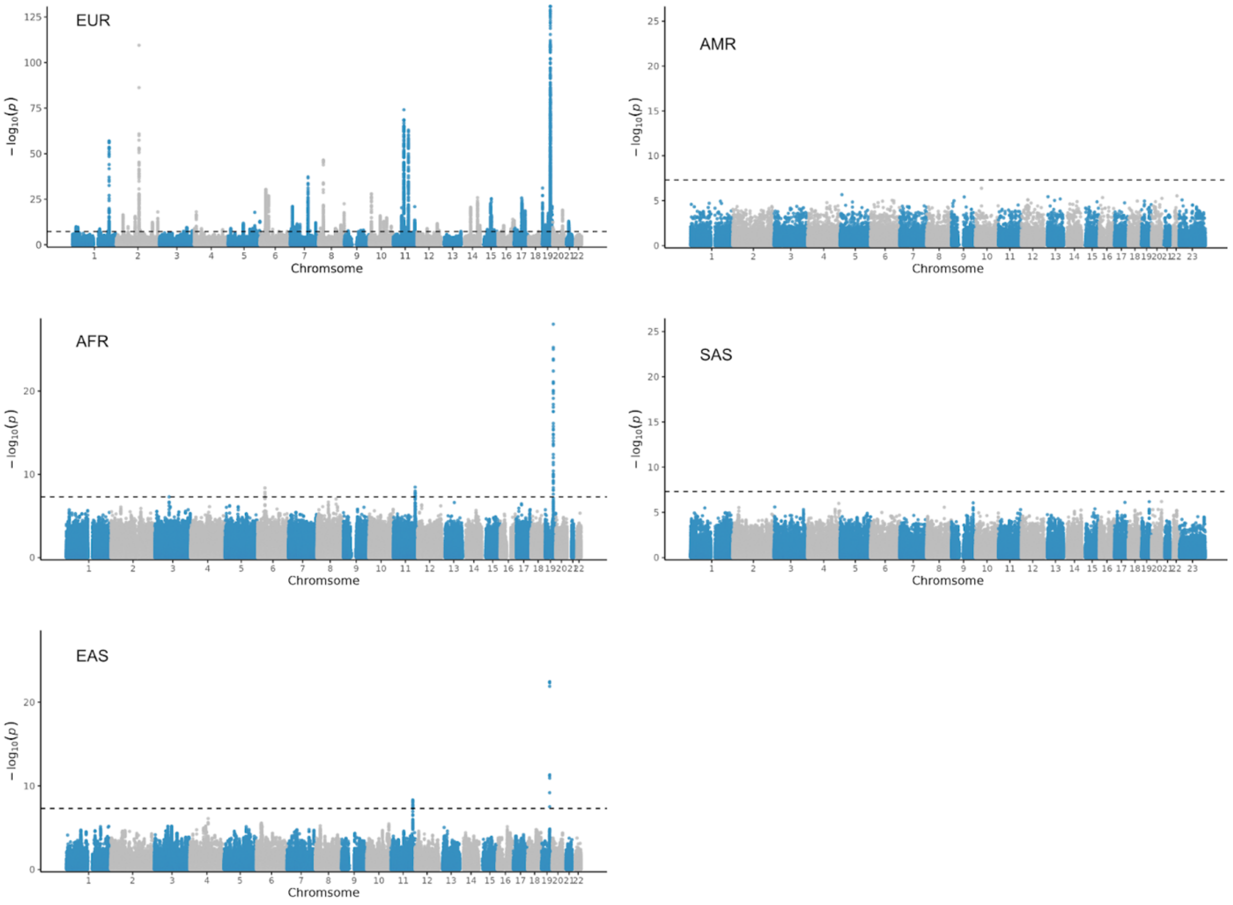

**Supplementary Figure 2. Manhattan plots of ancestry-specific GWAS.** The x-axis represents the chromosomal position of SNPs, and the y-axis their strength of association measured as $-\log_{10}(p)$. The dashed line represents genome-wide significant ($p$ < $5x10^{-8}$). The European y-axis is capped at 125 for visual clarity; however the APOE locus reaches $-\log_{10}(p)$ ~ 5000.

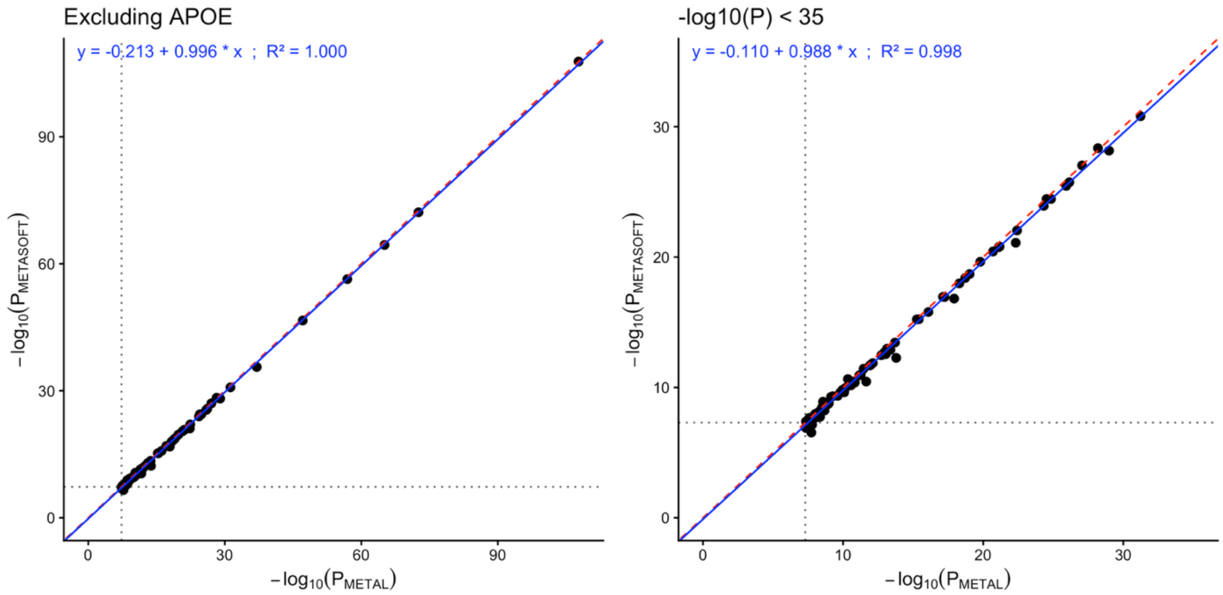

**Supplementary Figure 3. Comparison of METAL and METASOFT P-values.** Scatter plots of −log10(P) values from the cross-ancestry meta-analysis, comparing METAL (inverse-variance weighted fixed-effects) on the x-axis with METASOFT (Han-Eskin RE2 random-effects) on the y-axis. Each point represents the lead variant of a genome-wide significant locus from the main analysis, excluding the APOE locus (19:45411941:C:T). The left panel shows all non-APOE lead variants; the right panel zooms in on variants with −log10(P) < 35. The dashed red line is the identity line (y = x), the solid blue line is the linear regression fit, and the gray dotted lines mark the genome-wide significance threshold ($P = 5\times10^{-8}$).

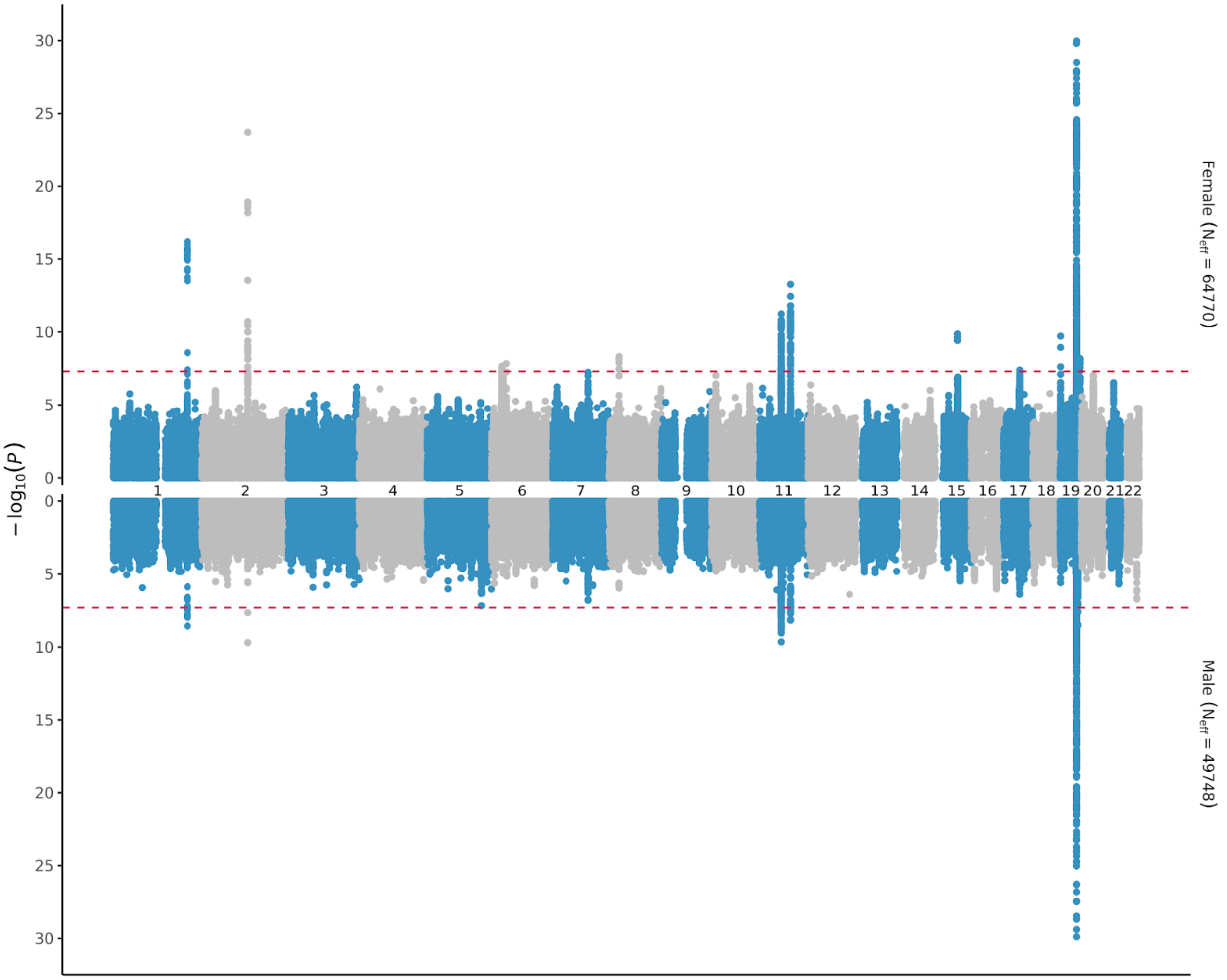

**Supplementary Figure 4. Miami plot of sex-stratified GWAS.** The x-axis represents the chromosomal position of SNPs, and the y-axis their strength of association measured as $-\log_{10}(p)$. The dashed line represents genome-wide significant ($p < 5x10^{-8}$). The y-axis was capped at 30.

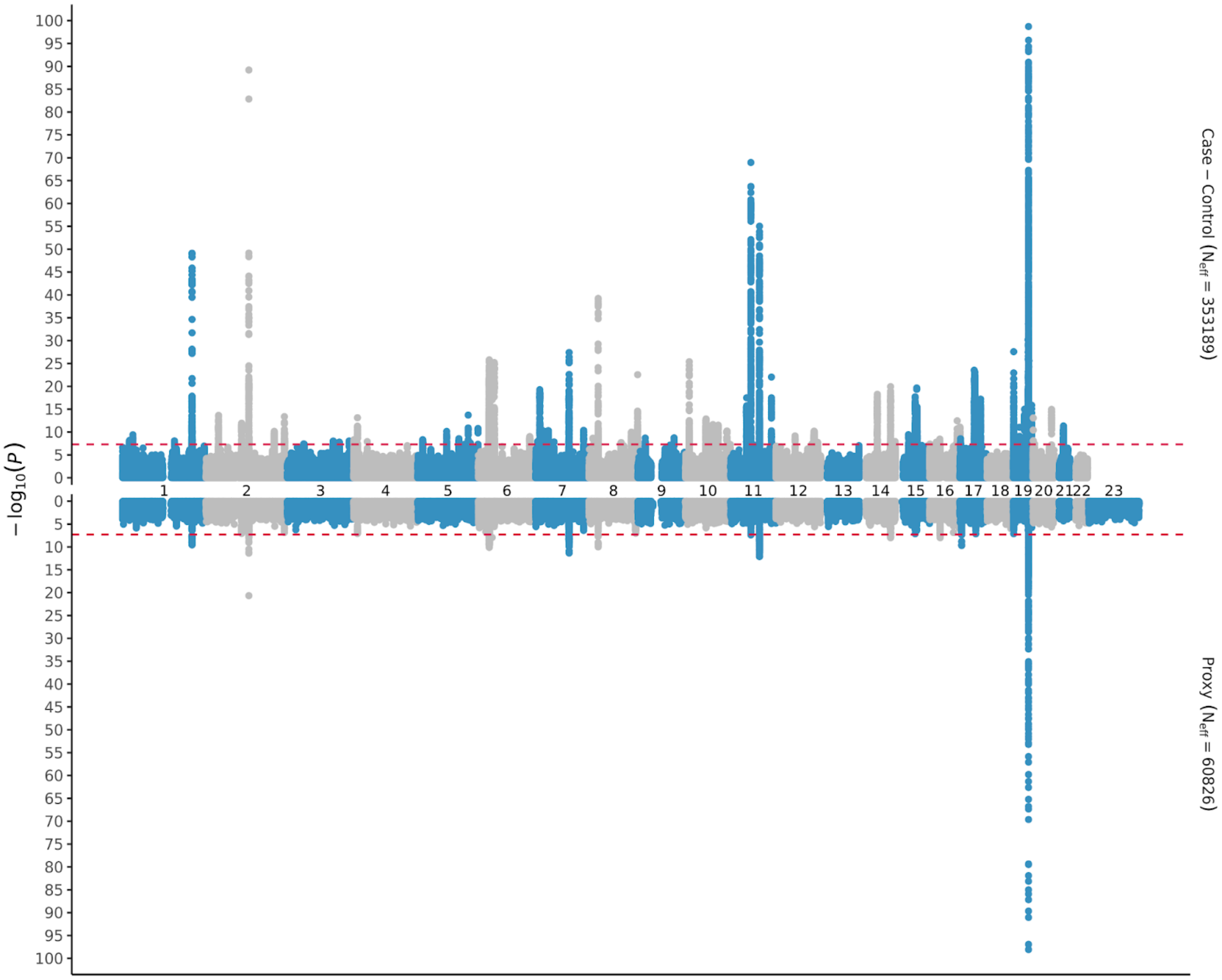

**Supplementary Figure 5. Miami plot of phenotype-stratified GWAS.** The x-axis represents the chromosomal position of SNPs, and the y-axis their strength of association measured as $-\log_{10}(p)$. The dashed line represents genome-wide significant ($p$ < 5x10$^{-8}$). The y-axis was capped at 100.

| Locus | All_Combined (414,019) | All_noProxy (353,193) | All_Proxy (60,826) | EUR_Combined (378,916) | EUR_noProxy (329,373) | EUR_Proxy (49,543) | AFR_Combined (19,354) | AFR_noProxy (14,555) | AFR_Proxy (4799) | AMR_Combined (1217) | AMR_noProxy (863) | AMR_Proxy (354) | EAS_Combined (13,681) | EAS_noProxy (8034) | EAS_Proxy (5647) | SAS_Combined (851) | SAS_noProxy (368) | SAS_Proxy (483) | Male (72,196) | Female (105,300) |
|---|---|---|---|---|---|---|---|---|---|---|---|---|---|---|---|---|---|---|---|---|
| Locus 1 HP1BP3/.. | 2.17e-11 | 6.73e-09 | 6.65e-05 | 1.22e-10 | 1.92e-08 | 0.000495 | 0.000527 | 0.000434 | 0.000932 | 0.0288 | 0.00845 | 0.000119 | 0.00138 | 0.00041 | 0.000296 | 0.00243 | 0.00759 | 0.00254 | 0.000201 | 0.000421 |
| Locus 2 LAMPTM5 | 5.48e-10 | 3.96e-10 | 0.000339 | 1.9e-10 | 2.66e-10 | 0.00035 | 0.00543 | 0.00266 | 0.000515 | 0.0014 | 0.0075 | 0.0126 | 0.0112 | 0.00848 | 0.00237 | 0.00035 | 0.00175 | 0.00111 | 5.09e-05 | 8.41e-05 |
| Locus 3 PLEKHO1 | 2.09e-08 | 1.97e-06 | 0.000632 | 3.98e-08 | 2.63e-06 | 0.00153 | 0.00133 | 0.00296 | 0.00896 | 0.00857 | 0.0205 | 0.0687 | 0.0447 | 0.118 | 0.00316 | 0.00602 | 0.0143 | 0.0159 | 0.000708 | 0.00249 |
| Locus 6 KIF21B/.. | 4.09e-08 | 7.88e-07 | 0.00272 | 3.67e-08 | 4.02e-07 | 0.00245 | 0.00124 | 0.000698 | 0.000329 | 0.00875 | 0.00277 | 0.000396 | 0.0163 | 0.00229 | 0.0024 | 0.00131 | 0.000346 | 0.0105 | 0.00296 | 4.88e-05 |
| Locus 15 ERC2 | 3.5e-08 | 3.18e-06 | 0.000201 | 3.37e-07 | 3.32e-05 | 0.000194 | 1.65e-05 | 0.000164 | 0.000431 | 0.0112 | 0.00215 | 0.00388 | 0.00379 | 0.00112 | 0.000543 | 0.0276 | 0.00292 | 0.00776 | 0.00249 | 0.00556 |
| Locus 16 NCK1 | 1.11e-08 | 8.97e-09 | 0.000223 | 4.31e-08 | 4.4e-08 | 0.000143 | 0.000225 | 0.000286 | 0.00104 | 0.00292 | 0.0158 | 0.00171 | 0.00446 | 0.00529 | 0.000593 | 0.00611 | 0.018 | 0.00387 | 0.00164 | 0.00126 |
| Locus 17 MBNL1 | 1.09e-08 | 2.59e-08 | 0.00177 | 1.62e-06 | 1.66e-06 | 0.00187 | 0.00291 | 0.00846 | 0.000741 | 0.00403 | 0.00502 | 0.000162 | 0.000487 | 0.000625 | 0.0125 | 0.0125 | 0.0116 | 0.00378 | 0.000692 | 0.00194 |
| Locus 20 MAP3K13 | 1.75e-07 | 2.93e-06 | 0.00559 | 2.21e-08 | 5.32e-07 | 0.00573 | 0.00342 | 0.00236 | 0.00272 | 0.0047 | 0.0349 | 0.000774 | 0.0301 | 0.0595 | 0.00371 | 0.0108 | 0.0122 | 0.00491 | 0.00227 | 6.44e-05 |
| Locus 21 UBXN7/.. | 4.45e-09 | 6.13e-08 | 0.00209 | 4.26e-09 | 4.05e-08 | 0.00701 | 0.000212 | 0.000655 | 4.72e-05 | 0.00457 | 0.0105 | 0.00432 | 0.00183 | 0.0127 | 0.00194 | 0.000933 | 0.00174 | 0.00366 | 0.000698 | 5.47e-05 |
| Locus 25 RAPGEF2 | 3.45e-09 | 9.78e-08 | 3e-04 | 4.36e-09 | 1.95e-07 | 0.000226 | 0.0017 | 0.00139 | 0.00149 | 0.00906 | 0.0204 | 4.83e-05 | 0.00709 | 0.00886 | 0.00248 | 0.00472 | 0.017 | 0.0076 | 0.00803 | 0.000822 |
| Locus 28 CEP120/.. | 1.23e-10 | 2.76e-09 | 0.000455 | 2.63e-09 | 2.72e-08 | 0.000984 | 8.72e-05 | 0.00266 | 8.63e-05 | 0.00375 | 0.00157 | 0.000612 | 0.00159 | 0.00125 | 0.000332 | 0.00193 | 0.00218 | 0.00215 | 7.89e-05 | 0.000548 |
| Locus 32 DOK3 | 1.85e-08 | 5.73e-07 | 0.00176 | 1.48e-08 | 1.89e-07 | 0.00128 | 0.00204 | 0.00183 | 0.0166 | 0.0234 | 0.0444 | 0.0181 | 0.0603 | 0.0171 | 0.0303 | 0.00389 | 0.0162 | 0.000504 | 0.00754 | 0.000344 |
| Locus 37 RUNX2 | 4.06e-08 | 3.21e-07 | 0.00397 | 9.5e-08 | 4.42e-07 | 0.00395 | 0.00262 | 0.00565 | 0.0104 | 0.0106 | 0.00724 | 0.028 | 0.0015 | 0.00281 | 0.00956 | 0.0013 | 0.0488 | 0.00023 | 0.003 | 0.000815 |
| Locus 39 FAM135A/.. | 1.59e-08 | 1.5e-07 | 6.28e-05 | 3.23e-09 | 1.08e-07 | 4.16e-05 | 0.000347 | 0.00456 | 0.00219 | 0.00894 | 0.00449 | 0.00215 | 0.009 | 0.0223 | 0.000253 | 0.00605 | 0.00946 | 0.000278 | 0.00335 | 0.000171 |
| Locus 40 RGS17 | 1.15e-08 | 9.86e-10 | 0.00534 | 6.21e-08 | 1.69e-08 | 0.00531 | 0.00103 | 0.000477 | 0.0028 | 0.00179 | 0.00287 | 0.000626 | 0.00141 | 0.000356 | 0.00607 | 0.00955 | 0.0138 | 0.013 | 0.003 | 0.000135 |
| Locus 41 MAFK | 3.08e-08 | 4.48e-07 | 0.00181 | 3.06e-08 | 2.48e-06 | 0.00113 | 0.00106 | 0.00202 | 0.000565 | 0.00222 | 0.00399 | 0.0115 | 0.0806 | 0.0263 | 0.0119 | 0.00259 | 0.0015 | 0.000422 | 0.00428 | 0.000297 |
| Locus 42 PMS2/.. | 4.21e-09 | 1.16e-08 | 0.00334 | 1e-09 | 2.29e-09 | 0.00372 | 0.00121 | 0.00472 | 2.67e-05 | 0.0136 | 0.00182 | 0.00648 | 0.0182 | 0.069 | 0.000997 | 0.00421 | 0.0126 | 0.00158 | 0.00145 | 6.63e-06 |
| Locus 45 HDAC9 | 8.39e-11 | 3.13e-11 | 0.000514 | 9.29e-11 | 7.46e-11 | 0.00222 | 0.000137 | 0.00655 | 1.43e-07 | 0.00336 | 0.000922 | 0.0019 | 0.00184 | 0.0019 | 0.00171 | 0.00182 | 0.00695 | 0.000277 | 0.000178 | 8.74e-06 |
| Locus 50 DOCK4 | 3.48e-08 | 2.09e-06 | 0.00255 | 2.69e-08 | 1.72e-06 | 0.00177 | 0.000786 | 0.00106 | 8.27e-06 | 0.052 | 0.027 | 0.00623 | 0.013 | 0.0167 | 0.0061 | 0.000176 | 0.00502 | 3e-05 | 0.00124 | 0.00164 |
| Locus 55 RRM2B/.. | 2.48e-08 | 1.05e-07 | 0.000235 | 9.48e-09 | 3.4e-08 | 0.000376 | 0.00542 | 0.00379 | 0.00178 | 0.000485 | 0.000403 | 0.0149 | 0.00354 | 0.00467 | 0.00455 | 2.78e-06 | 0.00105 | 1.84e-05 | 0.000255 | 0.0019 |
| Locus 57 SLA | 5.23e-09 | 3.84e-10 | 0.000465 | 3.09e-09 | 2.81e-10 | 0.000462 | 0.000165 | 0.00108 | 0.000686 | 0.00357 | 0.00391 | 0.00268 | 0.0325 | 0.0011 | 0.000495 | 0.00129 | 0.00102 | 0.0024 | 0.000436 | 0.000474 |
| Locus 59 IFNE/.. | 1.18e-08 | 2.01e-09 | 0.00541 | 2.47e-08 | 3.58e-09 | 0.00471 | 0.00259 | 0.00135 | 0.00286 | 0.0124 | 0.000976 | 0.00995 | 0.00267 | 0.00534 | 0.0037 | 0.000466 | 0.000981 | 0.000631 | 0.000218 | 0.000542 |
| Locus 60 GNAQ | 2.31e-08 | 4.21e-07 | 0.000162 | 5.64e-08 | 1.47e-06 | 0.000243 | 0.00134 | 0.00642 | 0.00142 | 0.00964 | 0.00623 | 0.00569 | 0.00261 | 0.0068 | 0.00351 | 0.0183 | 0.0106 | 0.00675 | 0.000191 | 0.00867 |
| Locus 61 SYK | 2.41e-09 | 1.33e-06 | 1.08e-05 | 1.12e-08 | 5.05e-06 | 1.03e-05 | 3.37e-05 | 2.87e-05 | 0.00242 | 0.00306 | 0.00268 | 0.00321 | 0.0216 | 0.00432 | 0.000427 | 2.56e-05 | 0.00138 | 1.8e-05 | 0.000245 | 0.00137 |
| Locus 62 FGD3/.. | 4.55e-08 | 7.14e-08 | 0.00405 | 5.1e-07 | 5.01e-07 | 0.00404 | 0.000128 | 4.16e-05 | 0.00298 | 0.0352 | 0.0312 | 0.00459 | 0.0186 | 0.0198 | 0.00662 | 0.00508 | 0.014 | 0.00734 | 0.00127 | 0.00157 |
| Locus 68 GPAM | 5.47e-08 | 1.67e-07 | 0.000691 | 4.54e-08 | 1.69e-07 | 0.00047 | 0.000567 | 0.00125 | 0.00146 | 0.0542 | 0.00485 | 0.00296 | 0.00844 | 0.0188 | 0.000638 | 0.00322 | 0.0187 | 0.0224 | 0.00778 | 0.00415 |
| Locus 72 CFL1/.. | 1.6e-10 | 4.93e-08 | 7.02e-05 | 1.44e-10 | 6.43e-08 | 9.48e-05 | 0.000125 | 6.4e-05 | 0.000429 | 0.00465 | 0.00827 | 0.00214 | 0.0231 | 0.00653 | 0.000466 | 0.00408 | 0.00497 | 0.00329 | 2.53e-06 | 0.00242 |
| Locus 75 NINJ2 | 2.12e-08 | 1.22e-07 | 0.000301 | 8.85e-08 | 8.52e-07 | 0.000442 | 0.000505 | 0.000446 | 0.00551 | 0.00422 | 0.00348 | 0.002 | 0.0234 | 0.0123 | 0.0211 | 0.00329 | 0.0117 | 0.00403 | 0.00085 | 0.000111 |
| Locus 76 GLS2/.. | 2.44e-10 | 7.02e-10 | 0.000231 | 1.19e-09 | 9.74e-10 | 0.000231 | 0.00266 | 0.00113 | 5.05e-05 | 1.81e-05 | 1.33e-05 | 0.00073 | 0.0378 | 0.0108 | 0.00282 | 0.00202 | 0.00738 | 0.00362 | 0.00582 | 0.000854 |
| Locus 78 HIP1R/.. | 1.76e-08 | 2.54e-06 | 0.000106 | 1.83e-07 | 3.21e-07 | 0.000468 | 0.000225 | 0.00194 | 0.000741 | 0.00719 | 0.0296 | 0.00167 | 0.00258 | 0.00335 | 0.00906 | 0.0147 | 0.00575 | 0.0061 | 0.00188 | 0.000249 |
| Locus 79 ATP6V0A2/.. | 2.39e-09 | 1.73e-08 | 6.51e-05 | 1.4e-08 | 2.62e-08 | 8.65e-06 | 0.000214 | 0.000422 | 0.000545 | 0.00374 | 0.0108 | 0.0208 | 0.0101 | 0.0414 | 0.000446 | 0.00255 | 0.0107 | 0.00431 | 0.00123 | 0.000737 |
| Locus 80 MCF2L | 1.29e-07 | 9.56e-08 | 0.00106 | 4.89e-08 | 6.37e-08 | 0.000796 | 0.00138 | 0.000128 | 0.000647 | 0.0017 | 0.00203 | 0.00176 | 0.0058 | 0.0387 | 0.00084 | 0.00133 | 0.0102 | 0.00173 | 0.00121 | 2.97e-05 |
| Locus 85 RASGRP1 | 1.22e-09 | 3.82e-10 | 0.000855 | 2.28e-08 | 6.99e-09 | 0.000508 | 0.00188 | 0.000384 | 0.00131 | 0.00422 | 0.016 | 0.0137 | 0.0279 | 0.00848 | 0.00292 | 0.0199 | 0.00116 | 0.00271 | 8.22e-05 | 2.17e-06 |
| Locus 86 PLCB2 | 4.48e-08 | 4.78e-09 | 0.00059 | 3.78e-09 | 1.29e-09 | 0.000611 | 0.000405 | 0.00274 | 0.00266 | 0.00036 | 0.00464 | 0.00263 | 0.0094 | 0.00518 | 0.0174 | 0.00603 | 0.00649 | 0.00915 | 0.00562 | 0.000127 |
| Locus 90 GLCE | 1.4e-07 | 2.19e-07 | 0.00309 | 3.56e-08 | 1.76e-07 | 0.0043 | 0.00251 | 0.00151 | 0.0013 | 0.00161 | 0.00153 | 0.0052 | 0.000147 | 0.0103 | 0.000218 | 0.013 | 0.0107 | 0.00284 | 0.000531 | 0.00374 |
| Locus 92 MCTP2 | 1.25e-07 | 1.29e-07 | 0.00165 | 3.92e-08 | 1.2e-07 | 0.00164 | 0.000456 | 0.000544 | 0.00053 | 0.0168 | 0.0173 | 0.00162 | 0.00504 | 0.0045 | 0.00135 | 0.0011 | 0.00596 | 0.00307 | 0.00302 | 0.000881 |
| Locus 94 GGA2 | 1.06e-09 | 2.2e-08 | 0.00017 | 5.2e-08 | 1.64e-07 | 0.00019 | 0.000393 | 0.00082 | 0.000936 | 0.00998 | 0.0128 | 0.00311 | 0.0116 | 0.00812 | 0.00128 | 0.00302 | 0.00489 | 0.000496 | 0.001 | 0.00014 |
| Locus 98 PMFBP1 | 1.15e-08 | 1.16e-07 | 0.000465 | 1.82e-09 | 2.09e-08 | 0.000162 | 0.000374 | 0.00422 | 0.00283 | 0.00593 | 0.0114 | 0.0506 | 7.68e-05 | 0.000434 | 0.000839 | 0.000333 | 0.0203 | 0.00018 | 0.000591 | 0.00159 |
| Locus 101 C16orf95 | 7.02e-12 | 5.44e-11 | 0.000454 | 9.86e-10 | 7.66e-10 | 0.000912 | 0.000261 | 0.00018 | 0.000981 | 0.0052 | 0.0188 | 0.0014 | 0.00606 | 0.0227 | 0.000997 | 0.00147 | 0.00189 | 0.000301 | 0.000942 | 0.000208 |
| Locus 104 MNT/.. | 6.16e-08 | 1.34e-06 | 0.00148 | 2.92e-08 | 8.39e-07 | 0.00252 | 0.00368 | 0.00245 | 0.00197 | 0.0289 | 0.0795 | 0.00702 | 0.00829 | 0.0019 | 0.00154 | 0.000944 | 0.0144 | 0.00128 | 0.000793 | 0.00309 |
| Locus 113 AC104532.2/.. | 5.7e-08 | 9.84e-06 | 0.000224 | 2.95e-08 | 9.19e-06 | 4.59e-05 | 0.00126 | 0.00211 | 0.000117 | 0.000825 | 0.000521 | 0.00281 | 0.0485 | 0.00929 | 0.0037 | 0.00556 | 0.0123 | 0.00191 | 0.000311 | 0.000436 |
| Locus 114 DNM2/.. | 1.55e-08 | 1.56e-06 | 0.000333 | 1.69e-07 | 9.86e-06 | 0.000782 | 0.00282 | 0.000572 | 0.00124 | 0.0283 | 0.0225 | 0.056 | 0.00551 | 0.0149 | 0.0142 | 0.000128 | 0.0122 | 3.63e-05 | 0.00016 | 0.00127 |
| Locus 115 LRRC25/.. | 1.11e-10 | 8.93e-12 | 0.00357 | 9.29e-12 | 1.54e-12 | 0.00373 | 0.00134 | 0.00133 | 0.00409 | 0.00127 | 8e-04 | 0.0188 | 0.000386 | 0.00553 | 6.29e-05 | 0.00116 | 0.00283 | 0.00116 | 0.000334 | 0.00114 |
| Locus 116 YJEFN3/.. | 6.9e-11 | 1.22e-09 | 0.000787 | 8.89e-10 | 4.11e-09 | 0.00163 | 0.00371 | 0.0064 | 0.000759 | 0.00597 | 0.0039 | 0.00671 | 0.00229 | 0.00898 | 0.000179 | 0.0141 | 0.0108 | 0.00171 | 0.00279 | 9.35e-06 |
| Locus 117 CEP89/.. | 7.41e-18 | 1.04e-15 | 4.17e-05 | 4.51e-18 | 7.23e-16 | 0.000145 | NA | NA | NA | 0.00532 | 0.00708 | 0.000412 | 0.00487 | 0.00167 | 0.0175 | 0.000289 | 0.000837 | 0.00236 | 5.46e-05 | 3.28e-06 |
| Locus 118 AXL | 7.47e-09 | 2.11e-07 | 0.000648 | 4.05e-09 | 1.45e-07 | 0.000628 | 0.000999 | 0.000136 | 0.00228 | 0.0225 | 0.0232 | 0.00284 | 0.0112 | 0.000743 | 0.0043 | 0.00236 | 0.00901 | 0.00675 | 0.000437 | 0.00432 |
| Locus 124 SMOX | 7.69e-09 | 4.26e-08 | 0.00145 | 3.88e-09 | 2.75e-08 | 0.0017 | 0.00396 | 0.00989 | 0.000424 | 0.00496 | 0.007 | 0.00112 | 0.0193 | 0.00512 | 0.000154 | 0.0025 | 0.00527 | 0.00483 | 0.00104 | 0.000221 |
| Locus 125 UBE2V1/.. | 7.47e-09 | 1.53e-07 | 0.0047 | 2.12e-08 | 2.02e-07 | 0.00132 | 0.0022 | 0.00102 | 0.00234 | 2.81e-05 | 2.78e-05 | 0.00458 | 0.0259 | 0.00508 | 0.00405 | 0.00237 | 0.00024 | 0.000671 | 0.000789 | 0.000307 |

**Supplementary Figure 6. The P-values of the new loci in the combined and stratified meta-analyses.** Red cells represent loci containing a significant variant ($P<5x10^{-8}$), orange cells represent loci containing a suggestive variant ($P<1x10^{-5}$). Male represents the no proxy all ancestry male only analysis; Female represents the no proxy all ancestry female only analysis. The max effective sample sizes are listed in brackets.

| | All_Combined (414,019) | All_noProxy (353,193) | All_Proxy (60,826) | EUR_Combined (378,916) | EUR_noProxy (329,373) | EUR_Proxy (49,543) | AFR_Combined(19,354) | AFR_noProxy (14,555) | AFR_Proxy (4799) | AMR_Combined (1217) | AMR_noProxy(863) | AMR_Proxy (354) | EAS_Combined(13,681) | EAS_noProxy(8034) | EAS_Proxy(5647) | SAS_Combined(851) | SAS_noProxy (368) | SAS_Proxy(483) | Male (72,196) | Female (105,300) |
|---|---|---|---|---|---|---|---|---|---|---|---|---|---|---|---|---|---|---|---|---|
| Locus 4 GBA1 | 1.83e-09 | 8.43e-09 | 0.00575 | 1.94e-09 | 7.44e-09 | 0.00434 | 4.13e-05 | 6.53e-06 | 0.00313 | 0.00599 | 0.00691 | 0.00308 | 0.0295 | 0.0274 | 0.00163 | 0.00547 | 0.0184 | 0.0011 | 0.000333 | 0.00627 |
| Locus 5 ADAMTS4 | 3.07e-08 | 2.4e-06 | 3.48e-06 | 1.51e-07 | 6.77e-06 | 1.14e-06 | 7.85e-05 | 0.00128 | 0.000504 | 0.0067 | 0.0172 | 0.00304 | 0.00556 | 0.00384 | 0.00514 | 0.000909 | 0.00252 | 0.00256 | 0.00226 | 0.00177 |
| Locus 7 CR1 | 1.21e-57 | 7.36e-50 | 2.73e-10 | 1.05e-57 | 2.05e-49 | 1.59e-10 | 0.000157 | 9.03e-05 | 9.62e-06 | 0.00216 | 0.00469 | 0.000445 | 0.00431 | 0.00718 | 0.00213 | 0.00231 | 0.00197 | 0.000488 | 2.76e-09 | 6.05e-17 |
| Locus 8 QPCT | 3.98e-16 | 2.26e-14 | 8.95e-05 | 3.64e-17 | 2.5e-15 | 0.000134 | 0.000142 | 0.00046 | 0.00151 | 0.00282 | 0.00176 | 0.000846 | 0.00754 | 0.000993 | 0.000243 | 0.00363 | 0.00145 | 0.00135 | 0.000175 | 1.04e-06 |
| Locus 9 SPRED2 | 2.75e-09 | 2.16e-07 | 0.00103 | 1.2e-10 | 2.09e-08 | 0.00011 | 0.00148 | 0.000389 | 0.000696 | 0.00128 | 0.00018 | 0.0128 | 0.054 | 0.0173 | 0.00595 | 0.00246 | 0.0111 | 0.00263 | 0.00057 | 0.00309 |
| Locus 10 NCK2 | 8.19e-17 | 1.17e-12 | 9.43e-08 | 2.39e-16 | 3.8e-13 | 5.11e-06 | 0.000469 | 0.00222 | 0.00566 | 0.000675 | 0.000458 | 4.45e-05 | 0.00737 | 0.0133 | 1.2e-05 | 0.000781 | 0.00217 | 0.000585 | 0.00272 | 0.000166 |
| Locus 11 BIN1 | 1.85e-108 | 6.4e-90 | 2.16e-21 | 3.09e-110 | 4.14e-91 | 7.39e-22 | 2.2e-05 | 2.74e-05 | 0.00013 | 0.00132 | 0.00165 | 0.00318 | 0.00227 | 0.00871 | 0.00213 | 0.00274 | 0.000254 | 0.00175 | 2.02e-10 | 1.95e-24 |
| Locus 12 TMEM163 | 5.68e-07 | 1.45e-05 | 0.000422 | 1.2e-08 | 1.15e-06 | 0.000266 | 0.0016 | 0.000273 | 0.00253 | 0.00432 | 0.0051 | 0.059 | 0.0184 | 0.00211 | 0.00618 | 0.0103 | 0.0132 | 0.00246 | 0.00489 | 0.00325 |
| Locus 13 WDR12/.. | 7.67e-13 | 7.07e-11 | 0.000518 | 4.34e-13 | 8.32e-11 | 0.000214 | 0.00134 | 0.00126 | 0.00196 | 0.00314 | 0.00218 | 0.012 | 0.059 | 0.0265 | 0.0201 | 0.000603 | 0.00202 | 0.000378 | 0.000136 | 0.00154 |
| Locus 14 INPP5D | 5.1e-19 | 4.21e-14 | 1.45e-07 | 9.6e-19 | 6.71e-14 | 4.65e-07 | 0.000603 | 0.00392 | 0.000785 | 0.00065 | 0.000689 | 0.0104 | 0.00408 | 0.00187 | 0.000146 | 0.00235 | 0.00877 | 0.000788 | 0.00379 | 2.19e-05 |
| Locus 18 MME | 1.41e-10 | 1.58e-08 | 0.000352 | 3.35e-10 | 3.55e-08 | 0.000286 | 3.16e-05 | 0.000981 | 5.09e-06 | 0.00374 | 0.00445 | 0.00689 | 0.00879 | 0.00487 | 0.00104 | 0.00224 | 0.0107 | 0.00248 | 8.62e-05 | 5e-04 |
| Locus 19 VWA5B2 | 2.1e-08 | 8.7e-09 | 0.00101 | 4.68e-07 | 1.77e-07 | 0.0064 | 1.83e-06 | 9.12e-06 | 0.00252 | 0.00974 | 0.00205 | 0.00484 | 0.00321 | 0.00261 | 0.000291 | 0.00393 | 0.00717 | 0.00424 | 6.91e-06 | 0.00123 |
| Locus 22 FGFRL1/.. | 2.09e-09 | 4.56e-09 | 0.000218 | 1.09e-09 | 2.98e-09 | 0.000214 | 0.000495 | 0.000266 | 0.000571 | 0.00202 | 0.000839 | 0.00487 | 0.014 | 0.00456 | 0.000199 | 0.00102 | 0.00596 | 0.00393 | 0.000222 | 0.000118 |
| Locus 23 CLNK | 1.2e-18 | 7.38e-14 | 8.77e-08 | 8e-19 | 1.88e-14 | 2.47e-07 | 0.000337 | 0.000699 | 3.74e-05 | 0.0182 | 0.00491 | 0.00416 | 0.000537 | 0.0081 | 0.000324 | 0.00121 | 0.00746 | 0.00142 | 0.00119 | 0.00139 |
| Locus 24 RHOH | 4.45e-09 | 1.23e-08 | 0.000349 | 1.26e-09 | 2.3e-09 | 0.000541 | 8.75e-05 | 0.000133 | 0.00142 | 0.00274 | 0.0084 | 0.00725 | 0.00289 | 0.00172 | 0.003 | 0.00114 | 0.00169 | 0.00067 | 2.97e-05 | 3.09e-05 |
| Locus 26 FAM105B | 1.22e-09 | 4.69e-09 | 0.000559 | 1.05e-09 | 1.31e-09 | 0.000772 | 0.000542 | 0.000326 | 7.31e-05 | 0.00765 | 0.00718 | 0.0093 | 0.00174 | 0.00404 | 0.00026 | 0.00292 | 0.0128 | 0.00336 | 0.00183 | 3.48e-05 |
| Locus 27 AC008394.1/.. | 5.51e-12 | 7.29e-11 | 1.6e-05 | 1.68e-12 | 8.75e-11 | 5.5e-05 | 3.63e-05 | 0.000117 | 0.000742 | 0.0144 | 0.0178 | 0.00021 | 0.000223 | 0.000437 | 0.00104 | 0.000132 | 0.00203 | 0.000408 | 7.22e-05 | 4.37e-06 |
| Locus 29 HBEGF | 2.3e-09 | 4.69e-08 | 0.00109 | 8.49e-10 | 4.26e-08 | 0.000334 | 0.00134 | 0.00093 | 0.000523 | 0.0172 | 0.0137 | 0.0227 | 1.67e-05 | 0.0121 | 1.52e-05 | 0.0245 | 0.00237 | 0.0461 | 1.03e-05 | 0.000481 |
| Locus 30 TNIP1 | 5.86e-18 | 1.95e-14 | 8.99e-05 | 1.61e-18 | 1.39e-15 | 0.000233 | 0.000623 | 0.00111 | 4e-04 | 0.0109 | 0.00956 | 0.000446 | 0.000464 | 0.00482 | 0.000113 | 0.00686 | 0.0289 | 0.00901 | 7.32e-05 | 6.69e-06 |
| Locus 31 HAVCR2 | 2.08e-09 | 2.44e-08 | 0.000352 | 8.35e-09 | 3.57e-08 | 0.000391 | 0.00452 | 0.00682 | 0.000995 | 0.0225 | 0.00522 | 0.00107 | 0.0102 | 0.00422 | 0.0081 | 0.00171 | 0.0105 | 0.00116 | 0.00132 | 0.00019 |
| Locus 33 RASGEF1C | 7.29e-14 | 1.68e-11 | 0.000173 | 1.19e-13 | 2.87e-11 | 0.000444 | 0.000144 | 5.67e-05 | 0.00113 | 0.00727 | 0.00382 | 0.0149 | 0.00186 | 0.00789 | 0.00294 | 0.0026 | 0.00257 | 0.00245 | 1.14e-05 | 0.00059 |
| Locus 34 OR2B2/.. | 1.18e-09 | 2.5e-07 | 3.62e-05 | 4.65e-09 | 5.17e-07 | 7.72e-05 | 0.000296 | 0.000959 | 0.000126 | 0.00731 | 0.00496 | 0.00855 | 2.88e-06 | 0.000247 | 0.00125 | 0.00084 | 0.00956 | 0.00037 | 0.000151 | 2.22e-08 |
| Locus 35 HLA | 6.48e-29 | 1.58e-26 | 7.8e-11 | 4.19e-31 | 8.88e-27 | 6.12e-11 | 0.000236 | 0.000166 | 0.00011 | 0.000518 | 0.000896 | 0.000566 | 5.18e-06 | 6.59e-05 | 9.04e-06 | 0.00036 | 0.000648 | 0.000568 | 1.11e-05 | 5.99e-08 |
| Locus 36 TREM2 | 2.23e-24 | 8.64e-18 | 1.15e-08 | 2.53e-24 | 8.06e-18 | 1.13e-08 | 4.38e-09 | 7.95e-07 | 0.000141 | 0.000553 | 0.000457 | 0.00168 | 0.00101 | 0.000351 | 0.000641 | 6.46e-05 | 0.00124 | 0.000111 | 9.71e-06 | 1.49e-08 |
| Locus 38 CD2AP | 1.06e-29 | 6.24e-26 | 2.82e-06 | 1.66e-27 | 2.82e-25 | 2.37e-06 | 2.81e-06 | 7.39e-05 | 9.61e-05 | 0.00494 | 0.00928 | 0.0189 | 0.0332 | 0.00168 | 0.00157 | 0.00293 | 0.00396 | 0.00126 | 4.89e-05 | 6.38e-05 |
| Locus 43 UMAD1/.. | 2.55e-08 | 1.35e-08 | 0.00145 | 2.74e-08 | 2.2e-08 | 0.00214 | 9.36e-05 | 0.000126 | 0.000648 | 0.00207 | 0.000641 | 0.0188 | 0.0083 | 0.00822 | 0.000889 | 0.00128 | 0.00329 | 0.00112 | 0.000439 | 0.00137 |
| Locus 44 TMEM106B | 4.78e-23 | 5.49e-20 | 2.85e-05 | 8.58e-22 | 9.93e-20 | 2.85e-05 | 9.02e-06 | 0.000137 | 0.000286 | 0.0191 | 0.0394 | 0.00112 | 0.000911 | 0.000976 | 0.00161 | 0.000913 | 0.00614 | 0.000249 | 0.001 | 5.82e-07 |
| Locus 46 JAZF1 | 3.81e-11 | 8.04e-11 | 0.000153 | 2.36e-10 | 6.96e-09 | 0.000238 | 0.00095 | 0.000293 | 0.00342 | 0.00072 | 0.000118 | 0.000471 | 0.0013 | 0.0231 | 0.00101 | 0.00013 | 0.00342 | 0.000718 | 0.000195 | 4.86e-05 |
| Locus 47 ELMO1/.. | 2.27e-12 | 4.91e-11 | 0.000766 | 3.89e-12 | 1.21e-11 | 0.00337 | 0.000251 | 0.00113 | 0.000544 | 0.000974 | 0.000191 | 0.00414 | 0.00566 | 0.0168 | 0.000356 | 0.0033 | 0.00135 | 0.000623 | 3.21e-06 | 1.77e-05 |
| Locus 48 EGFR | 4.57e-11 | 1.46e-08 | 4.88e-06 | 6.88e-12 | 1.84e-09 | 8.54e-06 | 0.00116 | 0.001 | 0.000684 | 6.24e-05 | 1.05e-05 | 0.000713 | 0.00505 | 0.00424 | 0.00369 | 0.00082 | 0.00616 | 0.000722 | 0.00192 | 0.000341 |
| Locus 49 ZCWPW1/.. | 9.57e-38 | 3.88e-28 | 4.5e-12 | 5.66e-38 | 5.75e-29 | 7.01e-12 | 0.00438 | 0.000782 | 0.000252 | 0.000525 | 9.94e-05 | 0.00227 | 0.00438 | 0.0235 | 0.00278 | 0.0071 | 0.0101 | 0.00254 | 1.54e-07 | 5.9e-08 |
| Locus 51 EPHA1 | 4.2e-14 | 4.16e-11 | 4.03e-07 | 8.08e-13 | 2.37e-10 | 2.17e-06 | 0.00066 | 0.000894 | 0.00204 | 0.0162 | 0.0156 | 0.00176 | 0.0404 | 0.0124 | 0.00274 | 0.0119 | 0.014 | 0.00168 | 0.00137 | 4.59e-05 |
| Locus 52 CTSB | 1.38e-09 | 2.21e-09 | 0.000276 | 6.34e-10 | 8.87e-10 | 0.00123 | 2.92e-05 | 3.03e-05 | 0.000169 | 0.00487 | 0.0102 | 0.00259 | 0.00645 | 0.0189 | 3.7e-05 | 0.0018 | 0.00133 | 0.00104 | 0.000494 | 9.14e-05 |
| Locus 53 CLU | 7.6e-48 | 5.9e-40 | 1.03e-10 | 3.21e-47 | 7.29e-40 | 7.84e-10 | 9.35e-06 | 1.53e-05 | 0.00177 | 0.00232 | 0.0023 | 0.00167 | 5.96e-06 | 0.000119 | 0.000431 | 0.00122 | 0.00922 | 0.00129 | 1.08e-06 | 4.79e-09 |
| Locus 54 TP53INP1 | 1.66e-10 | 2.07e-08 | 0.00123 | 1.14e-09 | 1.33e-07 | 0.000621 | 0.000247 | 0.00172 | 0.00217 | 0.00717 | 0.00512 | 0.00219 | 0.0117 | 0.0123 | 0.00477 | 0.00899 | 0.0119 | 0.0113 | 0.00162 | 0.000103 |
| Locus 56 TRIB1 | 5.71e-12 | 1.05e-10 | 0.00395 | 1.27e-11 | 1.43e-10 | 0.00425 | 9.96e-05 | 0.000123 | 0.000108 | 0.00369 | 0.00712 | 0.002 | 0.00202 | 0.000342 | 0.000729 | 0.00109 | 0.000722 | 8.67e-05 | 0.000401 | 0.000289 |
| Locus 58 SHARPIN | 3.52e-23 | 2.81e-23 | 0.000131 | 3.14e-23 | 2.9e-23 | 0.000118 | 0.000128 | 6.79e-05 | 0.000449 | 0.0123 | 0.0128 | 0.0277 | 0.0191 | 0.018 | 0.00332 | 0.0116 | 0.0175 | 0.00242 | 8.16e-05 | 7.24e-07 |
| Locus 63 ABCA1 | 1.16e-09 | 1.85e-09 | 0.000163 | 8.5e-09 | 1.12e-08 | 0.00176 | 0.00015 | 0.000358 | 1e-04 | 0.00203 | 0.00435 | 0.00286 | 0.00382 | 0.00091 | 0.00129 | 0.00133 | 0.00558 | 0.00157 | 0.000128 | 0.000346 |
| Locus 64 USP6NL | 8.93e-28 | 4.11e-26 | 3.04e-05 | 1.09e-28 | 3.1e-26 | 1.3e-05 | 5.13e-05 | 0.000522 | 0.000304 | 0.00181 | 0.00276 | 0.0025 | 0.0012 | 0.00151 | 0.00244 | 0.00282 | 0.00494 | 0.000405 | 1.58e-05 | 9.5e-08 |
| Locus 65 CCDC6/.. | 5.5e-16 | 1.33e-13 | 1.46e-06 | 1.35e-16 | 2.43e-14 | 3.97e-07 | 0.00315 | 0.00368 | 3.98e-05 | 0.00381 | 0.00337 | 0.000917 | 0.00232 | 0.00352 | 0.00167 | 0.00291 | 0.000342 | 0.0108 | 0.00111 | 4.04e-05 |
| Locus 66 TSPAN14 | 1.12e-13 | 2.35e-12 | 0.000369 | 1.38e-13 | 1.83e-12 | 0.000328 | 0.000852 | 0.00172 | 0.000726 | 0.012 | 0.0176 | 0.000963 | 0.0133 | 0.00482 | 0.00172 | 0.00254 | 0.00509 | 0.000382 | 6.75e-05 | 0.00025 |
| Locus 67 BLNK | 1.54e-15 | 3.15e-12 | 2.22e-05 | 1.73e-15 | 3.17e-12 | 4.38e-05 | 0.000866 | 0.00106 | 0.00294 | 0.0168 | 0.00898 | 0.00489 | 0.0186 | 0.00116 | 0.0014 | 0.00194 | 0.0124 | 0.000179 | 7.5e-05 | 0.000199 |
| Locus 69 PLEKHA1 | 1.01e-12 | 6.08e-11 | 0.000311 | 8.05e-11 | 6.89e-10 | 0.000274 | 8.11e-06 | 0.000336 | 7.83e-05 | 0.00531 | 0.00316 | 9.94e-06 | 0.0108 | 0.0078 | 0.00519 | 0.00529 | 0.00332 | 0.00323 | 2.31e-05 | 0.00166 |
| Locus 70 SPI1 | 7.85e-22 | 3.22e-18 | 1.7e-05 | 4.88e-21 | 1.17e-17 | 4.58e-05 | 1.33e-05 | 0.000392 | 0.000283 | 0.0124 | 0.0024 | 0.00029 | 0.00399 | 0.00142 | 0.000131 | 0.00666 | 0.00387 | 0.00208 | 8.01e-07 | 8.15e-05 |
| Locus 71 MS4A gene cluster | 2.7e-73 | 1.09e-69 | 4.19e-08 | 8.37e-75 | 2.41e-68 | 1.04e-08 | 2.95e-05 | 7.44e-05 | 0.000959 | 0.00011 | 0.000104 | 0.00863 | 0.00184 | 0.0118 | 0.000463 | 0.00168 | 0.0132 | 0.00134 | 2.28e-10 | 5.63e-12 |
| Locus 73 PICALM | 7.9e-66 | 9.42e-56 | 7.81e-13 | 1.21e-63 | 9.6e-53 | 1.62e-13 | 8.02e-06 | 0.000204 | 0.000375 | 0.00661 | 0.00165 | 0.00206 | 0.000775 | 8.11e-06 | 6.69e-05 | 0.00214 | 0.00366 | 0.000408 | 6.9e-09 | 5.31e-14 |
| Locus 74 SORL1 | 1.2e-26 | 9.21e-23 | 1.27e-05 | 1.22e-21 | 7.3e-17 | 2.85e-06 | 3.46e-09 | 9.95e-07 | 0.000112 | 0.00892 | 0.0136 | 0.0103 | 4.9e-09 | 8.03e-11 | 0.000578 | 0.00539 | 0.00569 | 0.0188 | 0.00143 | 0.000252 |
| Locus 77 TPCN1 | 1.23e-12 | 6.46e-11 | 0.000954 | 2.6e-12 | 6.9e-11 | 0.00164 | 0.000906 | 0.00155 | 0.000113 | 0.013 | 0.0123 | 0.000399 | 0.00309 | 0.00443 | 0.000831 | 0.00551 | 0.00795 | 0.00103 | 7.81e-05 | 0.000837 |
| Locus 81 FERMT2 | 6.79e-22 | 5.43e-19 | 4.43e-05 | 2.43e-21 | 2.71e-19 | 8.15e-05 | 0.000248 | 0.00065 | 0.000354 | 0.00645 | 0.000908 | 0.00229 | 0.00192 | 0.00965 | 0.00354 | 0.0139 | 0.00263 | 0.00153 | 5.29e-05 | 0.000538 |
| Locus 82 SLC24A4 | 3.03e-25 | 1.16e-20 | 1.02e-08 | 1.38e-26 | 9.2e-21 | 4.97e-08 | 0.000902 | 0.00348 | 5.81e-05 | 0.00643 | 0.000704 | 0.00152 | 0.00055 | 0.00183 | 1.09e-06 | 0.000147 | 0.0161 | 0.00018 | 7.57e-05 | 1e-06 |
| Locus 83 IGH gene cluster | 1.47e-11 | 1.18e-08 | 0.000118 | 2.4e-11 | 1.59e-08 | 0.000143 | 0.000303 | 0.00492 | 0.000448 | 0.0446 | 0.0507 | 0.126 | 0.0786 | 0.0902 | 0.00723 | 0.0178 | 0.0335 | 0.00277 | 0.0324 | 0.0418 |
| Locus 84 KIAA0125/.. | 5.88e-08 | 2.53e-07 | 0.00443 | 6.28e-09 | 1.03e-07 | 0.00407 | 0.000688 | 7.64e-05 | 0.000549 | 0.00571 | 0.0331 | 0.0139 | 0.000494 | 0.0104 | 0.0106 | 0.0027 | 0.0219 | 0.00586 | 0.0172 | 0.00594 |
| Locus 87 ADAM10 | 3.87e-23 | 1.88e-18 | 7.49e-08 | 2.9e-23 | 1.49e-18 | 1.31e-07 | 0.00167 | 0.00252 | 0.000878 | 0.0013 | 0.0174 | 0.00139 | 0.014 | 0.000951 | 0.00265 | 0.00121 | 0.0119 | 0.000229 | 0.000479 | 4.15e-05 |
| Locus 88 APH1B | 1.45e-25 | 2.23e-20 | 4.08e-07 | 5.04e-26 | 7.47e-21 | 5.73e-07 | 0.00148 | 0.000999 | 0.000185 | 0.00313 | 0.00606 | 0.00303 | 0.0122 | 0.00502 | 0.00274 | 0.005 | 0.00315 | 0.00618 | 0.00312 | 1.36e-10 |
| Locus 89 SNX1 | 4.47e-16 | 3.13e-16 | 0.000609 | 6.73e-17 | 8.27e-17 | 0.00074 | 1.68e-05 | 0.000108 | 0.000611 | 0.00106 | 0.00112 | 0.00263 | 0.00182 | 0.00391 | 0.00226 | 0.0023 | 0.0106 | 0.00071 | 0.00015 | 1.21e-07 |
| Locus 91 CTSH | 2.11e-09 | 8.81e-08 | 0.000455 | 5.01e-09 | 1.86e-07 | 0.00395 | 0.00262 | 0.00117 | 3.8e-05 | 0.0178 | 0.00892 | 0.00984 | 0.00229 | 0.0263 | 0.00031 | 0.000424 | 0.00581 | 0.000925 | 0.000707 | 0.00176 |
| Locus 93 IQCK | 1.82e-08 | 1.59e-08 | 0.00036 | 2.78e-08 | 5.76e-09 | 0.00238 | 0.00185 | 0.00362 | 0.00289 | 4.49e-06 | 7.92e-06 | 0.00498 | 0.0237 | 0.004 | 0.00836 | 0.000384 | 0.00616 | 0.00043 | 0.0022 | 0.0024 |
| Locus 95 DOC2A | 2.76e-11 | 3.5e-09 | 0.000197 | 3.52e-11 | 1.35e-09 | 0.000249 | 0.00139 | 0.00244 | 0.00124 | 0.0388 | 0.0282 | 0.0591 | 0.0142 | 0.0129 | 0.0139 | 0.00997 | 0.00701 | 0.0278 | 0.00146 | 0.00117 |
| Locus 96 KAT8/.. | 3.47e-12 | 6.4e-08 | 1.02e-08 | 2.79e-11 | 2.45e-08 | 5.13e-08 | 0.00151 | 0.00336 | 0.000949 | 0.0336 | 0.0107 | 0.0057 | 0.00154 | 0.0177 | 0.00165 | 0.00125 | 0.0266 | 0.000932 | 0.00116 | 0.000105 |
| Locus 97 IL34 | 1.62e-08 | 2.65e-07 | 1.42e-07 | 5.75e-08 | 1.67e-07 | 1.19e-07 | 0.000334 | 0.00312 | 0.00317 | 0.00069 | 0.00186 | 0.000223 | 0.0111 | 0.000684 | 0.00194 | 0.000747 | 0.0429 | 0.00205 | 0.000635 | 0.000155 |
| Locus 99 MAF | 3.15e-09 | 3.58e-08 | 0.00197 | 5.79e-10 | 2.85e-08 | 0.00241 | 0.00254 | 0.000763 | 0.000688 | 0.005 | 0.00555 | 0.00352 | 0.0115 | 0.00348 | 0.00221 | 0.000211 | 0.00401 | 0.00118 | 0.00236 | 0.000416 |
| Locus 100 PLCG2 | 9.51e-15 | 3.46e-13 | 0.000548 | 1.02e-14 | 3.33e-13 | 0.000937 | 0.00113 | 0.000743 | 0.000953 | 0.00841 | 0.00761 | 0.00657 | 0.00209 | 0.00452 | 0.00018 | 0.00443 | 0.00194 | 0.0013 | 3.03e-05 | 5.04e-05 |
| Locus 102 PRDM7/.. | 2e-14 | 8.2e-12 | 0.000115 | 5.32e-14 | 1.13e-11 | 0.000259 | 0.000198 | 9.8e-05 | 0.000751 | 0.00501 | 0.014 | 0.000162 | 0.00838 | 0.0248 | 0.0042 | 0.000186 | 0.000249 | 0.00125 | 0.000305 | 8.88e-05 |
| Locus 103 WDR81 | 2.14e-09 | 2.88e-09 | 8.32e-05 | 2.4e-09 | 6.61e-09 | 0.000869 | 3.05e-05 | 0.00101 | 0.000164 | 0.0207 | 0.0106 | 0.00407 | 0.0134 | 0.00622 | 0.000297 | 0.000756 | 0.0122 | 3.49e-05 | 0.0014 | 0.00148 |
| Locus 105 MINK1/.. | 1.01e-08 | 1.39e-05 | 2.01e-10 | 7.28e-10 | 1.84e-06 | 3.88e-11 | 0.00209 | 0.00285 | 0.00108 | 0.00037 | 0.00166 | 0.00205 | 0.0142 | 0.0116 | 0.000989 | 0.0099 | 0.00474 | 0.00894 | 0.000434 | 0.00225 |
| Locus 106 MYO15A | 1.42e-08 | 3.77e-07 | 0.000589 | 6.27e-09 | 2.49e-07 | 0.00023 | 0.00187 | 0.00202 | 0.000997 | 0.000557 | 0.000912 | 0.00757 | 0.0371 | 0.0289 | 0.00372 | 0.000371 | 0.0135 | 0.00112 | 0.00105 | 2.72e-05 |
| Locus 107 GRN/.. | 7.16e-27 | 3.01e-24 | 0.000314 | 2.18e-26 | 3.03e-24 | 0.000462 | 0.000319 | 0.000962 | 0.00454 | 0.000196 | 0.00015 | 0.00558 | 0.00308 | 0.005 | 0.000982 | 0.00539 | 0.0149 | 0.000947 | 4.75e-06 | 0.0019 |
| Locus 108 MAPT | 4.82e-25 | 8.37e-24 | 0.000154 | 2.86e-25 | 2.37e-24 | 0.000143 | 0.000181 | 0.000174 | 4.39e-05 | 0.00268 | 0.000128 | 0.000217 | 0.000181 | 3.35e-05 | 3.62e-05 | 2.14e-05 | 0.000126 | 0.00163 | 4.05e-07 | 4.07e-08 |
| Locus 109 ABI3 | 1.13e-10 | 1.37e-07 | 7.31e-08 | 2.08e-17 | 4.58e-08 | 7.2e-08 | 0.000128 | 0.000975 | 0.000319 | 0.00161 | 0.00549 | 0.00333 | 0.00146 | 0.000725 | 0.00298 | 0.00334 | 0.00713 | 0.000673 | 2.29e-06 | 0.00317 |
| Locus 110 BZRAP1-AS1/.. | 1.64e-14 | 2.66e-14 | 0.00025 | 1.94e-13 | 1.08e-12 | 0.000365 | 0.00161 | 0.0027 | 0.00146 | 0.00287 | 0.000693 | 0.000571 | 6.78e-05 | 0.00206 | 0.000745 | 0.00231 | 0.00205 | 0.000107 | 4.31e-06 | 1.88e-06 |
| Locus 111 ACE | 9.43e-20 | 6.2e-18 | 0.00196 | 2.11e-19 | 5.9e-18 | 0.00185 | 0.00146 | 0.0031 | 0.00206 | 0.00264 | 0.00635 | 0.0025 | 0.055 | 0.057 | 0.00653 | 0.000114 | 0.033 | 0.000101 | 2.6e-05 | 0.00141 |
| Locus 112 ABCA7 | 5.9e-32 | 2.63e-28 | 7.62e-08 | 7.55e-32 | 3e-28 | 1.32e-07 | 1.85e-06 | 0.000158 | 6.18e-05 | 0.000103 | 1.57e-05 | 0.00253 | 0.00449 | 0.00101 | 0.00976 | 0.00404 | 0.00818 | 0.00514 | 2.55e-06 | 1.93e-10 |
| Locus 119 APOE | 0 | 0 | 0 | 0 | 0 | 0 | 4.58e-108 | 1.14e-62 | 1.66e-52 | 5.51e-05 | 0.000135 | 0.000769 | 2.73e-179 | 1.15e-134 | 2.58e-49 | 6.58e-07 | 7e-07 | 7.8e-06 | 0 | 0 |
| Locus 120 NTN5 | 7.45e-10 | 2.66e-09 | 0.00162 | 3.28e-10 | 1.97e-09 | 0.001 | 0.00158 | 0.001 | 0.0022 | 0.000854 | 0.000361 | 0.0154 | 0.0118 | 0.000655 | 0.00459 | 0.00173 | 0.0265 | 0.000528 | 3.23e-09 | 1.14e-05 |
| Locus 121 CD33 | 1.63e-20 | 1.58e-16 | 4.71e-06 | 3.34e-21 | 3.02e-17 | 3.55e-06 | 7.83e-05 | 0.000554 | 0.000241 | 0.000384 | 0.0015 | 0.00226 | 0.00182 | 0.000479 | 0.0116 | 0.000264 | 0.00127 | 0.000461 | 0.00103 | 3.73e-05 |
| Locus 122 LILRB2 | 2.02e-21 | 1.36e-16 | 1.74e-06 | 8.13e-22 | 2.62e-17 | 2.19e-06 | 0.0015 | 0.00193 | 6.84e-05 | 0.0138 | 0.00748 | 0.00411 | 0.00681 | 0.00634 | 0.000412 | 0.00105 | 0.00265 | 0.00068 | 0.000152 | 6.5e-09 |
| Locus 123 RBCK1 | 9.05e-14 | 8.31e-14 | 0.000241 | 3.81e-13 | 1.69e-13 | 0.000105 | 0.000395 | 0.00308 | 0.00122 | 0.000385 | 0.000365 | 0.0072 | 0.0189 | 0.0479 | 0.000412 | 0.01 | 0.00356 | 0.00278 | 0.000271 | 5.4e-05 |
| Locus 126 CASS4 | 1.93e-19 | 1.11e-15 | 2.35e-06 | 9.25e-20 | 3.12e-16 | 1.12e-05 | 0.00183 | 0.00317 | 9.6e-05 | 0.00186 | 0.00185 | 0.000296 | 0.00138 | 0.000331 | 0.000912 | 0.000607 | 0.0028 | 0.000196 | 0.000555 | 0.000155 |
| Locus 127 APP | 1.88e-13 | 4.68e-12 | 4.86e-05 | 1.26e-13 | 1.28e-11 | 0.000134 | 0.000117 | 0.000359 | 0.00052 | 0.0158 | 0.00596 | 0.00074 | 0.00219 | 0.00364 | 0.000197 | 0.000473 | 0.00586 | 6.06e-05 | 3.3e-05 | 3e-07 |

**Supplementary Figure 7. The P-values of the known loci in the combined and stratified meta-analyses.** Red cells represent loci containing a significant variant ($P<5x10^{-8}$), orange cells represent loci containing a suggestive variant ($P<1x10^{-5}$). Male represents the no proxy all ancestry male only analysis; Female represents the no proxy all ancestry female only analysis. The max effective sample sizes are listed in brackets.

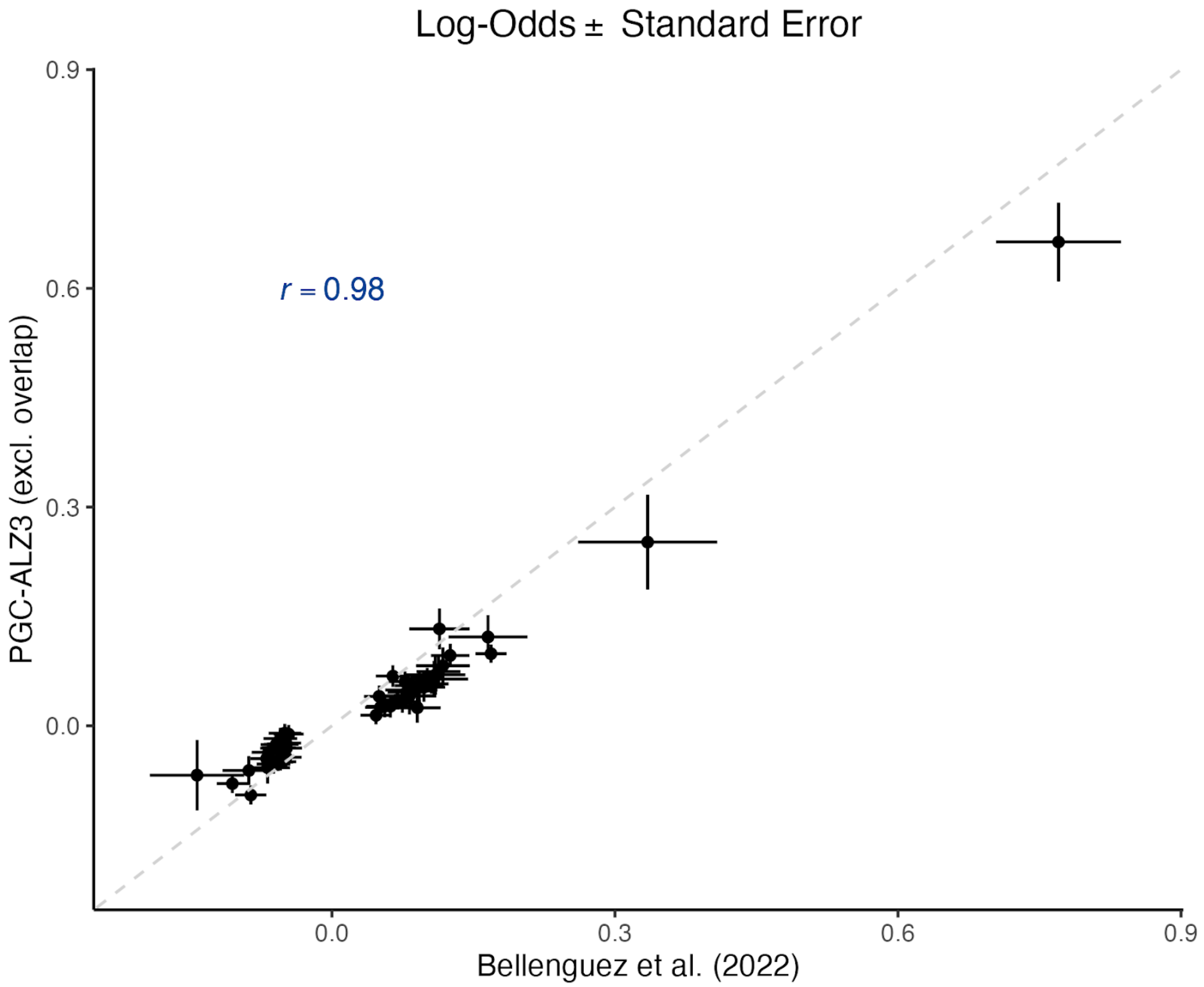

**Supplementary Figure 8. Correlation of effect sizes from an independent sample and Bellenguez et al. (2022).** We defined risk loci based on the publicly available summary statistics in Bellenguez et al. (2022). We correlated the effect sizes (Log-Odds) of the lead SNPs with the effect sizes in an independent meta-analysis (PGC-ALZ3 (excl. overlap)).

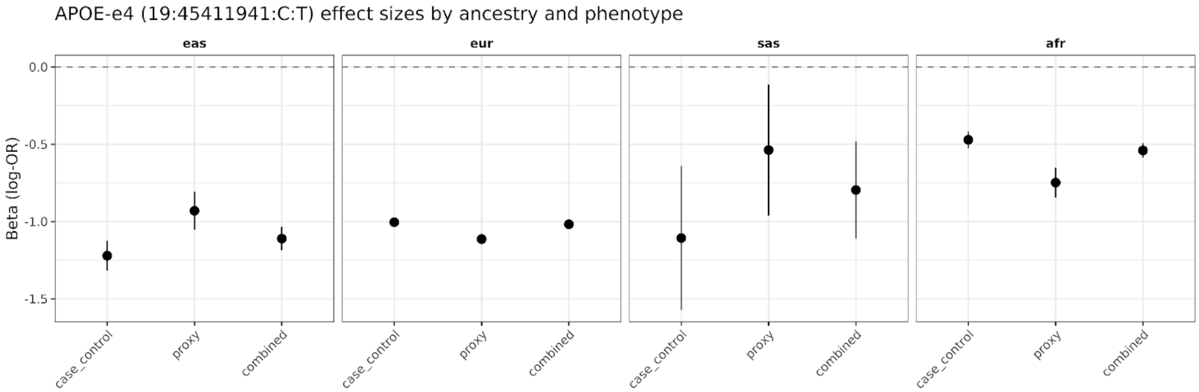

**Supplementary Figure 9. APOE-e4 effect sizes across ancestries and phenotype definitions.** Beta estimates (log odds ratios) with 95% confidence intervals for the APOE-ε4 tagging variant rs429358 (19:45411941:C:T) in case-control, proxy, and combined meta-analyses, shown separately for East Asian (EAS), European (EUR), South Asian (SAS), and African (AFR) ancestries. The protective direction reflects the T allele (non-ε4) as the effect allele; the ε4 risk allele is C.

**19:45411941:C:T**
**effect allele:T frequency:0.8214 neff:325,011**

| Cohort | log(OR) | Standard Error | P-value | 95%-CI | Weight |
|---|---|---|---|---|---|
| allof\|casec\|eur | NA | NA | NA | | 0.0% |
| biovu\|casec\|eur | -0.9734 | 0.0916 | 2.3767e-26 | -0.9734 [-1.1530; -0.7938] | 0.5% |
| decod\|casec\|eur | -1.0470 | 0.0236 | 0.0000e+00 | -1.0470 [-1.0931; -1.0008] | 7.3% |
| demge\|casec\|eur | -1.5158 | 0.0868 | 2.3871e-68 | -1.5158 [-1.6859; -1.3458] | 0.5% |
| eston\|casec\|eur | -1.1205 | 0.1092 | 1.0893e-24 | -1.1205 [-1.3346; -0.9064] | 0.3% |
| finng\|casec\|eur | -0.9730 | 0.0146 | 0.0000e+00 | -0.9730 [-1.0016; -0.9445] | 19.1% |
| genen\|casec\|eur | -1.6500 | 0.0486 | 1.1944e-252 | -1.6500 [-1.7453; -1.5547] | 1.7% |
| gothe\|casec\|eur | -1.0863 | 0.0907 | 4.5489e-33 | -1.0863 [-1.2640; -0.9085] | 0.5% |
| grace\|casec\|eur | -0.8201 | 0.0491 | 1.2588e-62 | -0.8201 [-0.9163; -0.7239] | 1.7% |
| lilly\|casec\|eur | -1.6297 | 0.0574 | 1.4762e-177 | -1.6297 [-1.7422; -1.5173] | 1.2% |
| rege1\|casec\|eur | -0.7777 | 0.0369 | 7.3742e-99 | -0.7777 [-0.8499; -0.7055] | 3.0% |
| rege2\|casec\|eur | -0.5214 | 0.1677 | 1.8722e-03 | -0.5214 [-0.8500; -0.1928] | 0.1% |
| rege3\|casec\|eur | -0.6850 | 0.0303 | 2.0374e-113 | -0.6850 [-0.7443; -0.6257] | 4.4% |
| rege4\|casec\|eur | -0.8280 | 0.1745 | 2.0824e-06 | -0.8280 [-1.1700; -0.4860] | 0.1% |
| rege5\|casec\|eur | -0.6564 | 0.0982 | 2.2741e-11 | -0.6564 [-0.8487; -0.4640] | 0.4% |
| rege6\|casec\|eur | -0.9941 | 0.1774 | 2.1063e-08 | -0.9941 [-1.3418; -0.6463] | 0.1% |
| rege7\|casec\|eur | -0.6774 | 0.0949 | 9.6843e-13 | -0.6774 [-0.8635; -0.4913] | 0.5% |
| twent\|casec\|eur | -1.2105 | 0.0262 | 0.0000e+00 | -1.2105 [-1.2618; -1.1592] | 5.9% |
| twing\|casec\|eur | -0.7860 | 0.1052 | 7.8430e-14 | -0.7860 [-0.9922; -0.5799] | 0.4% |
| vumcl\|casec\|eur | -1.2469 | 0.0455 | 1.4467e-165 | -1.2469 [-1.3360; -1.1578] | 2.0% |
| xchbb\|casec\|eur | -0.7537 | 0.0158 | 0.0000e+00 | -0.7537 [-0.7846; -0.7227] | 16.3% |
| xhunt\|casec\|eur | -1.2473 | 0.0382 | 2.0893e-234 | -1.2473 [-1.3221; -1.1725] | 2.8% |
| xhusk\|casec\|eur | -0.9493 | 0.0693 | 1.0353e-42 | -0.9493 [-1.0852; -0.8135] | 0.8% |
| xigap\|casec\|eur | -1.2017 | 0.0189 | 0.0000e+00 | -1.2017 [-1.2387; -1.1647] | 11.4% |
| xstsa\|casec\|eur | -0.7815 | 0.1274 | 8.5462e-10 | -0.7815 [-1.0312; -0.5318] | 0.3% |
| xukbb\|casec\|eur | -1.0794 | 0.0262 | 0.0000e+00 | -1.0794 [-1.1308; -1.0281] | 5.9% |
| allof\|proxy\|eur | NA | NA | NA | | 0.0% |
| life1\|proxy\|eur | -1.1912 | 0.1740 | 7.5956e-12 | -1.1912 [-1.5322; -0.8502] | 0.1% |
| life2\|proxy\|eur | -1.1363 | 0.6950 | 1.0206e-01 | -1.1363 [-2.4985; 0.2259] | 0.0% |
| life3\|proxy\|eur | NA | NA | NA | | 0.0% |
| xukbb\|proxy\|eur | -1.1122 | 0.0181 | 0.0000e+00 | -1.1122 [-1.1477; -1.0767] | 12.4% |
| **Common effect model** | | | | **-1.0175 [-1.0300; -1.0050]** | **100.0%** |

Heterogeneity: $I^2$ = 97.5%, $p$ = 8.1893e-210

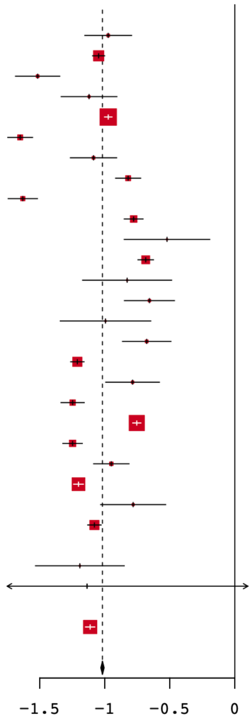

**Supplementary Figure 10. APOE-e4 effect sizes for individual European cohorts.** Forest plot of per-cohort log odds ratios with 95% confidence intervals for the APOE-ε4 tagging variant rs429358 (19:45411941:C:T; effect allele T, frequency 0.8214; effective sample size = 325,011) across case-control (casec) and proxy cohorts of European ancestry. Red squares are scaled by cohort sample size in the meta-analysis. The diamond at the bottom represents the common-effect pooled estimate (log OR = −1.02, 95% CI: −1.03 to −1.01). Substantial between-cohort heterogeneity is observed ($I^2$ = 97.5%, P = 8.19 × $10^{-210}$).

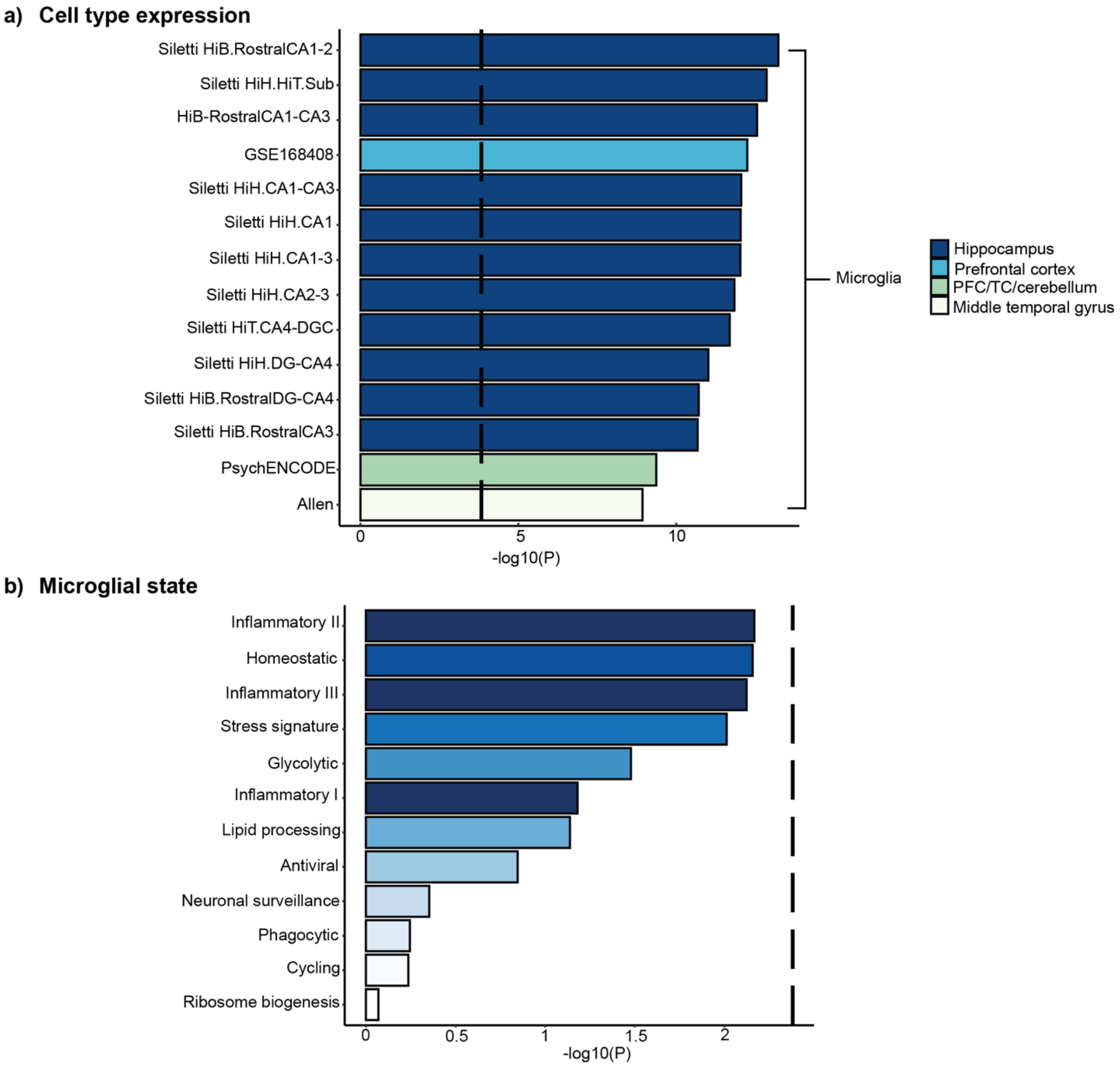

**Supplementary Figure 11. Microglia cell-type and state enrichment analysis.** The cell type analyses highlight cell types and cell states relevant for AD. **a)** The cell type enrichment analysis implemented in FUMA v1.7.0 identifies microglia as a cell type overexpressing AD-associated genes in multiple brain regions of controls. Only the significant cell types were visualized. **b)** Gene set association analysis does not find any Bonferroni-significant microglia states associated with the AD GWAS results. The dashed line represents Bonferroni significance after correcting for 12 microglia states (P<0.05/12).

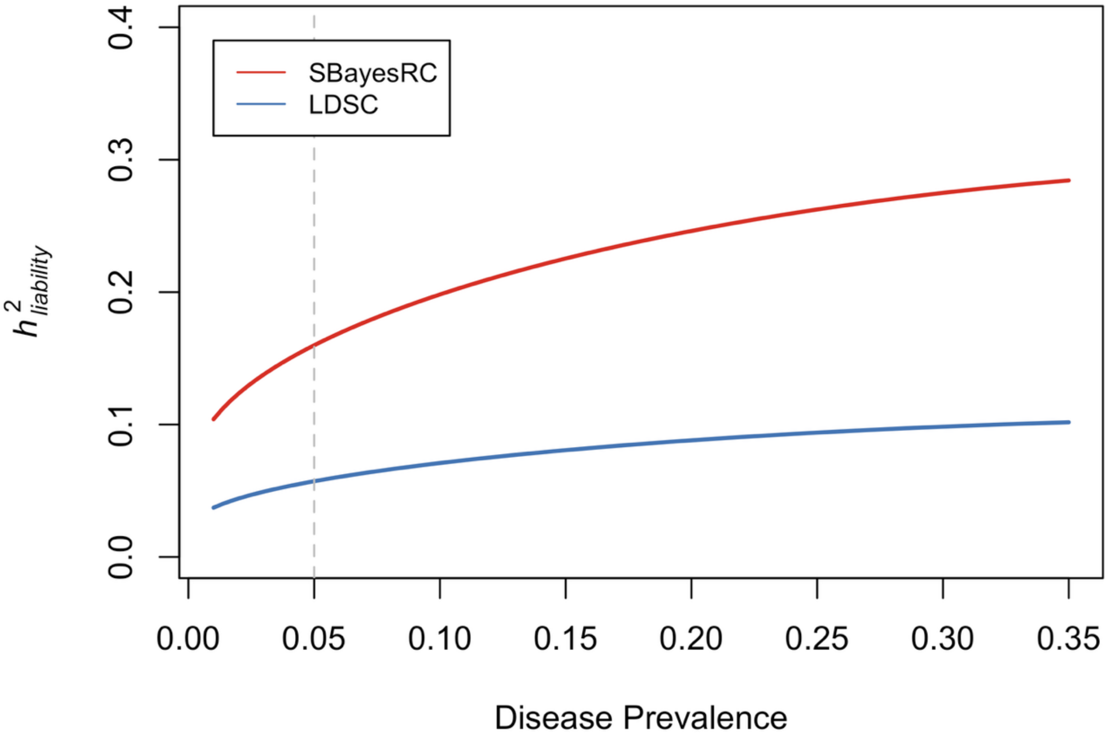

**Supplementary Figure 12. Heritability on the liability scale ($h^2_{liability}$) by disease prevalence.** $h^2_{liability}$ of AD (European and excluding proxy cases) as reported in Figure 4, but as a function of prevalence, using SBayesRC and LDSC.

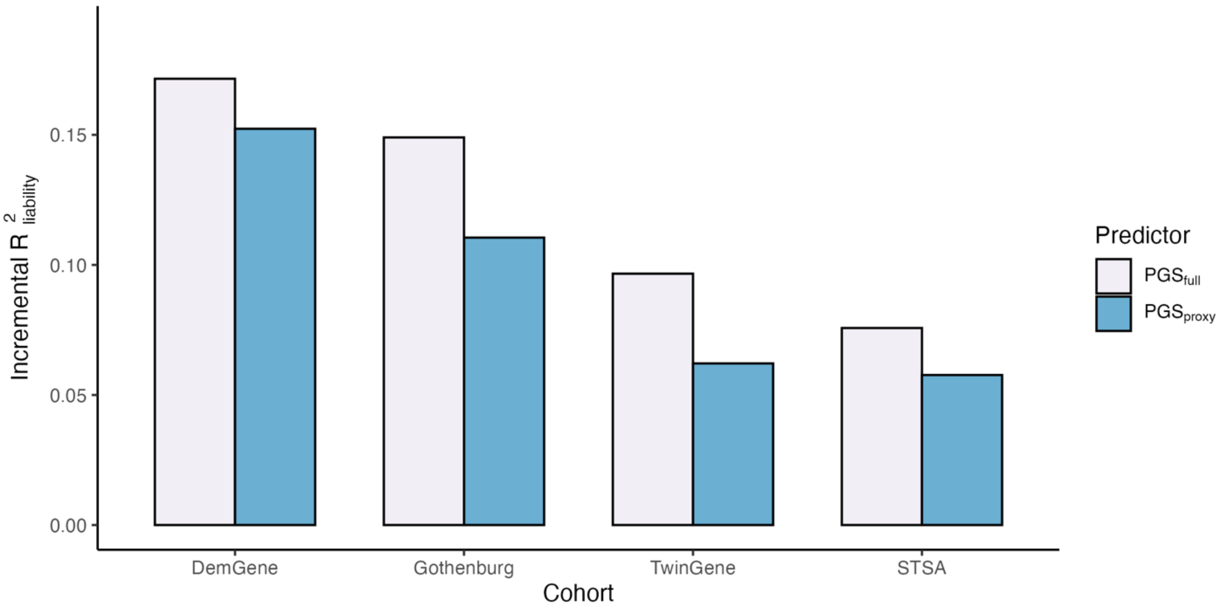

**Supplementary Figure 13. Comparison of Polygenic Scores (PGS) trained on the full GWAS versus a proxy-only GWAS.** The base model included the first 10 principal components, and the model to be evaluated additionally included the PGS. We computed the incremental $R^2_{liability}$ as the difference between the models. We applied a leave-one-cohort-out approach, in which the PGS for each testing cohort was constructed using GWAS summary statistics that excluded that cohort. $PGS_{full}$ was derived from the full meta-analysis (clinical and proxy cases), whereas $PGS_{proxy}$ was derived from a meta-analysis restricted to proxy cases.

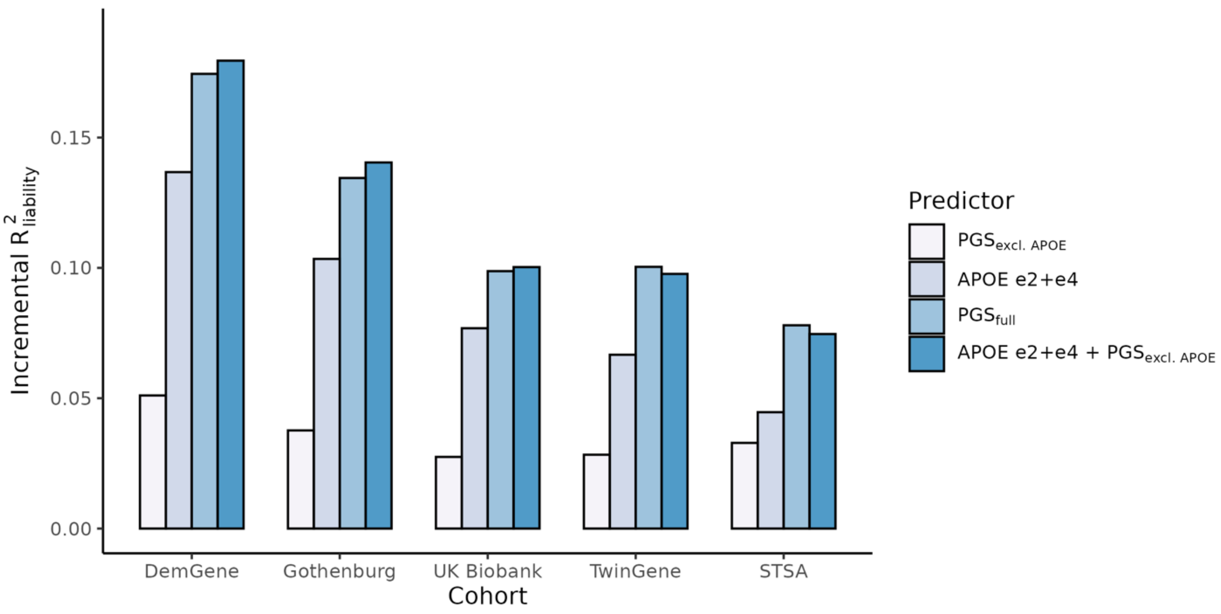

**Supplementary Figure 14. Predicting AD using SBayesRC-based Polygenic Scores (PGS) and APOE genotype counts.** The base model included the first 10 principal components, and the model to be evaluated additionally included the PGS. We computed the incremental $R^2_{liability}$ as the difference between the models. We applied a leave-one-cohort-out approach, in which the PGS for each testing cohort was constructed using GWAS summary statistics that excluded that cohort. For $PGS_{excl.\ APOE}$, we excluded all variants 5Mb downstream and upstream of position 19:45411941 on hg37.

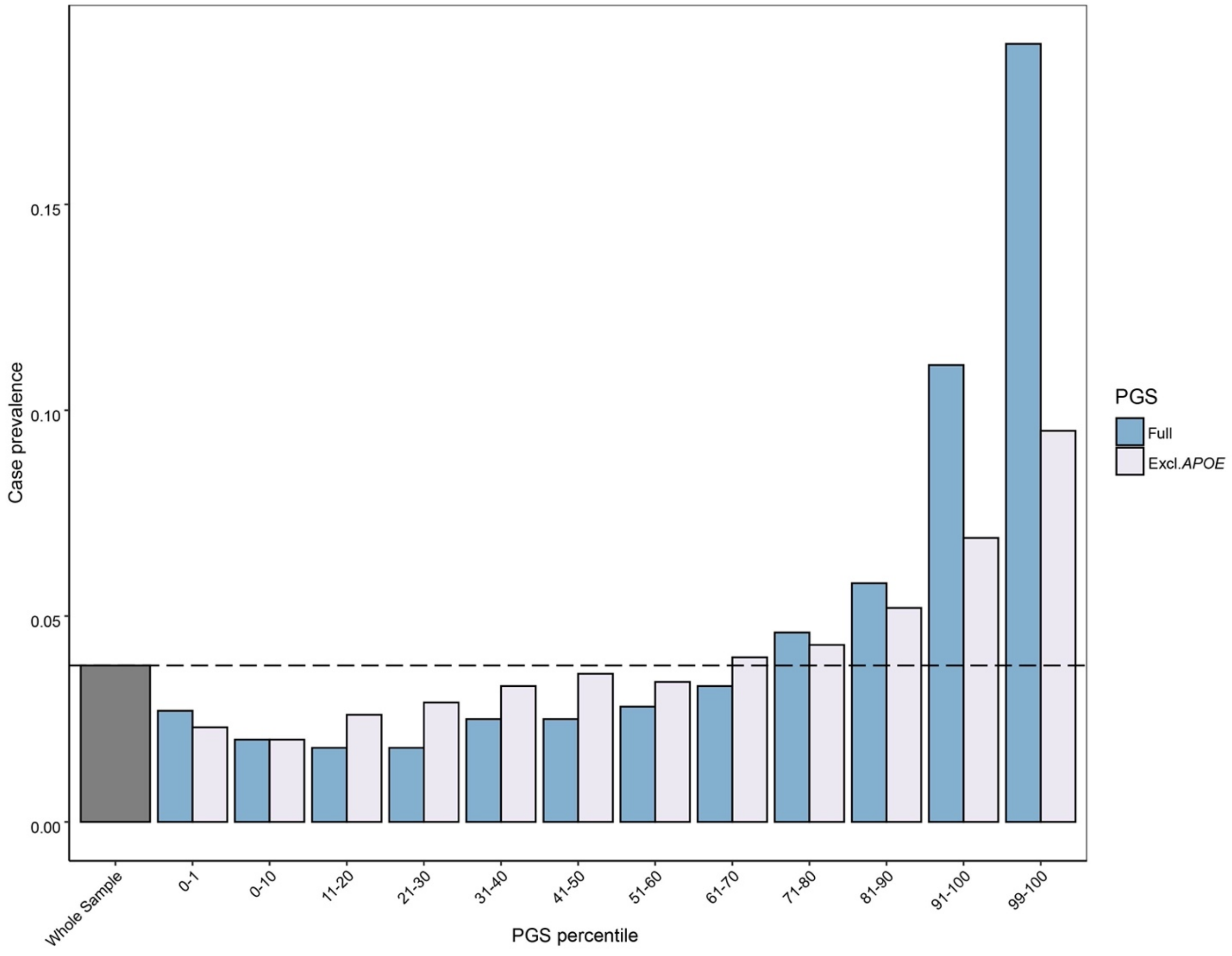

**Supplementary Figure 15. The number of cases and controls in each PGS percentile in unrelated EUR ancestry individuals 65 and older in the UKB.** The PGS model did not include UKB data. Excl.APOE represents the PGS with the *APOE* region excluded. For Excl.APOE, we excluded all variants 5Mb downstream and upstream of position 19:45411941 on hg37.

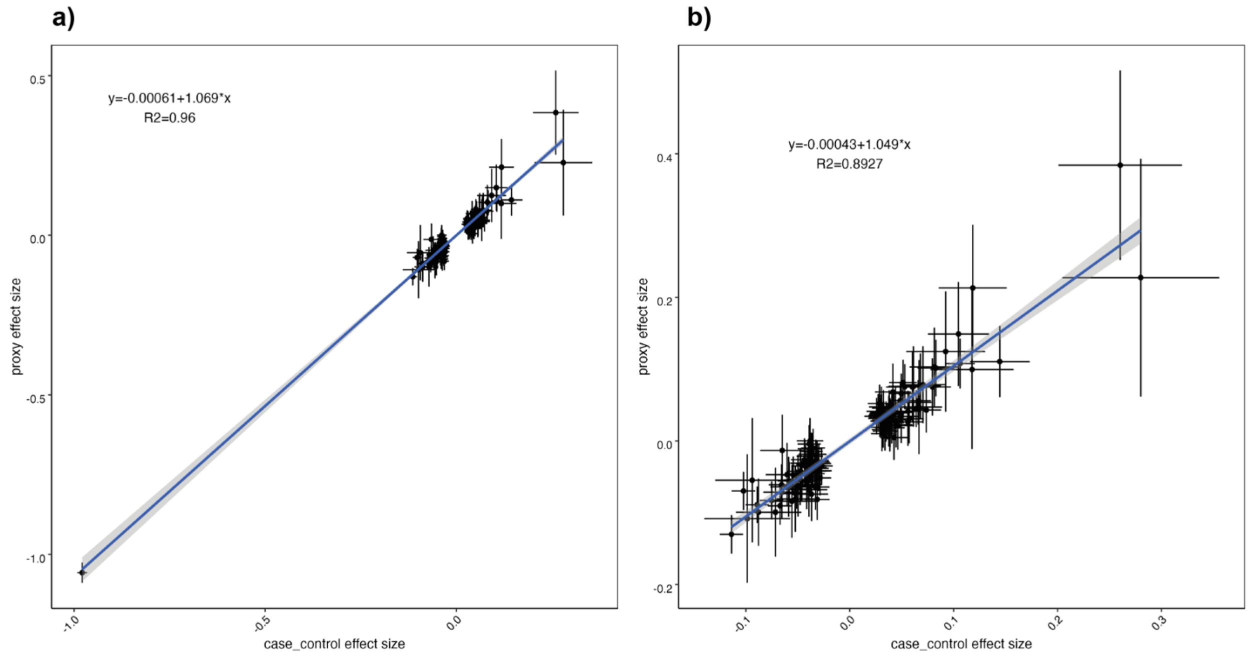

**Supplementary Figure 16. Comparison between the effect sizes observed in the case control only analysis and the proxy only analysis**. a) The effect sizes of the 124 overlapping lead SNPs across the case control only (case control) and proxy only meta-analyses. 2 lead SNPs (loci 24 and 58) were not present in the proxy data and 1 lead SNP (locus 105) was not present in the case control data. The error bars represent the 95% confidence interval. The line of best fit is coloured blue with the resulting formula displayed in the top left. b) The same plot as a but with the *APOE* region lead SNP removed.

### References

1. Wu, Y. *et al.* Pervasive biases in proxy genome-wide association studies based on parental history of Alzheimer's disease. *Nat. Genet.* 1–8 (2024) doi:10.1038/s41588-024-01963-9.
2. Wightman, D. P. *et al.* A genome-wide association study with 1,126,563 individuals identifies new risk loci for Alzheimer's disease. *Nat. Genet.* **53**, 1276–1282 (2021).
3. Bellenguez, C. *et al.* New insights into the genetic etiology of Alzheimer's disease and related dementias. *Nat. Genet.* **54**, 412–436 (2022).
4. Ma, C. *et al.* Identification and prediction of m7G-related Alzheimer's disease subtypes: insights from immune infiltration and machine learning models. *Front. Aging Neurosci.* **Volume 15-2023**, (2023).
5. Neuner, S. M., Ding, S. & Kaczorowski, C. C. Knockdown of heterochromatin protein 1 binding protein 3 recapitulates phenotypic, cellular, and molecular features of aging. *Aging Cell* **18**, e12886 (2019).
6. Salih, D. A. *et al.* Genetic variability in response to amyloid beta deposition influences Alzheimer's disease risk. *Brain Commun* **1**, fcz022 (2019).
7. Graham, A. C. *et al.* Human longevity and Alzheimer's disease variants act via microglia and oligodendrocyte gene networks. *Brain* **148**, 969–984 (2025).
8. Fominykh, V. *et al.* Shared genetic loci between Alzheimer's disease and multiple sclerosis: Crossroads between neurodegeneration and immune system. *Neurobiol. Dis.* **183**, 106174 (2023).
9. A European Multicentre Double-blind Placebo-controlled Phase III Trial of Nilvadipine in Mild to Moderate Alzheimer's Disease. (2013).
10. Lawlor, B. *et al.* Nilvadipine in mild to moderate Alzheimer disease: A randomised controlled trial. *PLOS Med.* **15**, e1002660 (2018).

11. Kreft, K. L. *et al.* Abundant kif21b is associated with accelerated progression in neurodegenerative diseases. *Acta Neuropathol. Commun.* **2**, 144 (2014).

12. Hares, K. *et al.* Overexpression of Kinesin Superfamily Motor Proteins in Alzheimer's Disease. *J. Alzheimer's Dis.* **60**, 1511–1524 (2017).

13. Asselin, L. *et al.* Mutations in the KIF21B kinesin gene cause neurodevelopmental disorders through imbalanced canonical motor activity. *Nat. Commun.* **11**, 2441 (2020).

14. Lyons, C. E. *et al.* Lifelong chronic psychosocial stress induces a proteomic signature of Alzheimer's disease in wildtype mice. *Eur. J. Neurosci.* **55**, 2971–2985 (2022).

15. Li, Z. *et al.* From genes to drugs: targeting Alzheimer's with circadian insights. *Front. Aging Neurosci.* **Volume 17-2025**, (2025).

16. Abdullah, M. N., Wah, Y. B., Abdul Majeed, A. B., Zakaria, Y. & Shaadan, N. Identification of blood-based transcriptomics biomarkers for Alzheimer's disease using statistical and machine learning classifier. *Inform. Med. Unlocked* **33**, 101083 (2022).

17. Andreev, V. P. *et al.* Label-Free Quantitative LC–MS Proteomics of Alzheimer's Disease and Normally Aged Human Brains. *J. Proteome Res.* **11**, 3053–3067 (2012).

18. Chen, D., Wang, X., Huang, T. & Jia, J. Sleep and Late-Onset Alzheimer's Disease: Shared Genetic Risk Factors, Drug Targets, Molecular Mechanisms, and Causal Effects. *Front. Genet.* **Volume 13-2022**, (2022).

19. Jang, Y.-N. *et al.* RAPGEF2 mediates oligomeric Aβ-induced synaptic loss and cognitive dysfunction in the 3xTg-AD mouse model of Alzheimer's disease. *Neuropathol Appl Neurobiol* **47**, 625–639 (2021).

20. Zhang, H. *et al.* The Retromer Complex and Sorting Nexins in Neurodegenerative Diseases. *Front. Aging Neurosci.* **10**, (2018).

21. Fan, C. C. *et al.* Sex-dependent autosomal effects on clinical progression of Alzheimer's disease. *Brain* **143**, 2272–2280 (2020).

22. Butler, C. A. *et al.* The Abca7V1613M variant reduces Aβ generation, plaque load, and neuronal damage. *Alzheimers Dement.* **20**, 4914–4934 (2024).

23. Peng, Q., Long, C. L., Malhotra, S. & Humphrey, M. B. A Physical Interaction Between the Adaptor Proteins DOK3 and DAP12 Is Required to Inhibit Lipopolysaccharide Signaling in Macrophages. *Sci. Signal.* **6**, ra72–ra72 (2013).

24. Cai, X., Xing, J., Long, C. L., Peng, Q. & Humphrey, M. B. DOK3 Modulates Bone Remodeling by Negatively Regulating Osteoclastogenesis and Positively Regulating Osteoblastogenesis. *J. Bone Miner. Res.* **32**, 2207–2218 (2017).

25. Lin, H. *et al.* Related Genes and Potential Biomarkers for Early Diagnosis of Alzheimer's Disease: A Preliminary Study Based on DNA Microarray. *Am. J. Alzheimers Dis. Dementias®* **29**, 90–95 (2014).

26. Chung, J. *et al.* Genome-wide pleiotropy analysis of neuropathological traits related to Alzheimer's disease. *Alzheimers Res. Ther.* **10**, 22 (2018).

27. Zhang, H.-L. *et al.* HDAC9-mediated calmodulin deacetylation induces memory impairment in Alzheimer's disease. *CNS Neurosci. Ther.* **30**, e14573 (2024).

28. Bone, W. P. *et al.* Multi-trait association studies discover pleiotropic loci between Alzheimer's disease and cardiometabolic traits. *Alzheimers Res. Ther.* **13**, 34 (2021).

29. Brokate-Llanos, A. M. *et al.* Ribonucleotide reductase inhibition improves the symptoms of a Caenorhabditis elegans model of Alzheimer's disease. *G3 GenesGenomesGenetics* **14**, jkae040 (2024).

30. Schweig, J. E. *et al.* Alzheimer's disease pathological lesions activate the spleen tyrosine kinase. *Acta Neuropathol. Commun.* **5**, 69 (2017).

31. Ennerfelt, H. *et al.* SYK coordinates neuroprotective microglial responses in neurodegenerative disease. *Cell* **185**, 4135-4152.e22 (2022).

32. Stuchbury, G., Ko, A. & Dymock, B. A brain penetrant small-molecule SYK inhibitor for the treatment of Alzheimer's and neuroinflammatory diseases. *Alzheimers Dement.* **19**, e082900 (2023).

33. Wang, S. *et al.* TREM2 drives microglia response to amyloid-β via SYK-dependent and -independent pathways. *Cell* **185**, 4153-4169.e19 (2022).

34. Kang, D. E. & Woo, J. A. Cofilin, a Master Node Regulating Cytoskeletal Pathogenesis in Alzheimer's Disease. *J Alzheimers Dis* **72**, S131–S144 (2019).

35. Liu, N. *et al.* Characterization of gene expression profiles in Alzheimer's disease and osteoarthritis: A bioinformatics study. *PLOS ONE* **20**, e0316708 (2025).

36. Pyun, J.-M. *et al.* Transcriptional risk scores in Alzheimer's disease: From pathology to cognition. *Alzheimers Dement.* **20**, 243–252 (2024).

37. Kosoy, R. *et al.* Genetics of the human microglia regulome refines Alzheimer's disease risk loci. *Nat. Genet.* **54**, 1145–1154 (2022).

38. Sase, S., Takanohashi, A., Vanderver, A. & Almad, A. Astrocytes, an active player in Aicardi–Goutières syndrome. *Brain Pathol.* **28**, 399–407 (2018).

39. Polanco, J. C. *et al.* CRISPRi screening reveals regulators of tau pathology shared between exosomal and vesicle-free tau. *Life Sci. Alliance* **6**, e202201689 (2023).

40. Prissette, M. *et al.* Disruption of nuclear envelope integrity as a possible initiating event in tauopathies. *Cell Rep.* **40**, (2022).

41. Lin, K.-P. *et al.* Genetic polymorphisms of a novel vascular susceptibility gene, Ninjurin2 (NINJ2), are associated with a decreased risk of Alzheimer's disease. *PLoS One* **6**, e20573 (2011).

42. Liu, S. *et al.* SLAMF8 and NINJ2 promote neuroinflammation and oxidative stress through TLR4 NF kappa B pathway in Alzheimer's disease. *Sci. Rep.* **15**, 17501 (2025).

43. Gong, Y. *et al.* Stereo-seq of the prefrontal cortex in aging and Alzheimer's disease. *Nat. Commun.* **16**, 482 (2025).

44. vom Berg, J. *et al.* Inhibition of IL-12/IL-23 signaling reduces Alzheimer's disease–like pathology and cognitive decline. *Nat. Med.* **18**, 1812–1819 (2012).

45. Schneeberger, S. *et al.* Interleukin-12 signaling drives Alzheimer's disease pathology through disrupting neuronal and oligodendrocyte homeostasis. *Nat. Aging* **5**, 622–641 (2025).

46. Llera-Oyola, J. *et al.* The role of microRNAs in understanding sex-based differences in Alzheimer's disease. *Biol. Sex Differ.* **15**, 13 (2024).

47. Rahman, Md. R. *et al.* Identification of molecular signatures and pathways to identify novel therapeutic targets in Alzheimer's disease: Insights from a systems biomedicine perspective. *Genomics* **112**, 1290–1299 (2020).

48. Alves, V. C., Carro, E. & Figueiro-Silva, J. Unveiling DNA methylation in Alzheimer's disease: a review of array-based human brain studies. *Neural Regen. Res.* **19**, (2024).

49. Whyte, L. S. *et al.* Lysosomal Dysregulation in the Murine AppNL-G-F/NL-G-F Model of Alzheimer's Disease. *Neuroscience* **429**, 143–155 (2020).

50. Hashimoto, T. *et al.* Age-dependent increase in lysosome-associated membrane protein 1 and early-onset behavioral deficits in APPSL transgenic mouse model of Alzheimer's disease. *Neurosci. Lett.* **469**, 273–277 (2010).

51. Chang, W.-S., Wang, Y.-H., Zhu, X.-T. & Wu, C.-J. Genome-Wide Profiling of miRNA and mRNA Expression in Alzheimer's Disease. *Med. Sci. Monit. Int. Med. J. Exp. Clin. Res.* **23**, 2721–2731 (2017).

52. Ozsan McMillan, I., Li, J.-P. & Wang, L. Heparan sulfate proteoglycan in Alzheimer's disease: aberrant expression and functions in molecular pathways related to amyloid-β metabolism. *Am. J. Physiol.-Cell Physiol.* **324**, C893–C909 (2023).

53. Kang, M. *et al.* Whole-genome sequencing study in Koreans identifies novel loci for Alzheimer's disease. *Alzheimers Dement* **20**, 8246–8262 (2024).

54. Einem, B. von *et al.* The Golgi-Localized γ-Ear-Containing ARF-Binding (GGA) Proteins Alter Amyloid-β Precursor Protein (APP) Processing through Interaction of Their GAE Domain with the Beta-Site APP Cleaving Enzyme 1 (BACE1). *PLOS ONE* **10**, e0129047 (2015).
55. Carter, C. Alzheimer's Disease: APP, Gamma Secretase, APOE, CLU, CR1, PICALM, ABCA7, BIN1, CD2AP, CD33, EPHA1, and MS4A2, and Their Relationships with Herpes Simplex, C. Pneumoniae, Other Suspect Pathogens, and the Immune System. *Int. J. Alzheimer's Dis.* **2011**, 501862 (2011).
56. Sandusky-Beltran, L. A. *et al.* Spermidine/spermine-N1-acetyltransferase ablation impacts tauopathy-induced polyamine stress response. *Alzheimers Res. Ther.* **11**, 58 (2019).
57. Park, K.-R. *et al.* Prevention of multiple system atrophy using human bone marrow-derived mesenchymal stem cells by reducing polyamine and cholesterol-induced neural damages. *Stem Cell Res. Ther.* **11**, 63 (2020).
58. Jansen, I. E. *et al.* Genome-wide meta-analysis for Alzheimer's disease cerebrospinal fluid biomarkers. *Acta Neuropathol. (Berl.)* **144**, 821–842 (2022).
59. Pottier, C. *et al.* Shared brain transcriptomic signature in TDP-43 type A FTLD patients with or without GRN mutations. *Brain* **145**, 2472–2485 (2022).
60. Kamagata, E. *et al.* Decrease of dynamin 2 levels in late-onset Alzheimer's disease alters Abeta metabolism. *Biochem Biophys Res Commun* **379**, 691–695 (2009).
61. Aidaralieva, N. J. *et al.* Dynamin 2 gene is a novel susceptibility gene for late-onset Alzheimer disease in non-APOE-ε4 carriers. *J. Hum. Genet.* **53**, 296–302 (2008).
62. Fakhri, S. *et al.* Attenuation of Nrf2/Keap1/ARE in Alzheimer's Disease by Plant Secondary Metabolites: A Mechanistic Review. *Molecules* **25**, (2020).
63. Kerr, F. *et al.* Direct Keap1-Nrf2 disruption as a potential therapeutic target for Alzheimer's disease. *PLOS Genet.* **13**, e1006593 (2017).

64. Devadoss, D., Akkaoui, J., Nair, M. & Lakshmana, M. K. LRRC25 expression during physiological aging and in mouse models of Alzheimer's disease and iPSC-derived neurons. *Front. Mol. Neurosci.* **17**, 1365752 (2024).

65. Mamchur, A. *et al.* Cognitive Impairment in Nonagenarians: Potential Metabolic Mechanisms Revealed by the Synergy of In Silico Gene Expression Modeling and Pathway Enrichment Analysis. *Int. J. Mol. Sci.* **25**, (2024).

66. Aladeokin, A. C. *et al.* Network-guided analysis of hippocampal proteome identifies novel proteins that colocalize with Aβ in a mice model of early-stage Alzheimer's disease. *Neurobiol. Dis.* **132**, 104603 (2019).

67. Kim, S.-M. *et al.* TREM2 promotes Aβ phagocytosis by upregulating C/EBPα-dependent CD36 expression in microglia. *Sci. Rep.* **7**, 11118 (2017).

68. Wilson, E. N. & Andreasson, K. I. TAM-ping down amyloid in Alzheimer's disease. *Nat. Immunol.* **22**, 543–544 (2021).

69. Chen, G., Ozturk, G., Porta, S. & Lee, V. M.-Y. Evaluate the Efficacy of the Axl inhibitor Bemcentinib in the 5xFAD mouse model of Alzheimer's Disease. *Alzheimers Dement.* **20**, e089525 (2024).

70. Mattsson, N. *et al.* CSF protein biomarkers predicting longitudinal reduction of CSF β-amyloid42 in cognitively healthy elders. *Transl. Psychiatry* **3**, e293–e293 (2013).

71. Owlett, L., Olschowka, J. A., Elliott, M. R. & O'Banion, M. K. AXL activation leads to reduced amyloid plaque deposition in APP/PS-1 mice. *Alzheimers Dement.* **16**, e046330 (2020).

72. Huang, Y. *et al.* Microglia use TAM receptors to detect and engulf amyloid β plaques. *Nat. Immunol.* **22**, 586–594 (2021).

73. Wyss-Coray, T. *et al.* TGF-β1 promotes microglial amyloid-β clearance and reduces plaque burden in transgenic mice. *Nat. Med.* **7**, 612–618 (2001).

74. Lesné, S. *et al.* Transforming Growth Factor-β1 Potentiates Amyloid-β Generation in Astrocytes and in Transgenic Mice *. *J. Biol. Chem.* **278**, 18408–18418 (2003).

75. Han, S.-W. *et al.* Transcriptome analysis of early- and late-onset Alzheimer's disease in Korean cohorts. *Alzheimers Dement.* **21**, e14563 (2025).

76. Chen, W., Zhang, T. & Zhang, H. Genes related to neurotransmitter receptors as potential biomarkers for Alzheimer's disease. *Neurosci. Lett.* **832**, 137816 (2024).

77. Gallart-Palau, X. *et al.* Brain-derived and circulating vesicle profiles indicate neurovascular unit dysfunction in early Alzheimer's disease. *Brain Pathol.* **29**, 593–605 (2019).

78. Xu, N. *et al.* Ube2v1 Positively Regulates Protein Aggregation by Modulating Ubiquitin Proteasome System Performance Partially Through K63 Ubiquitination. *Circ. Res.* **126**, 907–922 (2020).

79. Liu, H. *et al.* Ubiquitin–proteasome system in the different stages of dominantly inherited Alzheimer's disease. *Alzheimers Dement.* **21**, e70243 (2025).

80. Masola, V., Greco, N., Tozzo, P., Caenazzo, L. & Onisto, M. The role of SPATA2 in TNF signaling, cancer, and spermatogenesis. *Cell Death Dis.* **13**, 977 (2022).

81. Chen, K. *et al.* Identifying risk loci for FTD and shared genetic component with ALS: A large-scale multitrait association analysis. *Neurobiol. Aging* **134**, 28–39 (2024).

82. de Rojas, I. *et al.* Common variants in Alzheimer's disease and risk stratification by polygenic risk scores. *Nat. Commun.* **12**, 3417 (2021).

83. Liu, Y., Shi, Y. & Wang, P. Functions of glutaminyl cyclase and its isoform in diseases. *Vis Cancer Med* **4**, (2023).

84. Bayer, T. A. Pyroglutamate Aβ cascade as drug target in Alzheimer's disease. *Mol. Psychiatry* **27**, 1880–1885 (2022).

85. He, S., Xu, Z. & Han, X. Lipidome disruption in Alzheimer's disease brain: detection, pathological mechanisms, and therapeutic implications. *Mol. Neurodegener.* **20**, 11 (2025).

86. Qu, H.-Q. *et al.* Risk of Alzheimer's disease in Down syndrome: Insights gained by multi-omics. *Alzheimers Dement.* **21**, e14604 (2025).

87. Yu, Y. & Martins, L. M. Mitochondrial One-Carbon Metabolism and Alzheimer's Disease. *Int. J. Mol. Sci.* **25**, (2024).

88. Lake, J. *et al.* Multi-ancestry meta-analysis and fine-mapping in Alzheimer's disease. *Mol. Psychiatry* 1–12 (2023) doi:10.1038/s41380-023-02089-w.

89. Nagle, M. W. *et al.* The 4p16.3 Parkinson Disease Risk Locus Is Associated with GAK Expression and Genes Involved with the Synaptic Vesicle Membrane. *PLOS ONE* **11**, e0160925 (2016).

90. Wang, J. *et al.* Distinct effects of SDC3 and FGFRL1 on selective neurodegeneration in AD and PD. *FASEB J.* **37**, e22773 (2023).

91. Alam, R. *et al.* New insights into the role of fibroblast growth factors in Alzheimer's disease. *Mol. Biol. Rep.* **49**, 1413–1427 (2022).

92. Benitez, B. A. *et al.* Haploinsufficiency of lysosomal enzyme genes in Alzheimer's disease. *bioRxiv* 2024.11.16.623962 (2024) doi:10.1101/2024.11.16.623962.

93. Schwartzentruber, J. *et al.* Genome-wide meta-analysis, fine-mapping and integrative prioritization implicate new Alzheimer's disease risk genes. *Nat. Genet.* **53**, 392–402 (2021).

94. Zhao, J. *et al.* 3-O-Sulfation of Heparan Sulfate Enhances Tau Interaction and Cellular Uptake. *Angew. Chem. Int. Ed.* **59**, 1818–1827 (2020).

95. Xu, M. *et al.* Clnk plays a role in TNF-alpha-induced cell death in murine fibrosarcoma cell line L929. *Biochem. Biophys. Res. Commun.* **463**, 275–279 (2015).

96. Tesi, N. *et al.* Cognitively healthy centenarians are genetically protected against Alzheimer's disease. *Alzheimers Dement.* **20**, 3864–3875 (2024).

97. Zhao, G. *et al.* Activation of nuclear factor-kappa B accelerates vascular calcification by inhibiting ankylosis protein homolog expression. *Kidney Int.* **82**, 34–44 (2012).

98. Hao, S., Wang, R., Zhang, Y. & Zhan, H. Prediction of Alzheimer's Disease-Associated Genes by Integration of GWAS Summary Data and Expression Data. *Front. Genet.* **Volume 9-2018**, (2019).

99. Tangavelou, K. *et al.* The deubiquitinase OTULIN regulates tau expression and RNA metabolism in neurons. *bioRxiv* 2025.04.09.648063 (2025) doi:10.1101/2025.04.09.648063.

100. Vromen, E. M. *et al.* CSF proteomic signature predicts progression to Alzheimer's disease dementia. *Alzheimers Dement. Transl. Res. Clin. Interv.* **8**, e12240 (2022).

101. Thiex, R. *et al.* A Novel Association between RASA1 Mutations and Spinal Arteriovenous Anomalies. *Am. J. Neuroradiol.* **31**, 775 (2010).

102. Xue, W., Li, J., Fu, K. & Teng, W. Differential Expression of mRNAs in Peripheral Blood Related to Prodrome and Progression of Alzheimer's Disease. *BioMed Res. Int.* **2020**, 4505720 (2020).

103. Niu, Y., Meng, J., Xue, Z. & Chen, Z. PSMA3-AS1: a promising LncRNA as a diagnostic and prognostic biomarker in human cancers. *Gene* **960**, 149521 (2025).

104. Glover, J. *et al.* UMAD1 contributes to ESCRT-III dynamic subunit turnover during cytokinetic abscission. *J. Cell Sci.* **136**, jcs261097 (2023).

105. Ji, L. *et al.* ICA1 affects APP processing through the PICK1-PKCα signaling pathway. *CNS Neurosci. Ther.* **30**, e14754 (2024).

106. Lambert, J.-C. *et al.* Meta-analysis of 74,046 individuals identifies 11 new susceptibility loci for Alzheimer's disease. *Nat. Genet.* **45**, 1452–1458 (2013).

107. Chen, Y. *et al.* Microglia efferocytosis: an emerging mechanism for the resolution of neuroinflammation in Alzheimer's disease. *J. Neuroinflammation* **22**, 96 (2025).

108. Srinivasan, K. *et al.* Alzheimer's Patient Microglia Exhibit Enhanced Aging and Unique Transcriptional Activation. *Cell Rep.* **31**, (2020).

109. Sawabe, A., Okazaki, S., Nakamura, A., Goitsuka, R. & Kaifu, T. The orphan G protein–coupled receptor 141 expressed in myeloid cells functions as an inflammation suppressor. *J. Leukoc. Biol.* **115**, 935–945 (2024).

110. Liu, S.-L. *et al.* NME8 rs2718058 polymorphism with Alzheimer's disease risk: a replication and meta-analysis. *Oncotarget Vol 7 No 24* https://www.oncotarget.com/article/9086/text/ (2016).

111. Liu, Y. *et al.* Association between NME8 Locus Polymorphism and Cognitive Decline, Cerebrospinal Fluid and Neuroimaging Biomarkers in Alzheimer's Disease. *PLOS ONE* **9**, e114777 (2014).

112. Li, M. *et al.* Dysregulated gene-associated biomarkers for Alzheimer's disease and aging. **12**, 83–95 (2021).

113. Feng, Y. *et al.* Widespread transposable element dysregulation in human aging brains with Alzheimer's disease. *Alzheimers Dement.* **20**, 7495–7517 (2024).

114. Papuć, E. & Rejdak, K. The role of myelin damage in Alzheimer's disease pathology. *Arch Med Sci* **16**, 345–341 (2020).

115. Hook, V. *et al.* Cathepsin B in neurodegeneration of Alzheimer's disease, traumatic brain injury, and related brain disorders. *Biochim. Biophys. Acta BBA - Proteins Proteomics* **1868**, 140428 (2020).

116. Hook, G. *et al.* Cathepsin B Gene Knockout Improves Behavioral Deficits and Reduces Pathology in Models of Neurologic Disorders. *Pharmacol. Rev.* **74**, 600–629 (2022).

117. Dalmasso, M. C. *et al.* First GWAS on Alzheimer's Disease in Argentina and Chile populations. 2023.01.16.23284609 Preprint at https://doi.org/10.1101/2023.01.16.23284609 (2023).

118. Uhrig, M. *et al.* New Alzheimer Amyloid β Responsive Genes Identified in Human Neuroblastoma Cells by Hierarchical Clustering. *PLOS ONE* **4**, e6779 (2009).

119. Rodriguez, O. L. *et al.* Genetic variation in the immunoglobulin heavy chain locus shapes the human antibody repertoire. *Nat. Commun.* **14**, 4419 (2023).

120. Fass, S. B. *et al.* Relationship between sex biases in gene expression and sex biases in autism and Alzheimer's disease. *Biol. Sex Differ.* **15**, 47 (2024).

121. Donner, L. *et al.* Platelets contribute to amyloid-β aggregation in cerebral vessels through integrin αIIbβ3–induced outside-in signaling and clusterin release. *Sci. Signal.* **9**, ra52–ra52 (2016).

122. Rhinn, H., Tatton, N., McCaughey, S., Kurnellas, M. & Rosenthal, A. Progranulin as a therapeutic target in neurodegenerative diseases. *Trends Pharmacol. Sci.* **43**, 641–652 (2022).

123. Kuo, M.-L. *et al.* RRM2B Suppresses Activation of the Oxidative Stress Pathway and is Up-regulated by P53 During Senescence. *Sci. Rep.* **2**, 822 (2012).

124. van Rheenen, W. *et al.* Common and rare variant association analyses in amyotrophic lateral sclerosis identify 15 risk loci with distinct genetic architectures and neuron-specific biology. *Nat. Genet.* **53**, 1636–1648 (2021).

125. Nalls, M. A. *et al.* Identification of novel risk loci, causal insights, and heritable risk for Parkinson's disease: a meta-analysis of genome-wide association studies. *Lancet Neurol.* **18**, 1091–1102 (2019).

126. Giambartolomei, C. *et al.* Bayesian test for colocalisation between pairs of genetic association studies using summary statistics. *PLoS Genet.* **10**, e1004383 (2014).

127. Werme, J., van der Sluis, S., Posthuma, D. & de Leeuw, C. A. An integrated framework for local genetic correlation analysis. *Nat. Genet.* **54**, 274–282 (2022).

128. Verheijen, J. & Sleegers, K. Understanding Alzheimer Disease at the Interface between Genetics and Transcriptomics. *Trends Genet.* **34**, 434–447 (2018).

129. Cissé, M. & Checler, F. Eph receptors: New players in Alzheimer's disease pathogenesis. *Neurobiol. Dis.* **73**, 137–149 (2015).

130. Pocernich, C. B. & Butterfield, D. A. Elevation of glutathione as a therapeutic strategy in Alzheimer disease. *Biochim. Biophys. Acta BBA - Mol. Basis Dis.* **1822**, 625–630 (2012).

131. Laskovs, M., Partridge, L. & Slack, C. Molecular inhibition of RAS signalling to target ageing and age-related health. *Dis. Model. Mech.* **15**, dmm049627 (2022).

132. Li, S. & Kim, H.-E. Implications of Sphingolipids on Aging and Age-Related Diseases. *Front. Aging* **2**, (2022).

133. Musardo, S. *et al.* The development of ADAM10 endocytosis inhibitors for the treatment of Alzheimer's disease. *Mol. Ther.* **30**, 2474–2490 (2022).

134. Sun, N. *et al.* Human microglial state dynamics in Alzheimer's disease progression. *Cell* **186**, 4386-4403.e29 (2023).

135. Cheng, J. *et al.* Early glycolytic reprogramming controls microglial inflammatory activation. *J. Neuroinflammation* **18**, 129 (2021).

136. Rodríguez-Giraldo, M. *et al.* Astrocytes as a Therapeutic Target in Alzheimer's Disease–Comprehensive Review and Recent Developments. *Int. J. Mol. Sci.* **23**, 13630 (2022).

137. Kim, M. *et al.* Maf links Neuregulin1 signaling to cholesterol synthesis in myelinating Schwann cells. *Genes Dev.* **32**, 645–657 (2018).

138. Mathys, H. *et al.* Single-cell transcriptomic analysis of Alzheimer's disease. *Nature* **570**, 332–337 (2019).

139. Xu, W. *et al.* Mbnl1 Protects Against Cerebral Ischemia–Reperfusion Injury by Modulating Microglia/Macrophage Polarization via NF-κB Pathway. *Mol. Neurobiol.* **62**, 13899–13916 (2025).

140. Li, X., Long, J., He, T., Belshaw, R. & Scott, J. Integrated genomic approaches identify major pathways and upstream regulators in late onset Alzheimer's disease. *Sci. Rep.* **5**, 12393 (2015).

141. Sierksma, A. *et al.* Novel Alzheimer risk genes determine the microglia response to amyloid-β but not to TAU pathology. *EMBO Mol. Med.* **12**, EMMM201910606 (2020).

142. Kozlova, A. *et al.* PICALM Alzheimer's risk allele causes aberrant lipid droplets in microglia. *Nature* **646**, 1178–1186 (2025).

143. Kimura, K. *et al.* Immune checkpoint TIM-3 regulates microglia and Alzheimer's disease. *Nature* **641**, 718–731 (2025).

144. Jaworski, M. & Thome, M. The paracaspase MALT1: biological function and potential for therapeutic inhibition. *Cell. Mol. Life Sci.* **73**, 459–473 (2016).

145. Serneels, L. *et al.* γ-Secretase Heterogeneity in the Aph1 Subunit: Relevance for Alzheimer's Disease. *Science* **324**, 639–642 (2009).

146. Lim, F. T., Ogawa, S. & Parhar, I. S. Spred-2 expression is associated with neural repair of injured adult zebrafish brain. *J. Chem. Neuroanat.* **77**, 176–186 (2016).

147. Stefansson, H. *et al.* Homozygosity for R47H in TREM2 and the Risk of Alzheimer's Disease. *N. Engl. J. Med.* **390**, 2217–2219 (2024).

148. McKhann, G. *et al.* Clinical diagnosis of Alzheimer's disease: report of the NINCDS-ADRDA Work Group under the auspices of Department of Health and Human Services Task Force on Alzheimer's Disease. *Neurology* **34**, 939–944 (1984).

149. Helgason, A., Yngvadóttir, B., Hrafnkelsson, B., Gulcher, J. & Stefánsson, K. An Icelandic example of the impact of population structure on association studies. *Nat. Genet.* **37**, 90–95 (2005).

150. Refsum, H. *et al.* The Hordaland Homocysteine Study: a community-based study of homocysteine, its determinants, and associations with disease. *J. Nutr.* **136**, 1731S-1740S (2006).

151. Kuriyama, S. *et al.* The Tohoku Medical Megabank Project: Design and Mission. *J. Epidemiol.* **26**, 493–511 (2016).

152. Kawai, Y. *et al.* Japonica array: improved genotype imputation by designing a population-specific SNP array with 1070 Japanese individuals. *J. Hum. Genet.* **60**, 581–587 (2015).

153. Ojima, T. *et al.* Body mass index stratification optimizes polygenic prediction of type 2 diabetes in cross-biobank analyses. *Nat. Genet.* **56**, 1100–1109 (2024).

154. Nagai, A. *et al.* Overview of the BioBank Japan Project: Study design and profile. *J. Epidemiol.* **27**, S2–S8 (2017).

155. Akiyama, M. *et al.* Characterizing rare and low-frequency height-associated variants in the Japanese population. *Nat. Commun.* **10**, 4393 (2019).

156. Okada, Y. *et al.* Deep whole-genome sequencing reveals recent selection signatures linked to evolution and disease risk of Japanese. *Nat. Commun.* **9**, 1631 (2018).

157. Manzoni, C. *et al.* Genome-wide analyses reveal a potential role for the MAPT, MOBP, and APOE loci in sporadic frontotemporal dementia. *Am. J. Hum. Genet.* **111**, 1316–1329 (2024).

158. Witoelar, A. *et al.* Meta-analysis of Alzheimer's disease on 9,751 samples from Norway and IGAP study identifies four risk loci. *Sci. Rep.* **8**, 18088 (2018).

159. Medbøen, I. T. *et al.* Cohort profile: the Norwegian Registry of Persons Assessed for Cognitive Symptoms (NorCog) – a national research and quality registry with a biomaterial collection. https://doi.org/10.1136/bmjopen-2021-058810 (2022) doi:10.1136/bmjopen-2021-058810.

160. Eldholm, R. S. *et al.* Progression of Alzheimer's Disease: A Longitudinal Study in Norwegian Memory Clinics. *J. Alzheimer's Dis.* **61**, 1221–1232 (2018).

161. Aarsland, D. *et al.* Frequency and Case Identification of Dementia with Lewy Bodies Using the Revised Consensus Criteria. *Dement. Geriatr. Cogn. Disord.* **26**, 445–452 (2008).

162. Fladby, T. *et al.* Detecting At-Risk Alzheimer's Disease Cases. *J. Alzheimer's Dis.* **60**, 97–105 (2017).

163. Demensstudien i Nord-Norge. https://dsnn.no/dokumentbase.htm.

164. Røen, I. *et al.* Resource Use and Disease Course in dementia - Nursing Home (REDIC-NH), a longitudinal cohort study; design and patient characteristics at admission to Norwegian nursing homes. *BMC Health Serv. Res.* **17**, 365 (2017).

165. TrønderBrain - Department of Neuroscience - NTNU. https://www.ntnu.edu/inb/tronderbrain.

166. Pihlstrøm, L., Morset, K. R., Grimstad, E., Vitelli, V. & Toft, M. A cumulative genetic risk score predicts progression in Parkinson's disease. *Mov. Disord.* **31**, 487–490 (2016).

167. Aalten, P. *et al.* The Dutch Parelsnoer Institute - Neurodegenerative diseases; methods, design and baseline results. *BMC Neurol.* **14**, 254 (2014).

168. Konijnenberg, E. *et al.* Onset of Preclinical Alzheimer Disease in Monozygotic Twins. *Ann. Neurol.* **89**, 987–1000 (2021).

169. Zwan, M. D. *et al.* Dutch Brain Research Registry for study participant recruitment: Design and first results. *Alzheimers Dement. Transl. Res. Clin. Interv.* **7**, e12132 (2021).

170. Marek, K. *et al.* The Parkinson's progression markers initiative (PPMI) – establishing a PD biomarker cohort. *Ann. Clin. Transl. Neurol.* **5**, 1460–1477 (2018).

171. Holstege, H. *et al.* The 100-plus Study of cognitively healthy centenarians: rationale, design and cohort description. *Eur. J. Epidemiol.* **33**, 1229–1249 (2018).

172. Konijnenberg, E. *et al.* The EMIF-AD PreclinAD study: study design and baseline cohort overview. *Alzheimers Res. Ther.* **10**, 75 (2018).

173. Dubois, B. *et al.* Research criteria for the diagnosis of Alzheimer's disease: revising the NINCDS–ADRDA criteria. *Lancet Neurol.* **6**, 734–746 (2007).

174. Jack Jr., C. R. *et al.* NIA-AA Research Framework: Toward a biological definition of Alzheimer's disease. *Alzheimers Dement.* **14**, 535–562 (2018).

175. van der Flier, W. M. & Scheltens, P. Amsterdam Dementia Cohort: Performing Research to Optimize Care. *J. Alzheimer's Dis.* **62**, 1091–1111 (2018).

176. Rubinacci, S., Ribeiro, D. M., Hofmeister, R. J. & Delaneau, O. Efficient phasing and imputation of low-coverage sequencing data using large reference panels. *Nat. Genet.* **53**, 120–126 (2021).

177. Taliun, D. *et al.* Sequencing of 53,831 diverse genomes from the NHLBI TOPMed Program. *Nature* **590**, 290–299 (2021).

178. Li, Y., Willer, C. J., Ding, J., Scheet, P. & Abecasis, G. R. MaCH: using sequence and genotype data to estimate haplotypes and unobserved genotypes. *Genet. Epidemiol.* **34**, 816–834 (2010).

179. Mbatchou, J. *et al.* Computationally efficient whole-genome regression for quantitative and binary traits. *Nat. Genet.* **53**, 1097–1103 (2021).

180. Jansen, I. E. *et al.* Genome-wide meta-analysis identifies new loci and functional pathways influencing Alzheimer's disease risk. *Nat. Genet.* **51**, 404–413 (2019).

181. Åsvold, B. O. *et al.* Cohort Profile Update: The HUNT Study, Norway. *Int. J. Epidemiol.* **52**, e80–e91 (2023).

182. Krokstad, S. *et al.* Cohort Profile: the HUNT Study, Norway. *Int. J. Epidemiol.* **42**, 968–977 (2013).

183. Brumpton, B. M. *et al.* The HUNT study: A population-based cohort for genetic research. *Cell Genomics* **2**, 100193 (2022).

184. Kent, W. J. BLAT--the BLAST-like alignment tool. *Genome Res.* **12**, 656–664 (2002).

185. Zhang, D., Dey, R. & Lee, S. Fast and robust ancestry prediction using principal component analysis. *Bioinforma. Oxf. Engl.* **36**, 3439–3446 (2020).

186. Loh, P.-R. *et al.* Reference-based phasing using the Haplotype Reference Consortium panel. *Nat. Genet.* **48**, 1443–1448 (2016).

187. McCarthy, S. *et al.* A reference panel of 64,976 haplotypes for genotype imputation. *Nat. Genet.* **48**, 1279–1283 (2016).

188. Zhou, W. *et al.* Efficiently controlling for case-control imbalance and sample relatedness in large-scale genetic association studies. *Nat. Genet.* **50**, 1335–1341 (2018).

189. Kunkle, B. W. *et al.* Genetic meta-analysis of diagnosed Alzheimer's disease identifies new risk loci and implicates Aβ, tau, immunity and lipid processing. *Nat. Genet.* **51**, 414–430 (2019).

190. Shigemizu, D. *et al.* Ethnic and trans-ethnic genome-wide association studies identify new loci influencing Japanese Alzheimer's disease risk. *Transl. Psychiatry* **11**, 1–10 (2021).

191. Sherva, R. *et al.* African ancestry GWAS of dementia in a large military cohort identifies significant risk loci. *Mol. Psychiatry* 1–10 (2022) doi:10.1038/s41380-022-01890-3.

192. Milani, L. *et al.* The Estonian Biobank's journey from biobanking to personalized medicine. *Nat. Commun.* **16**, 3270 (2025).

193. Sørensen, E. *et al.* Data Resource Profile: The Copenhagen Hospital Biobank (CHB). *Int. J. Epidemiol.* **50**, 719–720e (2021).

194. Hansen, T. F. *et al.* DBDS Genomic Cohort, a prospective and comprehensive resource for integrative and temporal analysis of genetic, environmental and lifestyle factors affecting health of blood donors. https://doi.org/10.1136/bmjopen-2018-028401 (2019) doi:10.1136/bmjopen-2018-028401.

### Supplementary Author List

#### DBDS Genomic consortium

Karina Banasik[1], Jakob Bay[2], Andrea Barghetti[3], Mette Skou Bendtsen[3], Jens Kjærgaard Boldsen[4], Søren Brunak[5], Nanna Brøns[3], Alfonso Buil Demur[6], Johan Skov Bundgaard[3], Lea Arregui Nordahl Christoffersen[2], Maria Didriksen[3], Khoa Manh Dinh[4], Joseph Dowsett[3], Christian Erikstrup[4,7], Josephine Gladov[4,7], Daniel Gudbjartsson[8], Thomas Folkmann Hansen[9], Dorte Helenius Mikkelsen[6], Lotte Hindhede[4], Henrik Hjalgrim[10,11], Jakob Hjorth von Stemann[3], Bitten Aagaard Jensen[12], Kathrine Kaspersen[4], Bertram Dalskov Kjerulff[4], Lisette Kogelman[9], Mette Kongstad[3], Susan Mikkelsen[4], Christina Mikkelsen[3], Line Hjorth Sjernholm Nielsen[4,7], Janna Nissen[3], Mette Nyegaard[13], Sisse Rye Ostrowski[3,14], Frederikke Byron Pedersen[3], Ole Birger Pedersen[2,14], Liam James Elgaard Quinn[2], Þórunn Rafnar[8], Palle Duun Rohde[13], Klaus Rostgaard[10,11], Andrew Joseph Schork[6], Michael Schwinn[3], Erik Sørensen[3], Kari Stefansson[8], Hreinn Stefánsson[8], Jacob Træholt[3], Unnur Þorsteinsdóttir[8], Mie Topholm Bruun[15], Henrik Ullum[16], Thomas Werge[7,14], David Westergaard[1]

1. Department of Obstetrics and Gynaecology, Copenhagen Univesity Hospital, Hvidovre Hospital, Copenhagen, Denmark
2. Department of Clinical Immunology, Zealand University Hospital, Køge, Denmark
3. Department of Clinical Immunology, Copenhagen University Hospital, Rigshospitalet, Copenhagen, Denmark
4. Department of Clinical Immunology, Aarhus University Hospital, Aarhus, Denmark
5. Novo Nordisk Foundation Center for Protein Research, Faculty of Health and Medical Sciences, University of Copenhagen, Copenhagen, Denmark
6. Institute of Biological Psychiatry, Mental Health Centre, Sct. Hans, Copenhagen University Hospital, Roskilde, Denmark
7. Department of Clinical Medicine, Health, Aarhus University, Aarhus, Denmark
8. deCODE Genetics, Reykjavik, Iceland
9. Danish Headache Center, Department of Neurology, Copenhagen University Hospital, Rigshospitalet-Glostrup, Copenhagen, Denmark
10. Danish Cancer Society Research Center, Copenhagen, Denmark
11. Department of Epidemiology Research, Statens Serum Institut, Copenhagen, Denmark
12. Department of Clinical Immunology, Aalborg University Hospital, Aalborg, Denmark
13. Department of Health Science and Technology, Faculty of Medicine, Aalborg Univeristy, Aalborg, Denmark
14. Department of Clinical Medicine, Faculty of Health and Medical Sciences, University of Copenhagen, Copenhagen, Denmark
15. Department of Clinical Immunology, Odense University Hospital, Odense, Denmark
16. Statens Serum Institut, Copenhagen, Denmark

**Estonian Biobank research team**

Andres Metspalu[1], Lili Milani[1], Tõnu Esko[1], Reedik Mägi[1], Mait Metspalu, [1] Mari Nelis[1], and Georgi Hudjashov[1]

1. Estonian Genome Centre, Institute of Genomics, University of Tartu

**UMCG Genetics Lifelines Initiative (UGLI) group author: LifeLines Cohort Study**

Raul Aguirre-Gamboa[1], Patrick Deelen[1], Lude Franke[1], Jan A Kuivenhoven[2], Esteban A Lopera Maya[1], Ilja M Nolte[3], Serena Sanna[1], Harold Snieder[3], Morris A Swertz[1], Peter M. Visscher[3,4], Judith M Vonk[3], Cisca Wijmenga[1], Naomi Wray[4]

1. Department of Genetics, University of Groningen, University Medical Center Groningen, The Netherlands
2. Department of Pediatrics, University of Groningen, University Medical Center Groningen, The Netherlands
3. Department of Epidemiology, University of Groningen, University Medical Center Groningen, The Netherlands
4. Institute for Molecular Bioscience, The University of Queensland, Brisbane, Queensland, Australia.

**23andMe Research Institute**

Stella Aslibekyan[1], Adam Auton[1], Robert K. Bell[1], Katelyn Kukar Bond[1], Zayn Cochinwala[1], Sayantan Das[1], Kahsaia de Brito[1], Emily DelloRusso[1], Chris Eijsbouts[1], Sarah L. Elson[1], Chris German[1], Julie M. Granka[1], Barry Hicks[1], David A. Hinds[1], Reza Jabal[1], Aly Khan[1], Matthew J. Kmiecik[1], Alan Kwong[1], Yanyu Liang[1], Keng-Han Lin[1], Matthew H. McIntyre[1], Shubham Saini[1], Anjali J. Shastri[1], Jingchunzi Shi[1], Suyash Shringarpure[1], Qiaojuan Jane Su[1], Vinh Tran[1], Joyce Y. Tung[1], Catherine H. Weldon[1], Wanwan Xu[1]

1. 23andMe Research Institute, Los Altos, CA, USA.

**VA Million Veteran Program**

MVP Program Office
Sumitra Muralidhar[1] (Program Director), Jennifer Moser[1] (Associate Director, Scientific Programs), Jennifer E. Deen[1] (Associate Director, Cohort & Public Relations)

MVP Executive Committee
Philip S. Tsao[2] (Co-Chair), Sumitra Muralidhar[1] (Co-Chair), J. Michael Gaziano[3], Elizabeth Hauser[4], Amy Kilbourne[5], Michael Matheny[6], Dave Oslin[7], Deepak Voora[4]

MVP Co-Principal Investigators
J. Michael Gaziano[3], Philip S. Tsao[2]

MVP Core Operations
Jessica V. Brewer[3] (Director, MVP Cohort Operations), Mary T. Brophy[3] (Director, VA Central Biorepository), Kelly Cho[3] (Director, MVP Phenomics), Lori Churby[2] (Director, MVP Regulatory Affairs), Scott L. DuVall[8] (Director, VA Informatics and Computing Infrastructure (VINCI)), Saiju Pyarajan[3] (Director, Data and Computational Sciences), Robert Ringer[9] (Director, VA Albuquerque Central Biorepository), Luis E. Selva[3] (Director, MVP Biorepository Coordination), Shahpoor (Alex) Shayan[3] (Director, MVP PRE Informatics), Brady Stephens[10] (Principal Investigator, MVP Information Center), Stacey B. Whitbourne[3] (Director, MVP Cohort Development and Management)

1. US Department of Veterans Affairs, 810 Vermont Avenue NW, Washington, DC 20420
2. VA Palo Alto Health Care System, 3801 Miranda Avenue, Palo Alto, CA 94304
3. VA Boston Healthcare System, 150 S. Huntington Avenue, Boston, MA 02130
4. Durham VA Medical Center, 508 Fulton Street, Durham, NC 27705
5. VA HSR&D, 2215 Fuller Road, Ann Arbor, MI 48105
6. VA Tennessee Valley Healthcare System, 1310 24th Ave. South, Nashville, TN 37212
7. Philadelphia VA Medical Center, 3900 Woodland Avenue, Philadelphia, PA 19104
8. VA Salt Lake City Health Care System, 500 Foothill Drive, Salt Lake City, UT 84148
9. New Mexico VA Health Care System, 1501 San Pedro Drive SE, Albuquerque, NM 87108
10. Canandaigua VA Medical Center, 400 Fort Hill Avenue, Canandaigua, NY 14424

#### Regeneron Genetics Center

##### RGC Management & Leadership Team

Aris Baras, Gonçalo Abecasis, Adolfo Ferrando, Giovanni Coppola, Andrew Deubler, Luca A Lotta, John D Overton, Jeffrey G Reid, Alan Shuldiner, Katherine Siminovitch, Jason Portnoy, Marcus B Jones, Lyndon Mitnaul, Alison Fenney, Jonathan Marchini, Manuel Allen Revez Ferreira, Maya Ghoussaini, Mona Nafde, William Salerno, Cristen Willer, Lourdes Crane.

##### Sequencing & Lab Operations

John D Overton, Christina Beechert, Erin Fuller, Laura M Cremona, Eugene Kalyuskin, Hang Du, Caitlin Forsythe, Zhenhua Gu, Kristy Guevara, Michael Lattari, Alexander Lopez, Kia Manoochehri, Prathyusha Challa, Manasi Pradhan, Raymond Reynoso, Ricardo Schiavo, Maria Sotiropoulos Padilla, Chenggu Wang, Sarah E Wolf, Hang Du, Kristy Guevara.

##### Genome Informatics & Data Engineering

Jeffrey G Reid, Mona Nafde, Manan Goyal, George Mitra, Sanjay Sreeram, Rouel Lanche, Vrushali Mahajan, Sai Lakshmi Vasireddy, Gisu Eom, Krishna Pawan Punuru, Sujit Gokhale, Benjamin Sultan, Pooja Mule, Mudasar Sarwar, Muhammad Aqeel, Xiaodong Bai, Lance Zhang, Sean O'Keeffe, Razvan Panea, Evan Edelstein, Ayesha Rasool, William Salerno, Evan K Maxwell, Boris Boutkov, Alexander Gorovits, Ju Guan, Lukas Habegger, Alicia Hawes, Olga Krasheninina, Samantha Zarate, Adam J Mansfield, Lukas Habegger.

##### Analytical Genetics and Data Science

Gonçalo Abecasis, Manuel Allen Revez Ferreira, Joshua Backman, Kathy Burch, Adrian Campos, Liron Ganel, Sheila Gaynor, Benjamin Geraghty, Arkopravo Ghosh, Salvador Romero Martinez, Christopher Gillies, Lauren Gurski, Eric Jorgenson, Tyler Joseph, Michael Kessler, Jack Kosmicki, Adam Locke, Priyanka Nakka, Jonathan Marchini, Karl Landheer, Olivier Delaneau, Maya Ghoussaini, Anthony Marcketta, Joelle Mbatchou, Arden Moscati, Anita Pandit, Jonathan Ross, Carlo Sidore, Eli Stahl, Timothy Thornton, Sailaja Vedantam, Rujin Wang, Kuan-Han Wu, Bin Ye, Blair Zhang, Andrey Ziyatdinov, Yuxin Zou, Jingning Zhang, Kyoko Watanabe, Mira Tang, Frank Wendt, Suganthi Balasubramanian, Suying Bao, Kathie Sun, Chuanyi Zhang, Sean Yu, Aaron Zhang, David Corrigan, Dhruv Shidhaye, Chen Wang, Keyrun Adhikari, Alexander Lachmann.

##### Therapeutic Area Genetics

Adolfo Ferrando, Giovanni Coppola, Luca A. Lotta, Alan Shuldiner, Katherine Siminovitch, Brian Hobbs, Jon Silver, William Palmer, Rita Guerreiro, Amit Joshi, Antoine Baldassari, Cristen Willer, Sarah Graham, Ernst Mayerhofer, Erola Pairo Castineira, Mary Haas, Niek Verweij, George Hindy, Jonas Bovijn, Tanima De, Luanluan Sun, Olukayode Sosina, Arthur Gilly, Peter Dornbos, Juan Rodriguez-Flores, Moeen Riaz, Manav Kapoor, Gannie Tzoneva, Momodou W Jallow, Anna Alkelai, Ariane Ayer, Veera Rajagopal, Sahar Gelfman, Vijay Kumar, Jacqueline Otto, Jose Bras, Silvia Alvarez, Jessie Brown, Hossein Khiabanian, Joana Revez, Kimberly Skead, Valentina Zavala, Jae Soon Sul, Lei Chen, Sam Choi, Amy Damask, Nan Lin, Charles Paulding, Sameer Malhotra, Joseph Herman.

**Research Program Management & Strategic Initiatives**
Marcus B. Jones, Michelle G. LeBlanc, Nadia Rana, Jennifer Rico-Varela, Jaimee Hernandez, Larizbeth Romero, Ashley Paynter.

**Senior Partnerships & Business Operations**
Randi Schwartz, Lourdes Crane, Alison Fenney, Jody Hankins, Anna Han, Samuel Hart, Ryan Smith.

**Business Operations & Administrative Coordinators**
Ann Perez-Beals, Gina Solari, Johannie Rivera-Picart, Michelle Pagan, Sunilbe Siceron.

Affiliations:
1. Regeneron Genetics Center, Tarrytown, NY, USA.

**Penn Medicine Biobank (PMBB)**

**PMBB Leadership Team**
Daniel J. Rader, M.D., Marylyn D. Ritchie, Ph.D.
Contribution: All authors contributed to securing funding, study design and oversight. All authors reviewed the final version of the manuscript.

**Patient Recruitment and Regulatory Oversight**
JoEllen Weaver, Nawar Naseer, Ph.D., M.P.H., Giorgio Sirugo, M.D., P.h.D., Afiya Poindexter, Yi-An Ko, Ph.D., Kyle P. Nerz, Jenna Dever, Aidan Harvey, Sydney Linn
Contributions: JW manages patient recruitment and regulatory oversight of study. NN manages participant engagement, assists with regulatory oversight, and researcher access. GS assists with researcher access. AP, YK, KPN, JD, AH, and SH perform recruitment and enrollment of study participants.

**Lab Operations**
JoEllen Weaver, Meghan Livingstone, Fred Vadivieso, Stephanie DerOhannessian, Teo Tran, Julia Stephanowski, Salma Santos, Ned Haubein, P.h.D., Joseph Dunn
Contribution: JW, ML, FV, SD conduct oversight of lab operations. ML, FV, AK, SD, TT, JS, SS perform sample processing. NH, JD are responsible for sample tracking and the laboratory information management system.

**Clinical Informatics**
Anurag Verma, Ph.D., Colleen Morse Kripke, M.S. DPT, MSA, Marjorie Risman, M.S., Renae Judy, B.S., Colin Wollack, M.S.
Contribution: All authors contributed to the development and validation of clinical phenotypes used to identify study subjects and (when applicable) controls.

**Genome Informatics**
Anurag Verma Ph.D., Shefali S. Verma, Ph.D., Scott Damrauer, M.D., Yuki Bradford, M.S., Scott Dudek, M.S., Theodore Drivas, M.D., Ph.D.,
Contribution: AV, SSV, and SD are responsible for the analysis, design, and infrastructure needed to quality control genotype and exome data. YB performs the analysis. TD and AV provides variant and gene annotations and their functional interpretation of variants.

**GHS-RGC DiscovEHR collaboration**

Adam Buchanan[1], David J. Carey[1], Christa L. Martin[1], Michelle Meyer[1], Kyle Retterer[1], David Rolston[1]

1. Geisinger Health System, Danville, PA, USA

**Mayo Clinic-RGC Project Generation**

##### PG Leadership Team

Cerhan, James R., M.D.; Couch, Fergus J., Ph.D., Olson, Janet E., Ph.D.

##### Statistical Genetics and Bioinformatics

Larson, Nicholas B., Ph.D., M.S.; .; Fredericksen, Zachary S.;

##### Laboratory Operations

Cicek, Mine, Ph.D.

##### Registry Principal Investigators

(Alphabetical listing)

1. Alcohol Use Disorder (AUD): Biernacka, Joanna M., Ph.D., Karpyak, Victor M., M.D. Ph.D.
2. Alzheimer's Disease Research Center (ADRC): Vemuri, Prashanthi, Ph.D.; Ramanan, Vijay K., M.D., Ph.D.
3. Bipolar disorder registry: Biernacka, Joanna M., Ph.D., Frye, Mark A., M.D.
4. Brain: Eckel Passow, Jeanette E., Ph.D.
5. Breast - Mayo Florida: McLaughlin, Sarah A., M.D.
6. Breast - Mayo Rochester: Olson, Janet E., Ph.D.; Couch, Fergus J., Ph.D.
7. Cardiovascular Disease Specimen Repository: Bielinski, Suzette J., Ph.D., M.Ed.
8. Chronic Kidney Disease: Lieske, John C., M.D.
9. Chronic Pain: Hooten, W. Michael, M.D.
10. Colorectal: Boardman, Lisa A., M.D.
11. COVID-19 Biobank: Kennedy, Richard B., Ph.D.; Cerhan, James R., M.D., Ph.D.; Badley, Andrew D., M.D.
12. Endometrial: Dowdy, Sean C., M.D.; Bakkum-Gamez, Jamie N., M.D.; Glaser, Gretchen E., M.D.
13. Lung: Yang, Ping, M.D., Ph.D.
14. Lymphoma: Cerhan, James R., M.D., Ph.D.
15. Mayo Clinic Biobank: Olson, Janet E., Ph.D.
16. Mayo Clinic Study of Aging (MCSA): Vemuri, Prashanthi, Ph.D.; Ramanan, Vijay K., M.D., Ph.D.
17. Mayo Mammography Health Study: Vachon, Celine M., Ph.D.
18. Multiple Myeloma (MM)/Smouldering MM (SMM): Dispenzieri, Angela, M.D.; Vachon, Celine M., Ph.D.
19. Neuroendocrine pancreatic tumor registry: Antwi, Samuel O., Ph.D.; Ann L. Oberg, Ph.D.; Kari G. Rabe, MS.
20. Mayo Clinic Biospecimen Resource for Ovarian Cancer Research: Kaufmann, Scott H.,M.D., Ph.D.; Goode, Ellen L., Ph.D.; William A. Cliby, M.D.
21. Biospecimen Resource for Pancreas Research: Antwi, Samuel O., Ph.D.; Ann L. Oberg, Ph.D.; Kari G. Rabe, MS.
22. Parkinson’s Disease: Ahlskog, J. Eric, M.D., Ph.D.; Bower, James H., M.D.
23. Polycystic kidney disease: Harris, Peter C., Ph.D.

24. Polyps: Boardman, Lisa A., M.D.
25. Prevalence of Asymptomatic Ventricular Dysfunction: Pereira, Naveen L., M.D.
26. PRISM Mammography study: Couch, Fergus J., Ph.D.; Vachon, Celine M., Ph.D.; Olson, Janet E., Ph.D.
27. Prostate: Cicek, Mine, Ph.D.
28. Prostate Family: Cicek, Mine, Ph.D.
29. 2Radiation Oncology Registry: Ma, Daniel J., M.D.; Mutter, Robert W., M.D.
30. Renal: Eckel Passow, Jeanette E., Ph.D.
31. Vascular Diseases Biorepository: Kullo, Iftikhar J., M.D.

**Management**

Colborn, Lisa K., M.B.A.; Danielsen, Andrew J.; Harrington, Jonathan J.; Kushwaha, Jennifer M.

**Colorado Center for Personalized Medicine – RGC Collaboration**

Heather D. Anderson[1], Christina L. Aquilante[2], Kelsey Arbogast, Ian M. Brooks[3,4], Elizabeth E. Burke[5], Emily M. Casteel, Joanne B. Cole[3], Curtis R. Coughlin II[6], Jacob Crawford, Kristy Crooks[7], Erin Culver, Matthew J. Fisher, Teresa C. Frye, Hunter George, Chris R. Gignoux[3], Elizabeth K. Gilliland, Casey S. Greene[3], Emily Hearst[5], Audrey E. Hendricks[38], Randi K. Johnson[3,9], Shelby Jones, Dave Kao[10,5], Gabrielle A. Knortz, Danielle Koffenberger, Santhanagopalan Krishnamoorthy, Lisa Ku[11], Elizabeth L. Kudron[3,6], Rashawnda Lacy[4], Ethan M. Lange[3], Joe A. Lesny, Meng Lin[3], James L. Martin

1. Department of Clinical Pharmacy, University of Colorado Skaggs School of Pharmacy and Pharmaceutical Sciences, Anschutz Medical Campus
2. Department of Pharmaceutical Sciences, University of Colorado Skaggs School of Pharmacy and Pharmaceutical Sciences, Anschutz Medical Campus
3. Department of Biomedical Informatics, University of Colorado School of Medicine, Anschutz Medical Campus
4. Health Data Compass, Office of the Vice Chancellor for Health Affairs, Anschutz Medical Campus
5. CARE Innovation Center, UCHealth, Anschutz Medical Campus
6. Department of Pediatrics, University of Colorado School of Medicine, Anschutz Medical Campus
7. Department of Pathology, University of Colorado School of Medicine, Anschutz Medical Campus
8. Department of Mathematical and Statistical Sciences, College of Arts and Sciences, University of Colorado Denver Campus
9. Department of Epidemiology, Colorado School of Public Health, Anschutz Medical Campus
10. Division of Cardiology, Department of Medicine, University of Colorado School of Medicine, Anschutz Medical Campus
11. Hereditary Cancer Clinic, UCHealth, Anschutz Medical Campus

**UCLA-RGC ATLAS collaboration**

Shaiful Alam, Maryam Ariannejad, Yael Berkovich, Paul Boutros, Michael Broudy, Alex Bui, Tim Chang, Chris Denny, Sarah Dry, Albert Duntugan, Dan Geschwind, Roni Haas, Lora Illiev, Ankur Jain, Clara Lajonchere, Clara Magyar, Danielle Martinez, Ghouse Mohammed, Arash Naeim, Stan Nelson, Bogdan Pasaniuc, Yash Patel, Antonia Petruse, Paul Spellman, Paul Tung, Taka Yamaguchi

**INDIANA-CHALASANI**

Naga Chalasani, Tae-Hwi Linus Schwantes-An, Andrew J. Saykin

**Mount Sinai Million Health Discoveries Program**

Alexander W. Charney[1]

1. Icahn School of Medicine at Mount Sinai
